# Therapeutic Anticoagulation in Non-Critically Ill Patients with Covid-19

**DOI:** 10.1101/2021.05.13.21256846

**Authors:** The ATTACC, ACTIV-4a, and REMAP-CAP Investigators, Patrick R. Lawler, Ewan C. Goligher, Jeffrey S. Berger, Matthew D. Neal, Bryan J. McVerry, Jose C. Nicolau, Michelle N. Gong, Marc Carrier, Robert S. Rosenson, Harmony R. Reynolds, Alexis F. Turgeon, Jorge Escobedo, David T. Huang, Charlotte Ann Bradbury, Brett L. Houston, Lucy Z. Kornblith, Anand Kumar, Susan R. Kahn, Mary Cushman, Zoe McQuilten, Arthur S. Slutsky, Keri S. Kim, Anthony C. Gordon, Bridget-Anne Kirwan, Maria M. Brooks, Alisa M. Higgins, Roger J. Lewis, Elizabeth Lorenzi, Scott M. Berry, Lindsay R. Berry, Derek C. Angus, Colin J. McArthur, Steven A. Webb, Michael E. Farkouh, Judith S. Hochman, Ryan Zarychanski

**Author notes:** **Corresponding authors:** Ryan Zarychanski, MD, MSc, Sections of Hematology/Oncology and Critical Care, University of Manitoba, Winnipeg, Manitoba, Canada R3E 0V9, and Judith S. Hochman, MD, MA, New York University Grossman School of Medicine, New York University Langone Health, 530 First Ave., Skirball 9R, New York, NY, USA 10016. **Author and Group Information** The members of the writing committee appear at the end of the main text and the full list of investigators and collaborators appears in the Supplementary Appendix.

## Abstract

**Background:** Thrombo-inflammation may contribute to morbidity and mortality in Covid-19. We hypothesized that therapeutic-dose anticoagulation may improve outcomes in non-critically ill patients hospitalized for Covid-19.

**Methods:** In an open-label adaptive multiplatform randomized controlled trial, non-critically ill patients hospitalized for Covid-19, defined by the absence of critical care-level organ support at enrollment, were randomized to a pragmatic strategy of therapeutic-dose anticoagulation with heparin or usual care pharmacological thromboprophylaxis. The primary outcome combined survival to hospital discharge and days free of organ support through 21 days, which was evaluated with Bayesian statistical models according to baseline D-dimer.

**Results:** The trial was stopped when prespecified criteria for superiority were met for therapeutic-dose anticoagulation in groups defined by high (≥2-fold elevated) and low (<2-fold elevated) D-dimer. Among 2219 participants in the final analysis, the probability that therapeutic anticoagulation increased organ support-free days compared to thromboprophylaxis was 99.0% (adjusted odds ratio 1.29, 95% credible interval 1.04 to 1.61). The adjusted absolute increase in survival to hospital discharge without organ support with therapeutic-dose anticoagulation was 4.6% (95% credible interval 0.7 to 8.1). In the primary adaptive stopping groups, the final probabilities of superiority for therapeutic anticoagulation were 97.3% in the high D-dimer group and 92.9% in the low D-dimer group. Major bleeding occurred in 1.9% and 0.9% of participants randomized to therapeutic anticoagulation and thromboprophylaxis, respectively.

**Conclusions:** In non-critically ill patients with Covid-19, an initial strategy of therapeutic-dose anticoagulation with heparin increases the probability of survival to hospital discharge with reduced use of organ support.

Trial registration numbers: NCT02735707, NCT04505774, NCT04359277, NCT04372589

## Background

The clinical course of Covid-19 is characterized by an initial period of mild to moderate symptoms, followed by progressive respiratory failure and requirement for intensive care unit (ICU)-level organ support or death in some patients.^1,2^ The majority of patients requiring hospitalization are moderately ill, not initially requiring ICU-level organ support.^3-5^ Limited therapies are available to prevent clinical progression to ICU-level organ support and death among moderately ill hospitalized patients.

Patients with Covid-19 have a notable incidence of macro- and microvascular thrombosis and inflammation in association with poor clinical outcomes.^6,7^ Given the anti-thrombotic, anti-inflammatory, and possible anti-viral properties of heparins,^8-10^ it has been hypothesized that anticoagulation with heparin administered at doses higher than conventionally used for venous thromboprophylaxis may improve outcomes.^11^ Further, elevated D-dimer is associated with vascular thrombosis and poor clinical outcomes,^6,12^ and thus some have advocated using D-dimer to guide anticoagulant dosing in patients with Covid-19. In the absence of data from randomized trials, clinical guideline recommendations^13^ and practice^14^ vary widely.

To determine whether an initial strategy of therapeutic-dose anticoagulation with heparin improves survival to hospital discharge with reduced use of ICU-level organ support in hospitalized, non-critically ill patients with Covid-19, we conducted an international, adaptive, multiplatform randomized controlled trial (mpRCT).

## Methods

### Trial Design and Oversight

To accelerate evidence generation, three adaptive randomized controlled trial protocols evaluating therapeutic-dose anticoagulation with heparin in patients hospitalized for Covid-19 were integrated into a single mpRCT. Alignment of eligibility criteria, interventions, outcome measures, and data collection was prospectively undertaken. A joint analysis plan was implemented involving periodic evaluation of statistical stopping criteria (**Protocol Appendix**). Independent data and safety monitoring boards (DSMBs) oversaw the platforms using a collaborative cross-platform DSMB interaction plan. The participating platforms were Antithrombotic Therapy To Ameliorate Complications of Covid-19 (ATTACC; NCT04372589),^15^ Accelerating Covid-19 Therapeutic Interventions and Vaccines-4 Antithrombotics Inpatient platform trial (ACTIV-4a, NCT04505774; which included a vanguard/pilot, NCT04359277), and Randomized, Embedded, Multifactorial Adaptive Platform Trial for Community-Acquired Pneumonia (REMAP-CAP, NCT02735707).^16^ The trial was approved by relevant ethics committees and conducted in accordance with Good Clinical Practice guidelines and the Declaration of Helsinki. Written or verbal informed consent, in accordance with regional regulations, was obtained from all participants or their surrogates. The trial was supported by multiple international funding organizations which had no role in the design, analysis, or reporting of the trial results, apart from the ACTIV-4a protocol which received input on design from the National Institutes of Health professional staff and peer reviewers.

### Participants

The mpRCT enrolled patients hospitalized for Covid-19. The investigators hypothesized that the benefits and risks of therapeutic-dose anticoagulation with heparin may vary according to disease severity. As such, the design prospectively stratified participants into severe (ICU-level of care; critically ill) and moderate (hospitalized; non-critically ill) disease severity states at enrollment. Moderate disease severity was defined as hospitalization for Covid-19 without the requirement for ICU-level of care. ICU-level of care was defined by use of respiratory or cardiovascular organ support (high flow nasal oxygen, non-invasive or invasive mechanical ventilation, vasopressors, or inotropes) in an ICU. In ACTIV-4a, where definitions of an ICU were thought challenging to operationalize during the pandemic, receipt of organ support, irrespective of hospital setting, was used to define ICU-level of care. Participants admitted to an ICU but without the receipt of qualifying organ support were considered moderately ill.

Participants with moderate disease severity were further stratified by their baseline D-dimer: high D-dimer group (D-dimer ≥2 times the upper limit of normal per local assay); low D-dimer group (D-dimer <2 times the upper limit of normal per local assay); or unknown D-dimer group. Patients were ineligible for enrollment beyond 72 hours after hospital admission for Covid-19 or in-hospital SARS-CoV-2 confirmation (ATTACC, ACTIV-4a) or after 14 days following admission (REMAP-CAP). In addition, patients were excluded if discharge was expected within 72 hours, or if they had a clinical indication for therapeutic anticoagulation, high risk for bleeding, required dual antiplatelet therapy, or had a history of heparin allergy including heparin-induced thrombocytopenia (HIT). Detailed exclusion criteria are provided in the **Protocol Appendix**.

### Randomization

Using central web-based systems, participants were randomized to receive an initial pragmatic strategy of therapeutic-dose anticoagulation with heparin or usual care pharmacological thromboprophylaxis in an open label fashion. Therapeutic-dose low molecular weight or unfractionated heparin was administered according to local protocols used for the treatment of acute venous thromboembolism for up to 14 days or until recovery (defined as hospital discharge, or liberation from supplemental oxygen for ≥24 hours). Thromboprophylaxis was provided according to local practice.

A subset of REMAP-CAP participants were randomized in other platform domains, including an antiplatelet domain. Treatment assignments were initially randomized in a 1:1 ratio. The ATTACC and REMAP-CAP designs specified the possibility for response-adaptive randomization, whereby blinded randomization allocation ratios could be modified during the trial based on adaptive analyses to favor allocation of participants to the treatment arm demonstrating greater benefit. In participants with moderate severity Covid-19, response-adaptive randomization was implemented for ATTACC participants on December 15, 2020.

### Outcome Measures

The primary outcome was organ support-free days, an ordinal outcome composed of survival to hospital discharge and, among survivors, the number of days free of ICU-level organ support through day 21. Any death during the index hospitalization through 90 days was assigned –1. This endpoint thus reflects both utilization of critical care therapies and survival. Higher values for organ support-free days indicate better outcomes.

Pre-specified secondary efficacy outcomes included survival, survival without receipt of organ support, survival without receipt of invasive mechanical ventilation, mechanical respiratory support-free days (days free of invasive or non-invasive mechanical ventilation), time-to-hospital discharge, survival without symptomatic major thrombotic events (a composite of freedom from myocardial infarction, pulmonary embolism, ischemic stroke, systemic arterial embolism, and in-hospital death), and survival without any macrovascular thrombotic event (the components of major thrombotic events and symptomatic deep venous thrombosis) (**Supplementary Appendix**). Pre-specified secondary safety outcomes assessed during the treatment period were major bleeding, as defined by the International Society of Thrombosis and Haemostasis,^17^ and laboratory-confirmed HIT. All reported bleeding and thrombotic events were adjudicated in a blinded fashion by clinical endpoints committees using consensus definitions (**Supplementary Appendix**).

### Statistical Analysis

The primary analysis was a Bayesian cumulative logistic model that calculated the posterior probability distribution for the proportional odds ratio for therapeutic-dose anticoagulation compared with thromboprophylaxis on organ support-free days in participants with microbiologically confirmed SARS-CoV-2 infection. The primary model incorporated weakly-informative Dirichlet prior distributions for organ support-free days proportions, and was adjusted for age, sex, site, D-dimer group, and enrollment time period. The model was fit using a Markov Chain Monte Carlo algorithm with 100,000 samples from the joint posterior distribution, allowing calculation of the posterior distributions for the proportional odds ratios, including medians and 95% credible intervals (CrIs), and the posterior probabilities of superiority and futility for therapeutic-dose anticoagulation compared with usual care thromboprophylaxis. In this report, odds ratios >1.0 favor therapeutic-dose anticoagulation.

The primary model estimated treatment effects for each of the disease severity-defined groups (severe and moderate, the latter stratified by baseline D-dimer), utilizing a Bayesian hierarchical approach. The treatment effects of anticoagulation for the groups were nested in a hierarchical prior distribution centered on an overall intervention effect estimated with a neutral prior, but distinct group-specific effects were estimated. When consistent effects were observed for the groups, the posterior distribution for each intervention group effect is shrunk towards the overall estimate (dynamic borrowing).^18^ The stopping criteria for treatment superiority (>99% probability of odds ratio >1.0) and futility (<5% probability of odds ratio >1.2) were evaluated monthly by an independent statistical analysis committee and could be reached within the low and/or high D-dimer groups at each adaptive analysis. Sensitivity analyses of the primary model assuming independent treatment effects (without dynamic borrowing) in each D-dimer group are also reported. Additionally, analyses are presented from the overall moderate severity cohort assuming a single treatment effect irrespective of D-dimer.

Subgroup analyses assessed whether treatment effect varied according to age, sex, baseline respiratory support, and thromboprophylaxis dosing. Protocol adherence was defined by the anticoagulant dose equivalent administered within the first 24-48 hours following randomization (**Supplementary Appendix** for consensus dosing categories). Doses categorized as therapeutic and subtherapeutic heparin qualified as adherent in the therapeutic-dose anticoagulation arm, and low-and intermediate prophylactic-dose anticoagulants qualified as adherent in the thromboprophylaxis arm. The analysis of this dataset was pre-specified in a statistical analysis plan (**Protocol Appendix**).

## Results

The first participant was randomized on April 21, 2020. On January 22, 2021, enrollment was discontinued on advice from the DSMBs after planned adaptive analyses of 1398 moderate severity participants demonstrated that the pre-specified stopping criteria for superiority of therapeutic-dose anticoagulation had been reached in both the high and low D-dimer groups (**Supplementary Appendix Table S1**). By that time, 2245 moderate severity participants had been randomized. The primary analysis population consisted of 2219 participants with SARS-CoV-2 confirmation for whom the primary outcome was known (**Figure 1**). Parallel enrollment of patients with severe Covid-19 ran through December 19, 2020; these results are reported separately.^19^

**Figure 1.**
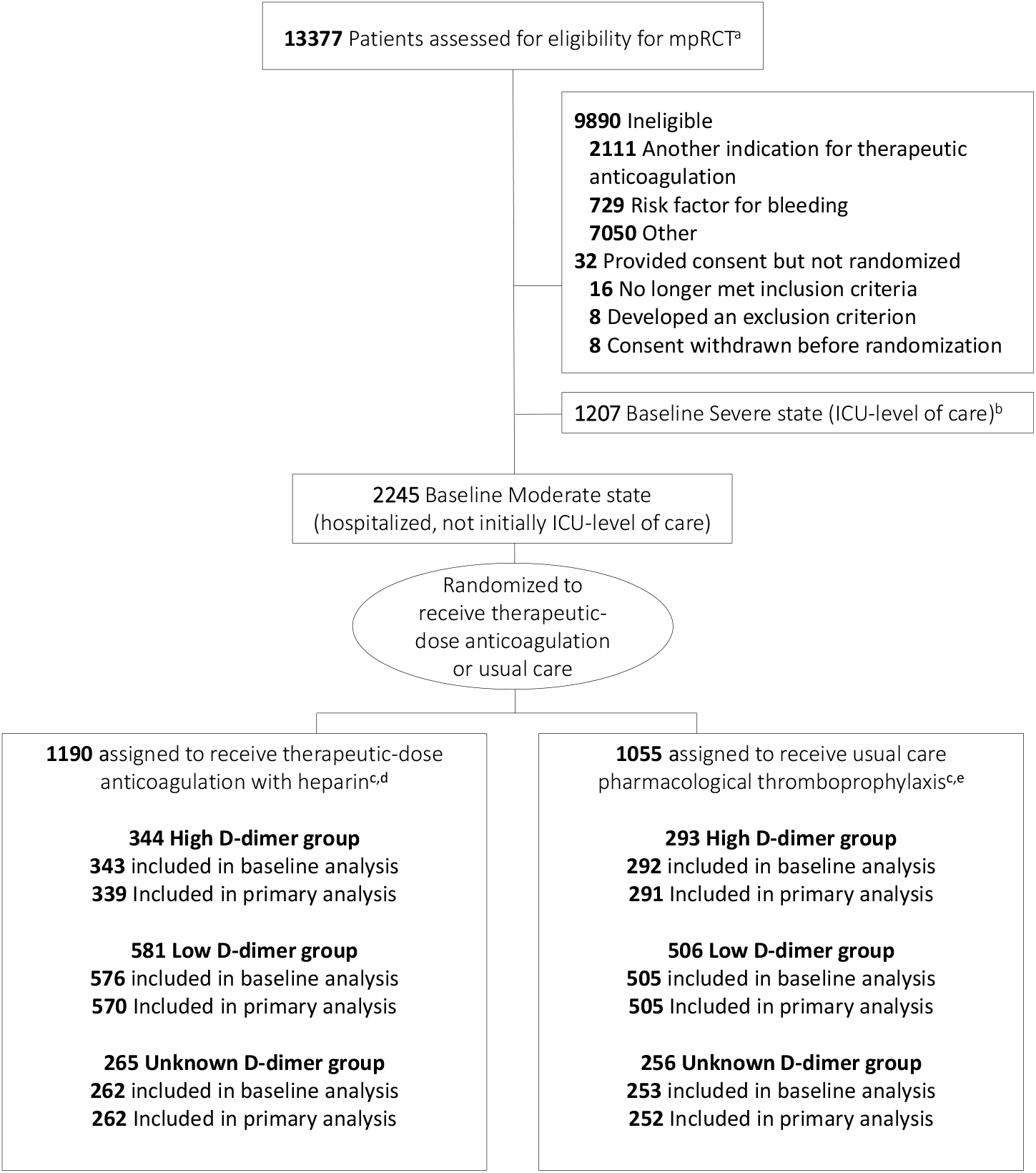
Screening, enrollment, randomization, and inclusion in analysis. **Footnotes**: **a**. sites used varying screening and documentation practices during the pandemic to identify eligible patients (**Protocol Appendix**); as reported, 3799 assessed for eligibility in ACTIV-4a, 7202 assessed for eligibility in ATTACC, and 2372 assessed for eligibility in REMAP-CAP; **b**. potentially used for covariate adjustment and dynamic borrowing; **c**. may be imbalanced due to response adaptive randomization; **d**. reasons (number of participants) for non-inclusion of Moderate severity participants assigned to receive therapeutic anticoagulation in the modified intention to treat primary analysis population included: withdrawal of consent (9), SARS-CoV-2 not confirmed (9), and outcome not available (1); **e**. reasons (number of participants) for non-inclusion of Moderate severity participants assigned to receive usual care pharmacological thromboprophylaxis in the modified intention to treat primary analysis population included: withdrawal of consent (2), SARS-CoV-2 not confirmed (3), and outcome not available (2).

### Participants

Baseline characteristics were similar between treatment arms (**Table 1**), including within each D-dimer group (**Supplementary Appendix Table S2**). Participants in the high D-dimer and unknown D-dimer groups were generally older with a higher prevalence of comorbid diseases compared to those in the low D-dimer group. Concomitant therapies at enrollment included antiplatelet agents (12%), corticosteroids (62%), and remdesivir (36%).

**Table 1.** Demographic and Clinical Characteristics of the Participants at Baseline^**a**^

|  |  |  |
| --- | --- | --- |
| Characteristic | Therapeutic-dose anticoagulation<br>(N=1181)<sup>b</sup> | Usual care pharmacological thromboprophylaxis<br>(N=1050)<sup>b</sup> |
| Age – year | 59.0 (14.1) | 58.8 (13.9) |
| Male Sex – no. (%) | 713/1181 (60.4) | 597/1050 (56.9) |
| Race |  |  |
| White – no. (%) | 622/994 (62.6) | 564/845 (66.7) |
| Asian – no. (%) | 41/994 (4.1) | 43/845 (5.1) |
| Black – no. (%) | 219/994 (22) | 162/845 (19.2) |
| First Nations/American Indian – no. (%) | 118/965 (12.2) | 82/819 (10.0) |
| Other – no. (%) | 17/1109 (1.5) | 16/968 (1.7) |
| Hispanic or Latino ethnicity – no. (%) | 574/1004 (57.2) | 537/879 (61.1) |
| Body mass index, kg/m<sup>2</sup> | 29.8 (26.3–34.7) | 30.3 (26.7–34.9) |
|  | N = 979 | N = 860 |
| Pre-existing conditions |  |  |
| Hypertension – no. (%) | 546/1023 (53.4) | 447/892 (50.1) |
| Diabetes mellitus – no. (%) | 352/1181 (29.8) | 311/1049 (29.6) |
| Severe cardiovascular disease<sup>c</sup> – no. (%) | 123/1165 (10.6) | 123/1165 (10.6) |
| Chronic kidney disease – no. (%) | 83/1173 (7.1) | 69/1037 (6.7) |
| Chronic respiratory disease<sup>d</sup> – no. (%) | 249/1132 (22) | 212/988 (21.5) |
| Immunosuppressive disease – no. (%) | 105/1143 (9.2) | 103/1005 (10.2) |
| Baseline treatments |  |  |
| Antiplatelet agent<sup>e</sup> – no. (%) | 148/1140 (13.0) | 111/1013 (11.0) |
| Remdesivir – no. (%) | 428/1178 (36.3) | 383/1048 (36.5) |
| Corticosteroids – no. (%) | 479/791 (60.6) | 415/656 (63.3) |
| Tocilizumab – no. (%) | 6/1178 (0.5) | 7/1048 (0.7) |
| Baseline respiratory support – n/N (%) |  |  |
| None – no. (%) | 156/1181 (13.2) | 123/1050 (11.7) |
| Low flow nasal cannula/face mask – no. (%) | 789/1181 (66.8) | 696/1050 (66.3) |
| High flow nasal cannula – no. (%) | 25/1181 (2.1) | 28/1050 (2.7) |
| Non-invasive mechanical ventilation – no. (%) | 21/1181 (1.8) | 24/1050 (2.3) |
| Unspecified<sup>f</sup> – no. (%) | 190/1181 (16.1) | 179/1050 (17.1) |
| Laboratory values |  |  |
| D-dimer relative to site upper limit of normal | 1.6 (0.9–2.6)<br>N = 900 | 1.5 (1–2.7)<br>N = 779 |
| Platelets x10<sup>9</sup>/L | 221 (171–290)<br>N = 1160 | 218 (172.5–289)<br>N = 1031 |
| Lymphocytes x10<sup>9</sup>/L | 0.9 (0.7–1.3)<br>N = 1032 | 1 (0.7–1.4)<br>N = 908 |
| Creatinine (mg/dL) | 0.9 (0.7–1.1)<br>N = 1144 | 0.9 (0.7–1.1)<br>N = 1012 |
| Platform of enrollment<sup>g</sup> |  |  |
| ATTACC<sup>h</sup> – no. (%) | 650/1181 (55.0) | 509/1050 (48.5) |
| ACTIV-4a – no. (%) | 387/1181 (32.8) | 392/1050 (37.3) |
| REMAP-CAP – no. (%) | 144/1181 (12.2) | 149/1050 (14.2) |
| Country of enrollment |  |  |
| United Kingdom – no. (%) | 95/1181 (8) | 103/1049 (9.8) |
| United States – no. (%) | 573/1181 (48.5) | 506/1049 (48.2) |
| Canada – no. (%) | 102/1181 (8.6) | 83/1049 (7.9) |
| Brazil – no. (%) | 234/1181 (19.8) | 209/1049 (19.9) |
| Other<sup>i</sup> – no. (%) | 177/1181 (15) | 148/1049 (14.1) |

Median [IQR] or proportions. **Abbreviations:** No. = number.

Adherence to protocol-assigned anticoagulation dose on the day following randomization was 88.3% in the therapeutic-dose anticoagulation arm and 98.3% in the thromboprophylaxis arm (**Supplementary Appendix Table 3**). Among participants randomized to therapeutic-dose heparin, 94.7% (1035/1093) received a low molecular weight heparin, most commonly enoxaparin. Among participants allocated to usual care thromboprophylaxis, 71.7% (613/855) received low-dose and 26.5% (227/855) received intermediate-dose thromboprophylaxis.

### Primary Outcome

Among 2219 participants in the overall moderate severity cohort, the posterior probability that therapeutic-dose anticoagulation increased organ support-free days compared to usual care thromboprophylaxis was 99.0% (median adjusted odds ratio 1.29, 95% CrI 1.04 to 1.61) (**Figure 2** and **Figure 3**). The proportion of participants in the thromboprophylaxis arm surviving to hospital discharge without receipt of organ support during the first 21 days (control event frequency) was 76.4% (247/1048). The median adjusted absolute improvement in this proportion with therapeutic-dose anticoagulation was 4.6% (95% CrI 0.7 to 8.1). In the primary adaptive analysis groups, the final posterior probability for superiority of therapeutic-dose anticoagulation compared with usual care thromboprophylaxis was 97.3% (adjusted odds ratio 1.31, 95% CrI 1.00 to 1.76) in the high D-dimer group (n=630) and 92.9% (adjusted odds ratio 1.22, 95% CrI 0.93 to 1.57) in the low D-dimer group (n=1075) (**Figure 3** and **Supplementary Appendix Figure S1**). The posterior probability of superiority of therapeutic-dose anticoagulation in the unknown D-dimer group (n=514) was 97.3% (adjusted odds ratio 1.32, 95% CrI 1.00 to 1.86). The results were consistent in sensitivity analyses (**Supplementary Appendix Table S4**).

**Figure 2.**
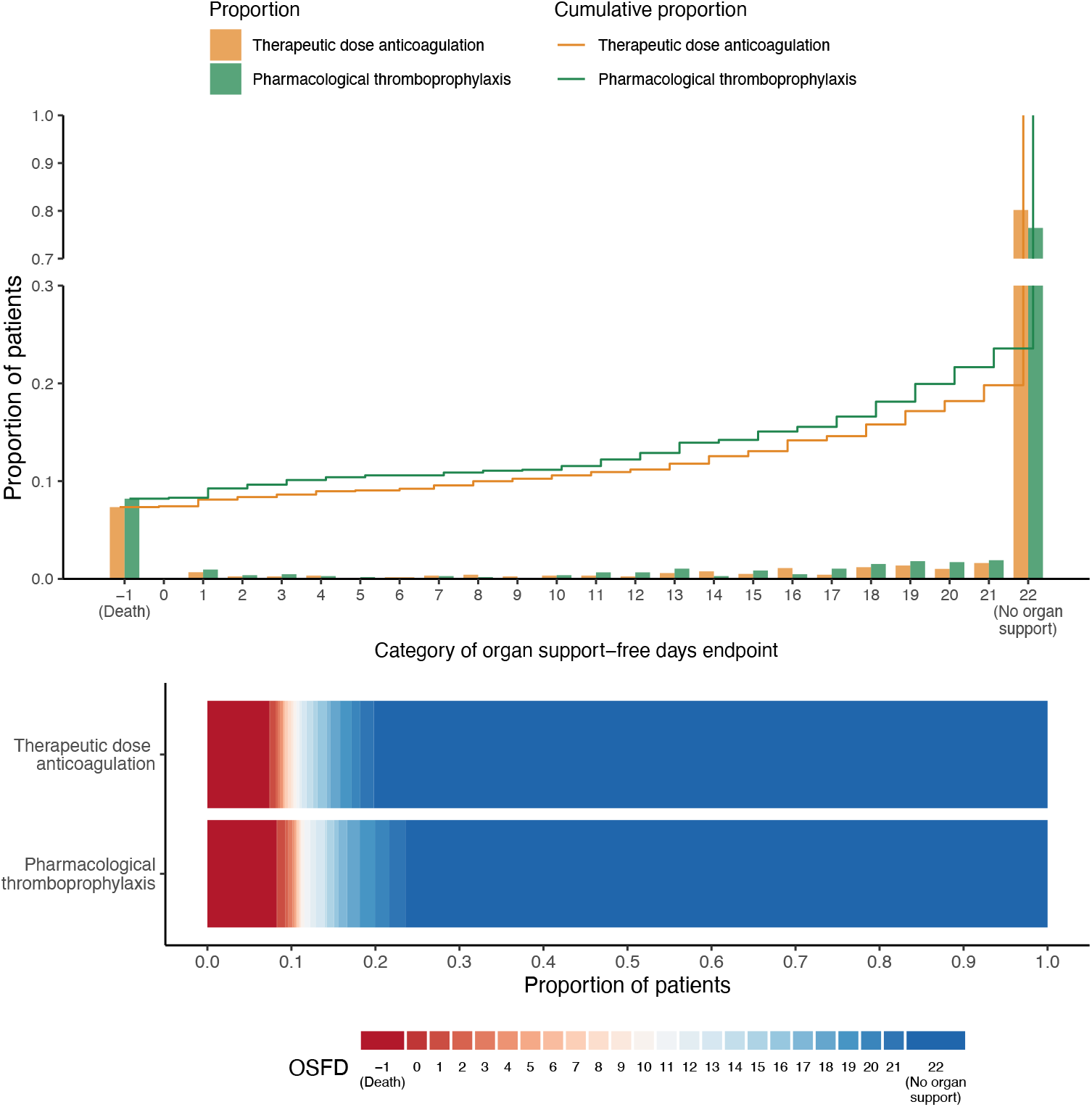
Organ support-free days to day 21. **Upper panel**) the cumulative proportion (y-axis) for each intervention group along the ordinal scale (x-axis). The ordinal scale ranges from –1 (in-hospital death; the worst possible outcome) through 0 to 21 (among survivors, the numbers of days alive without organ support; intermediate outcomes), to 22 (survival to hospital discharge without receipt of organ support; the best possible outcome). The height of each curve at any point, for example, at day = 10, indicates the proportion of participants with organ support-free days (OSFD) of 10 or lower (i.e., 10 or worse). The difference in height of the two curves at any point represents the difference in the cumulative probability of having a value for OSFDs less than or equal to that point on the x-axis. **Lower panel**) Organ support-free days are shown as horizontally stacked proportions by intervention group, with possible outcomes as: in-hospital death with or without the receipt of organ support (dark red; the worst possible outcome, corresponding to an ordinal scale score of -1); survival, requiring ICU-level organ support (red to blue gradient shading based on number of days alive without organ support; intermediate outcomes, corresponding to an ordinal scale scores of 0-21); and survival to hospital discharge, without requiring ICU-level organ support (dark blue; the best possible outcome, corresponding to an ordinal scale score of 22). The median adjusted proportional odds ratio for therapeutic dose vs. thromboprophylaxsis among all moderate severity participants in the modified intention-to-treat analysis was 1.29 (95% credible interval 1.04 to 1.61; posterior probability of superiority 99.0%).

**Figure 3.**
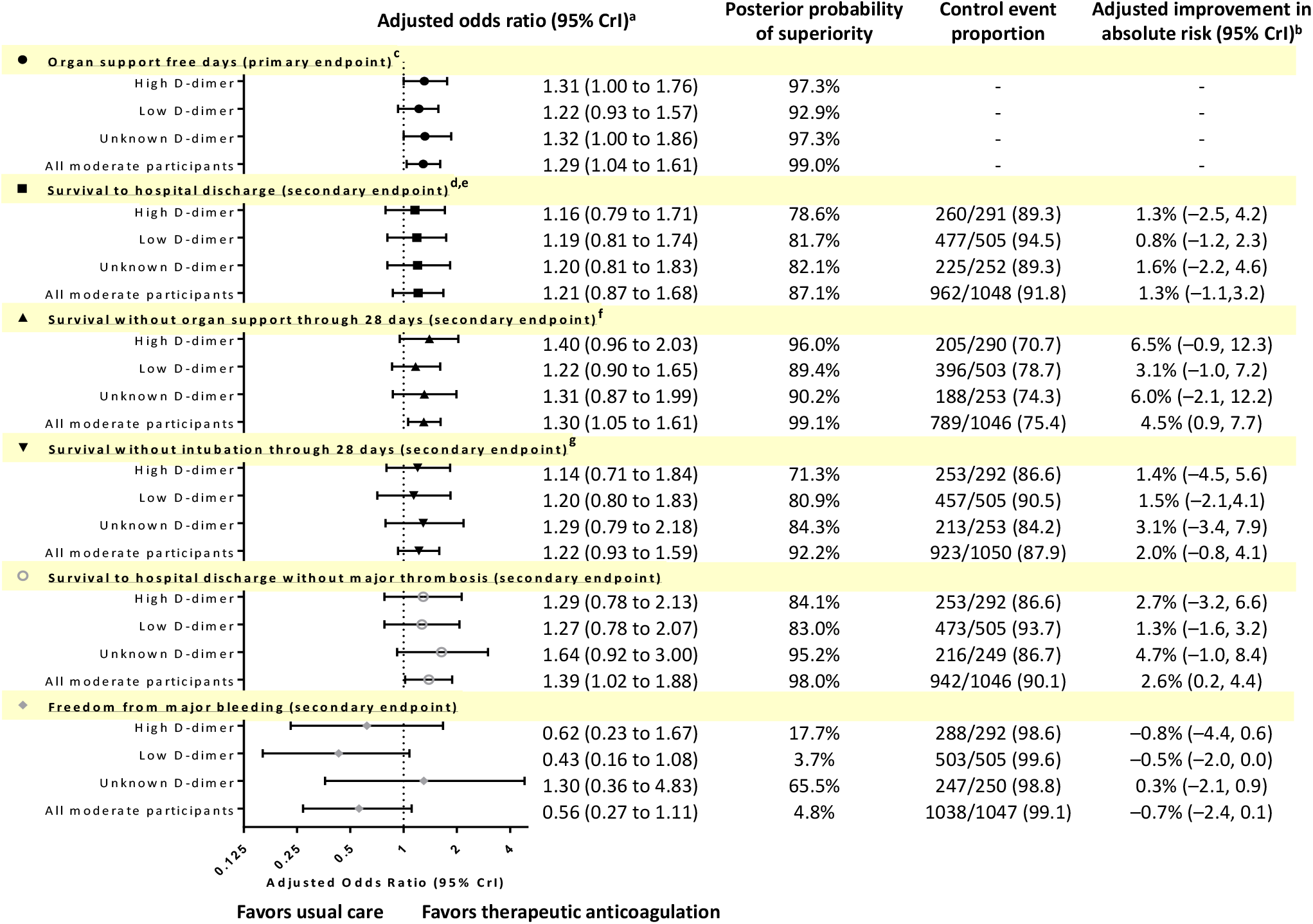
Primary and secondary outcomes. Median adjusted proportional odds ratios and 95% credible intervals are shown, along with the corresponding probabilities for superiority (adjusted odds ratio >1.0), the control event proportion, and the mean adjusted improvement in absolute risk. Forest plots are shown such that values >1.0 favor therapeutic heparin and values <1.0 favor usual care pharmacological thromboprophylaxis. The High D-dimer group is defined as moderate severity participants with baseline D-dimer ≥2 times local upper limit of normal for assay, the Low D-dimer group as moderate severity participants with baseline D-dimer <2 times local upper limit of normal for assay, and the baseline Unknown D-Dimer defined as moderate severity participants without available baseline D-dimer. Additional secondary endpoints are reported in **Figure 4** and **Supplementary Appendix Table S5. Abbreviations**: CrI = credible interval. **Footnotes**: **a**. effect estimates are adjusted for age, sex, site, D-dimer group, and time epoch, and are computed so that a value greater than 1 indicates benefit from therapeutic anticoagulation; **b**. calculated from the adjusted treatment effect and control event frequency; **c**. the primary analysis includes 630 High D-dimer moderate severity participants, 1075 Low D-dimer moderate severity participants, and 514 Unknown D-dimer moderate severity participants; the primary analysis uses dynamic borrowing across illness severity and D-dimer groups, whereby observations from one group are used to inform treatment effect estimation in other groups where effect is observed to be similar (results from a sensitivity analysis assuming independent treatment effects between D-dimer-defined groups are shown in **Supplementary Appendix Table S5**); treatment effects also reported for the overall moderate severity cohort (n=2219) assuming a single treatment effect irrespective of D-dimer; **d**. includes dynamic borrowing between D-dimer groups as derives from the primary model; other secondary outcomes do not employ dynamic borrowing; **e**. similar results were obtained from a time-to-event model of survival through 28 days: adjusted hazard ratio of 1.20 (95% CrI, 0.88-1.61; posterior probability of superiority 87.8%); **f**. dichotomous endpoint; similar results obtained following the exclusion of 52 participants receiving organ support at baseline (median adjusted odds ratio 1.30, 95% CrI 1.06-1.62; posterior probability of superiority 99.3%); **g**. ordinal outcome with death as the worst possible outcome.

In the overall moderate severity cohort, the treatment effect did not vary meaningfully by age, respiratory support at enrollment, or thromboprophylaxis dosing. There was a 95.5% probability that the odds ratio associated with therapeutic-dose anticoagulation was higher in men compared to women (**Supplementary Appendix Figure S2**).

### Secondary Outcomes

Secondary outcomes are shown in **Figure 3, Figure 4**, and **Supplementary Appendix Table S5**. In the overall moderate severity cohort, the posterior probabilities that therapeutic-dose anticoagulation increased survival without organ support or survival without invasive mechanical ventilation through 28 days were 99.1% and 92.2%, respectively. The posterior probability that therapeutic-dose anticoagulation improved survival to hospital discharge compared with thromboprophylaxis was 87.1% (median adjusted odds ratio 1.21, 95% CrI 0.87 to 1.68; median adjusted absolute improvement 1.3%, 95%CrI -1.1 to 3.2%).

**Figure 4.**
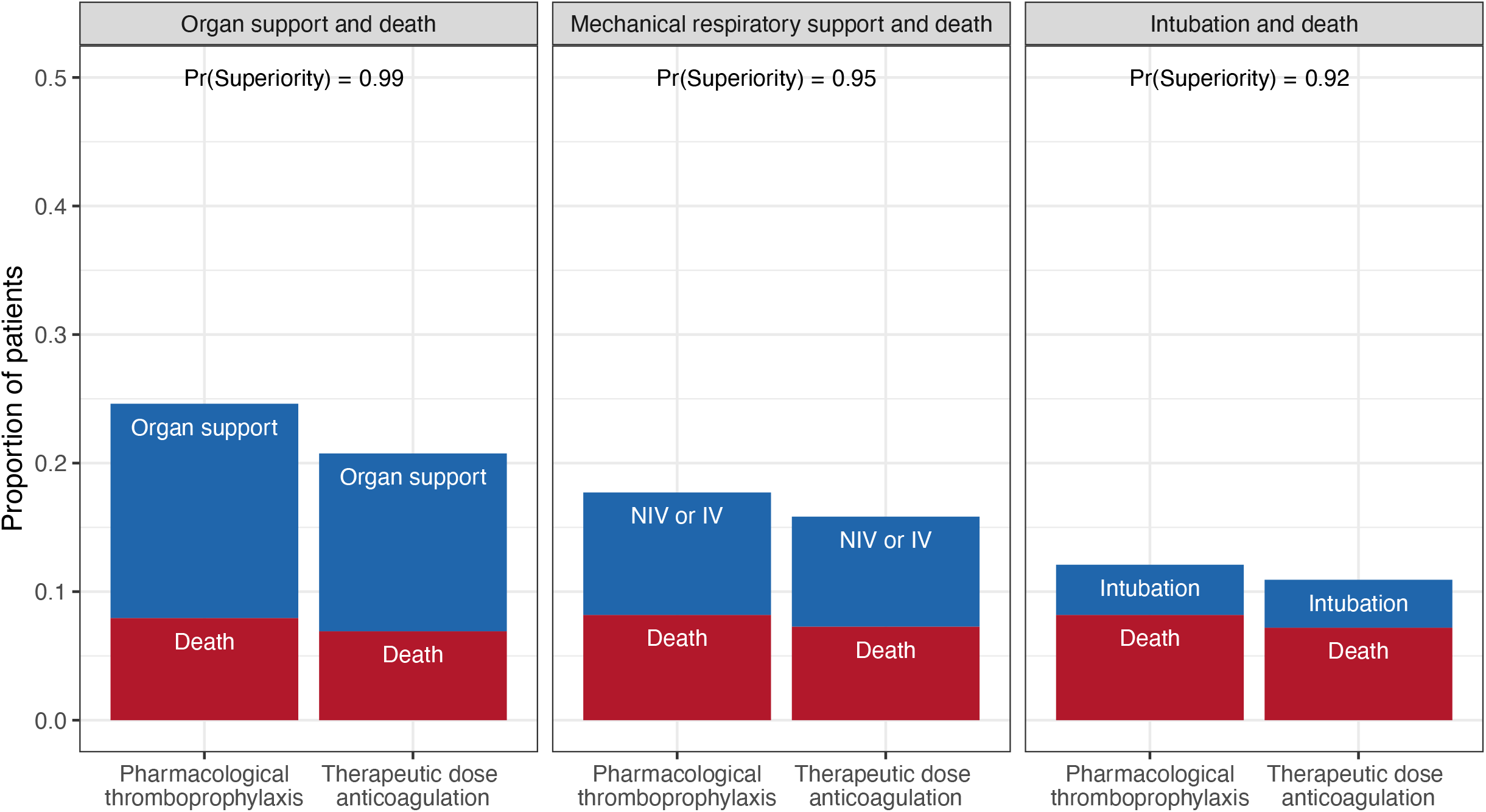
Effect of therapeutic anticoagulation on mortality, organ and respiratory support, in the overall moderate severity cohort. Unadjusted proportions are shown by treatment group. The posterior probability of superiority of therapeutic anticoagulation with heparin in comparison to usual care thromboprophylaxis is shown for the combined probabilities of death and receipt of either organ support, or the subsets of mechanical respiratory support (non-invasive or invasive mechanical ventilation), or intubation. **Abbreviations**: IV = invasive ventilation; NIV = non-invasive ventilation; Pr = probability. **Footnote**: Models analyzed as follows: survival without organ support through 28 days (dichotomous outcome); mechanical respiratory support-free days (ordinal outcome based on days free of support, with in-hospital death assigned as 0) through 28 days; and survival free of invasive mechanical ventilation through 28 days (ordinal, death as the worse outcome).

A major thrombotic event or death in-hospital occurred in 8.0% (94/1180) of participants randomized to therapeutic-dose anticoagulation and 9.9% (104/1046) of participants randomized to thromboprophylaxis (**Figure 3** and **Supplementary Appendix Tables S5** and **S6**). The composite secondary thrombosis outcome including deep vein thrombosis is reported in **Supplementary Appendix Table S5**. Major bleeding occurred in 1.9% (22/1180) of participants randomized to therapeutic-dose anticoagulation and 0.9% (9/1047) of participants randomized to thromboprophylaxis (**Supplementary Appendix Table S7**). Fatal bleeding occurred in three participants randomized to therapeutic-dose anticoagulation and one participant randomized to thromboprophylaxis. No episodes of intracranial bleeding or confirmed HIT occurred during the treatment window.

## Discussion

In this randomized trial report of non-critically ill patients hospitalized for Covid-19, therapeutic-dose anticoagulation with heparin increased the probability of surviving to hospital discharge with reduced need for ICU-level organ support, including invasive and non-invasive mechanical ventilation. Therapeutic-dose anticoagulation with heparin was beneficial irrespective of baseline D-dimer. Major bleeding occurred infrequently.

Several non-randomized cohort studies have observed favorable associations between anticoagulant use and survival from Covid-19, although these studies are at risk of bias.^20-22^ Because SARS-CoV-2 infection incites a dysregulated inflammatory response that may cascade to activation of coagulation and widespread thrombin generation,^23^ potentially contributing to organ failure,^24-26^ heparins may reduce the requirement for organ support by limiting thrombo-inflammation through anti-thrombotic, anti-inflammatory, and potentially anti-viral mechanisms.^8-10,27^ The results from this mpRCT show that empiric therapeutic-dose anticoagulation with heparin provided to patients hospitalized prior to the receipt of ICU-level organ support improved outcomes.

In contrast, in a parallel enrolling group of the mpRCT comprised of critically-ill patients (receiving ICU-level of care at enrollment), empiric therapeutic-dose anticoagulation was not beneficial.^19^ In a separate randomized trial in critically-ill patients with Covid-19, intermediate-dose heparin was likewise not beneficial.^28^ Differential treatment effects based on illness severity and the time course of SARS-CoV-2 infection have been reported in randomized trials of other Covid-19 therapies.^29-31^ It is possible that therapeutic-dose heparin is unable to influence the thrombo-inflammatory cascade and organ injury in patients with advanced disease and its sequale.^32-34^ Among moderately-ill participants in this mpRCT, those in the high D-dimer group were at increased risk for mortality and receipt of organ support compared with those in the low D-dimer group and, accordingly, adjusted absolute treatment benefits were more apparent. Participants in the high D-dimer group were generally older with a higher prevalence of comorbid diseases.

Strengths of our trial design include elements incorporated to speed evidence generation. First, we adopted a novel prospective mpRCT design, whereby three collaborating platforms developed and administered harmonized trial protocols across complementary global networks of sites to increase efficiency and reliability of evidence generation. Second, the trial employed an adaptive Bayesian design which accounted for uncertainties in the pandemic. This approach allowed for trial conclusions to be reached simultaneously or sequentially in groups defined by illness severity and D-dimer through periodic adaptive analyses. Dynamic borrowing was incorporated to enable the trial to reach conclusions more quickly across the stopping groups where treatment effects were similar, and to mitigate the influence of outlying treatment effects by shrinking similar treatment estimates together. Response-adaptive randomization allowed randomization probabilities to be modified as the trial acquired knowledge about treatment effects. Response-adaptive randomization may lead to imbalances in baseline covariates between treatment arms over time, and as such the primary models of treatment effect are necessarily adjusted for age, sex, site, D-dimer group, and time. Given the importance of accounting for these factors under this design, absolute risk reductions based on adjusted treatment effects and observed control event rates are presented. The adjusted absolute improvement in outcomes presented is a median; patients at higher baseline risk, including older age, male sex, and higher baseline D-dimer may derive greater absolute benefit.

Although the open-label design of the mpRCT represents a potential limitation, the primary outcome involving survival and receipt of organ support was selected to minimize bias and to function across a spectrum of illness severity. The potential for ascertainment bias cannot be excluded for the secondary outcomes of major bleeding or thrombosis. This, along with the absence of protocolized screening for venous thrombosis, exclusion of patients at increased bleeding risk, and changing disease epidemiology over time may have contributed to lower thrombotic event rates than have been previously reported.^35^ The treatment effect was attenuated in the final analysis relative to the adaptive stopping results; nevertheless, a high probability of benefit persisted in all D-dimer groups.

In conclusion, in hospitalized, non-critically ill patients with Covid-19, an initial strategy of therapeutic-dose anticoagulation with heparin improves organ support-free days. Therapeutic-dose anticoagulation increases the probability of survival to hospital discharge with reduced use of critical care-level organ support.

## Supporting information

Supplementary Appendix AC mpRCT Moderate

## Data Availability

The data is not available

## Acknowledgements

We are grateful for the support of the patients and their families who participated in this trial. We thank those who served on the Data Safety and Monitoring Boards of each platform; we gratefully acknowledge the support of multiple funding organizations for the participating platforms (**Supplementary Appendix**).

## Authors

### Executive Writing Committee

Patrick R. Lawler, M.D., M.P.H.^*1,2^, Ewan C. Goligher, M.D., Ph.D.^*2,3^, Jeffrey S. Berger, M.D.^*4^, Matthew D. Neal, M.D.^*5,6^, Bryan J. McVerry, M.D^*5,6^, Jose C. Nicolau, M.D., Ph.D.^7^, Michelle N. Gong, M.D.^8,9^, Marc Carrier, M.D., M.Sc.^10,11^, Robert S. Rosenson, M.D.^12,13^, Harmony R. Reynolds, M.D.^4^, Alexis F. Turgeon, M.D., M.Sc.^14,15^, Jorge Escobedo, M.D.^16^, David T. Huang, M.D., M.P.H.^5^, Charlotte Ann Bradbury, M.B., Ch.B., Ph.D.^17,18^, Brett L. Houston, M.D.^19,20^, Lucy Z. Kornblith, M.D.^21^, Anand Kumar, M.D.^19^, Susan R. Kahn, M.D., M.Sc.^22^, Mary Cushman, M.D., M.Sc.^23^, Zoe McQuilten, Ph.D.^24^, Arthur S. Slutsky, M.D.^2,25^, Keri S. Kim, Pharm. D.^26^, Anthony C. Gordon, M.B.B.S., M.D.^27,28^, Bridget-Anne Kirwan, Ph.D.^29,30^, Maria M. Brooks, Ph.D.^5^, Alisa M. Higgins, Ph.D.^24^, Roger J. Lewis, M.D., Ph.D.^31,32^, Elizabeth Lorenzi, Ph.D.^31^, Scott M. Berry, Ph.D.^31^, Lindsay R. Berry, Ph.D.^31^, Derek C. Angus, M.D., M.P.H.^5,6^, Colin J. McArthur, M.B., Ch.B.^**24,33,34^, Steven A. Webb, M.P.H., Ph.D.^**24,35^, Michael E. Farkouh, M.D., M.Sc.^**1,2^, Judith S. Hochman, M.D., M.A.^* *4^, Ryan Zarychanski, M.D., M.Sc.^* *19,20^.

^*. * *^denotes equal contribution

### Block Writing Committee

*(In alphabetical order)* Aaron W. Aday, M.D.^36^, Farah Al-Beidh, Ph.D.^27^, Djillali Annane, M.D., Ph.D.^37^, Yaseen M. Arabi, M.D.^38,39^, Diptesh Aryal, M.D.^40,41^, Lisa Baumann Kreuziger, M.D.^42^, Abi Beane, Ph.D.^43,44^, Zahra Bhimani, M.P.H.^25^, Shailesh Bihari, Ph.D.^45^, Henny H. Billett, M.D., M.Sc.^8,9^, Lindsay Bond ^46^, Marc Bonten, Ph.D.^47^, Frank Brunkhorst^48^, Meredith Buxton, Ph.D.^49^, Adrian Buzgau**Error! Bookmark not defined**., Lana A. Castellucci, M.D., M.Sc.^10,50^, Sweta Chekuri, M.D.^8^, Jen-Ting Chen, M.D., M.S.^8^, Allen C. Cheng, Ph.D.^51,52^, Tamta Chkhikvadze, M.D.^4,53^, Benjamin Coiffard, M.D., M.Sc.^54^, Todd W. Costantini, M.D.^55^, Sophie de Brouwer, Ph.D.^32^, Lennie P. G. Derde, M.D., Ph.D.^47^, Michelle A. Detry, Ph.D.^31^, Abhijit Duggal, M.D., M.P.H., M.Sc.^56^, VladimÍr DžavÍk, M.D.^1,2^, Mark B. Effron, M.D.^57^, Lise J. Estcourt, M.B.B.Chir., D.Phil.^42,58^, Brendan M. Everett, M.D., M.P.H.^59^, Dean A. Fergusson, Ph.D.^10,50^, Mark Fitzgerald, Ph.D.^31^, Robert A. Fowler, M.D.^2^, Jean Philippe Galanaud, M.D.^1,60^, Benjamin T. Galen, M.D.^9^, Sheetal Gandotra, M.D.^61^, Sebastian GarcÍa-Madrona, M.D.^62^, Timothy D. Girard, M.D.^63^, Lucas C. Godoy, M.D.^1,2,7^, Andrew L. Goodman, M.D.^64^, Herman Goossens, M.D.^65^, Cameron Green, M.Sc.^24^, Yonatan Y. Greenstein, M.D.^66^, Peter L. Gross, M.D., M.Sc.^67,68^, Naomi M. Hamburg, M.D., M.Sc.^69,70^, Rashan Haniffa, Ph.D.^71,72^, George Hanna, M.D.^1,2^, Nicholas Hanna, M.D.^73,74^, Sheila M. Hegde, M.D., M.P.H.^75^, Carolyn M. Hendrickson, M.D.^21^, R. Duncan Hite, M.D.^78^, Alexander A. Hindenburg, M.D.^76^, Aluko A. Hope, M.D., M.S.C.E.^9^, James M. Horowitz, M.D.^53^, Christopher M. Horvat, M.D., M.H.A.^77^, Kristin Hudock, M.D., M.S.T.R.^78^, Beverley J. Hunt, O.B.E.^79^, Mansoor Husain, M.D.^2,3^, Robert C. Hyzy, M.D.^80^, Vivek N. Iyer, M.D., M.P.H.^81^, Jeffrey R. Jacobson, M.D.^26^, Devachandran Jayakumar, M.D.^82^, Norma M. Keller, M.D.^4,83^, Akram Khan, M.D.^84^, Yuri Kim, M.D., Ph.D.^59^, Andrei L. Kindzelski,M.D., Ph.D.^85^, Andrew J. King, Ph.D.^5^, M. Margaret Knudson, M.D.^21^, Aaron E. Kornblith, M.D.^21^, Vidya Krishnan, M.D., M.H.S.^86^, Matthew E. Kutcher, M.D., M.S.^87^, Michael A. Laffan, D.M.^27^, Francois Lamontagne, M.D.^88^, Grégoire Le Gal, M.D., Ph.D.^10,11^, Christine M. Leeper, M.D., M.Sc.^5^, Eric S. Leifer, Ph.D.^84^, George Lim, M.D.^89^, Felipe Gallego Lima, M.D.^7^, Kelsey Linstrum, M.S.^5,6^, Edward Litton, Ph.D.^35,90^, Jose Lopez-Sendon, Ph.D.^91^, Jose Luis Lopez-Sendon Moreno, M.D.^62^, Sylvain A. Lother, M.D.**Error! Bookmark not defined**., Saurabh Malhotra, M.D., M.P.H.^92,93^, Miguel Marcos, Ph.D.^94^, Andréa Saud Marinez, Pharm.D., M.Sc.^95^, John C. Marshall, M.D.^25^, Nicole Marten, R.N.^96^, Michael A. Matthay, M.D.^21^, Daniel F. McAuley, M.D.^97,98^, Emily G. McDonald, M.D., M.Sc.^22^, Anna McGlothlin, Ph.D.^31^, Shay P. McGuinness, M.B., Ch.B.^24,99^, Saskia Middeldorp, M.D., Ph.D.^100^, Stephanie K. Montgomery, M.Sc.^5,6^, Steven C. Moore, M.D.^101^, Raquel Morillo Guerrero, Ph.D.^62^, Paul R. Mouncey, M.Sc.^102^, Srinivas Murthy, M.D.^103^, Girish B. Nair, M.D., M.S.^104,105^, Rahul Nair, M.D.^8^, Alistair D. Nichol, M.B., Ph.D.^24,52,106^, Brenda Nunez-Garcia, B.A.^21^, Ambarish Pandey, M.D.^107^, Pauline K. Park, M.D.^80^, Rachael L. Parke, Ph.D.^99,108^, Jane C. Parker, B.N.^24^, Sam Parnia, M.D., Ph.D.^4^, Jonathan D. Paul, M.D.^109^, Yessica Sara Pérez González, M.D.^16^, Mauricio Pompilio, Ph.D.^110,111^, Matthew E. Prekker, M.D., M.P.H.^112^, John G. Quigley, M.D.^26^, Natalia S. Rost, M.D.^113,114^, Kathryn Rowan, Ph.D.^102^, Fernanda O. Santos, M.D.^115^, Marlene Santos, M.D., M.Sc.^25^, Mayler Olombrada Santos, M.Sc.^116^, Lewis Satterwhite, M.D.^117^, Christina T. Saunders, Ph.D.^31^, Roger E.G. Schutgens, M.D., Ph.D.^47^, Christopher W. Seymour, M.D., M.Sc.**Error! Bookmark not defined**., Deborah M. Siegal, M.D., M.Sc.^10,50^, Delcio G. Silva Junior, M.Med.^118,119^, Manu Shankar-Hari, Ph.D.^120,121^, John P. Sheehan, M.D.^122^, Aneesh B. Singhal, M.D.^113,114^, Dayna Solvason^19^, Simon J. Stanworth, F.R.C.P., D.Phil.^43,58^, Tobias Tritschler, M.D., M.Sc.^123^, Anne M. Turner, M.P.H.^34^, Wilma van Bentum-Puijk, M.Sc.^47^, Frank L. van de Veerdonk, M.D., Ph.D.^100^, Sean van Diepen, M.D., M.Sc.^124^, Gloria Vazquez-Grande, M.D., M.Sc.^19^, Lana Wahid, M.D.^125^, Vanessa Wareham, H.B.Sc.^46^, Bryan J. Wells, M.D.^126^, R. Jay Widmer, M.D., Ph.D.^127^, Jennifer G. Wilson, M.D.^128^, Eugene Yuriditsky, M.D.^53^, Fernando G. Zampieri, M.D., Ph.D.^129^.

1 Peter Munk Cardiac Centre at University Health Network, Toronto, Canada

2 University of Toronto, Toronto, Canada

3 University Health Network, Toronto, Canada

4 NYU Grossman School of Medicine, New York City, United States

5 University of Pittsburgh, Pittsburgh, United States

6 UPMC, Pittsburgh, United States

7 Instituto do Coração (InCor), Hospital das ClÍnicas HCFMUSP, Universidade de São Paulo, São Paulo, Brazil

8 Montefiore Medical Center, Bronx, United States

9 Albert Einstein College of Medicine, Bronx, United States

10 Ottawa Hospital Research Institute, Ottawa, Canada

11 Institut du Savoir Montfort, Ottawa, Canada

12 Icahn School of Medicine at Mount Sinai, New York City, United States

13 Mount Sinai Heart, New York City, United States

14 Université Laval, Québec City, Canada

15 CHU de Québec – Université Laval Research Center, Québec City, Canada

16 Instituto Mexicano del Seguro Social, Mexico City, Mexico

17 University of Bristol, Bristol, United Kingdom

18 University Hospitals Bristol and Weston NHS Foundation Trust, Bristol, United Kingdom

19 University of Manitoba, Winnipeg, Canada

20 CancerCare Manitoba, Winnipeg, Canada

21 Zuckerberg San Francisco General Hospital/University of California, San Francisco, United States

22 McGill University, Montreal, Canada

23 Larner College of Medicine at the University of Vermont, Burlington, United States

24 Australian and New Zealand Intensive Care Research Centre, Monash University, Melbourne, Australia

25 St. Michael’s Hospital Unity Health, Toronto, Canada

26 University of Illinois, Chicago, United States

27 Imperial College London, London, United Kingdom

28 Imperial College Healthcare NHS Trust, St. Mary’s Hospital, London, United Kingdom

29 SOCAR Research SA, Nyon, Switzerland

30 London School of Hygiene and Tropical Medicine, London, UK

31 Berry Consultants, LLC, Austin, United States

32 Harbor-UCLA Medical Center, Torrance, United States

33 Auckland City Hospital, Auckland, New Zealand

34 Medical Research Institute of New Zealand, Wellington, New Zealand

35 St John of God Hospital, Subiaco, Australia

36 Vanderbilt University Medical Center, Nashville, United States

37 Fédération Hospitalo Universitaire, Raymond Poincaré Hospital, UVSQ, Garches, France

38 King Saud bin Abdulaziz University for Health Sciences, Riyadh, Kingdom of Saudi Arabia

39 King Abdullah International Medical Research Center, Riyadh, Kingdom of Saudi Arabia

40 Nepal Mediciti Hospital, Lalitpur, Nepal

41 Nepal Intensive Care Research Foundation, Kathmandu, Nepal

42 Versiti Blood Research Institute, Milwaukee, United States

43 Oxford University, Oxford, United Kingdom

44 NICS-MORU, Colombo, Sri Lanka

45 Flinders University, Bedford Park, Australia

46 Ozmosis Research Inc., Toronto, Ontario

47 University Medical Center Utrecht, Utrecht University, Utrecht, The Netherlands

48 Jena University Hospital, Jena, Germany

49 Global Coalition for Adaptive Research, Los Angeles, United States

50 University of Ottawa, Ottawa, Canada

51 Monash University, Melbourne, Australia

52 Alfred Health, Melbourne, Australia

53 NYU Langone Health, NYU Langone Hospital, New York City, United States

54 Aix-Marseille University, Marseille, France

55 University of California San Diego School of Medicine, San Diego, United States

56 Cleveland Clinic, Cleveland, Ohio

57 Ochsner Medical Center, University of Queensland-Ochsner Clinical School, New Orleans, United States

58 NHS Blood and Transplant, Oxford, United Kingdom

59 Harvard Medical School and Brigham and Women’s Hospital, Boston, United States

60 Sunnybrook Health Sciences Centre, Toronto, Canada

61 University of Alabama, Birmingham, United States

62 Hospital Ramón y Cajal (IRYCIS), Madrid, España

63 The Clinical Research, Investigation, and Systems Modeling of Acute Illness (CRISMA) Center, University of Pittsburgh, Pittsburgh, United States

64 TriStar Centennial Medical Center, Nashville, United States

65 University of Antwerp, Wilrijk, Belgium

66 Rutgers New Jersey Medical School, Newark, United States

67 McMaster University, Hamilton, Canada

68 Thrombosis and Atherosclerosis Research Institute, Hamilton, Canada

69 Boston University School of Medicine, Boston, United States

70 Boston Medical Center, Boston, United States

71 University of Oxford, Bangkok, Thailand

72 University College London Hospital, London, United Kingdom

73 Ascension St. John Heart and Vascular Center, Tulsa, United States

74 University of Oklahoma College of Medicine, Oklahoma City, United States

75 Brigham and Women’s Hospital, Boston, United States

76 NYU Langone Long Island, Mineola, United States

77 UPMC Children’s Hospital of Pittsburgh, Pittsburgh, United States

78 University of Cincinnati, Cincinnati, United States

79 Kings Healthcare Partners, London, United Kingdom

80 University of Michigan, Ann Arbor, United States

81 Mayo Clinic, Rochester, United States

82 Apollo Speciality Hospital -OMR, Chennai, India

83 Bellevue Hospital, New York City, United States

84 Oregon Health & Science University, Portland, United States

85 National Heart Lung & Blood Institute, NIH, Bethesda, United States

86 Case Western Reserve University, The Metro Health Medical Centre, Cleveland, United States

87 University of Mississippi Medical Center, Jackson, United States

88 Université de Sherbrooke, Sherbrooke, Canada

89 University of California Los Angeles, Los Angeles, United States

90 Fiona Stanley Hospital, Perth, Australia

91 IdiPaz Research Institute, Universidad Autonoma, Madrid, Spain

92 Cook County Health, Chicago, United States

93 Rush Medical College, Chicago, United States

94 University Hospital of Salamanca-University of Salamanca-IBSAL, Salamanca, Spain

95 Avanti Pesquisa ClÍnica, Sao Paulo, Brazil

96 St Boniface Hospital, Winnipeg, Canada

97 Queen’s University Belfast, Belfast, Northern Ireland

98 Royal Victoria Hospital, Belfast, Northern Ireland

99 Auckland City Hospital, Auckland, New Zealand

100 Radboud University Medical Center, Nijmegen, The Netherlands

101 Penn State Hershey Medical Center, Emergency Medicine, Hershey, United States of America

102 Intensive Care National Audit & Research Centre (ICNARC), London, United Kingdom

103 University of British Columbia, Vancouver, Canada

104 Beaumont Health, Royal Oak, United States

105 OUWB School of Medicine, Auburn Hills, United States

106 University College Dublin, Dublin, Ireland

107 University of Southwestern Medical Center, Dallas, United States

108 The University of Auckland, Auckland, New Zealand

109 University of Chicago, Chicago, United States

110 Hospital do Coração de Mato Grosso do Sul (HCMS), Campo Grande, Brazil

111 Federal University of Mato Grosso do Sul (UFMS), FAMED, Campo Grande, Brazil

112 Hennepin County Medical Center, Minneapolis, United States

113 Massachusetts General Hospital, Boston, United States

114 Harvard Medical School, Boston, United States

115 Hospital 9 de Julho, São Paulo, Brazil

116 INGOH, Clinical Research Center, Goiânia, Brazil.

117 University of Kansas School of Medicine, Kansas City, United States

118 Hospital Universitário Maria Aparecida Pedrossia, Campo Grande, Brazil

119 Hospital Unimed Campo Grande, Campo Grande, Brazil

120 Guy’s and St. Thomas’ NHS Foundation Trust, London, United Kingdom

121 King’s College London, London, United Kingdom

122 University of Wisconsin School of Medicine and Public Health, Madison, United States

123 Inselspital, Bern University Hospital, University of Bern, Switzerland

124 University of Alberta, Edmonton, Canada

125 Duke University Hospital, Durham, North Carolina

126 Emory University, Atlanta, United States

127 Baylor Scott and White Health, Temple, United States

128 Stanford University School of Medicine, Palo Alto, United States

129 HCor-Hospital do Coração, São Paulo, Brazil

