## Supplementary Appendix AC mpRCT Moderate for "Therapeutic Anticoagulation in Non-Critically Ill Patients with Covid-19"

### Multi-Platform Randomized Controlled Trial (mpRCT) Therapeutic anticoagulation in patients with moderate Covid-19

### Section 1 - mpRCT Investigators and Collaborators

#### 1.1 Data Safety and Monitoring Board Members

##### 1.1.1 ATTACC

Jason Connor (Chair); Victoria Manax (Deputy Chair); Julian Bion; Simon Gates; John Reynolds; James Douketis; Damon Scales

##### 1.1.2 ACTIV-4a

Richard C. Becker (Chair); Gregory del Zoppo; Peter Henke; Richard Holubkov; Kim Kerr, Agnes Lee; Fedor Lurie; Sara K. Vesely

##### 1.1.3 REMAP-CAP

Victoria Manax (Chair); Jason Connor (Deputy Chair); Julian Bion; Simon Gates; John Reynolds; Tom van der Poll

#### 1.2 ATTACC

**Lead Principal Investigators**

Ryan Zarychanski, Patrick Lawler, Ewan Goligher

**International Trial Steering Committee**

Ryan Zarychanski, (Chair) (University of Manitoba, Winnipeg, Canada), Patrick Lawler (University of Toronto, Toronto, Canada), Ewan Goligher (University of Toronto, Toronto, Canada), Robert Rosenson (Mount Sinai New York, New York, USA) U.S. Country Lead, Jose Nicolau (University of Sao Paulo, Sao Paulo, Brazil) Brazil Country Lead, Jorge Escobedo (Unidad de Investigación en Epidemiología Clínica, Mexico City, Mexico) Mexico Country Lead, Michael Farkouh (University of Toronto, Toronto, Canada), Dean Fergusson (Ottawa Hospital Research Institute, Ottawa, Canada), Anand Kumar (University of Manitoba, Canada), Nicole Marten (St Boniface Hospital, Winnipeg, Canada), John Marshall (University of Toronto, Toronto, Canada), Alexis F. Turgeon (Université Laval, Québec, Canada), Charlotte Bradbury (University of Bristol, Bristol, United Kingdom), Marc Carrier (University of Ottawa, Canada), Vlad Dzavik (University of Toronto, Toronto, Canada), Rob Fowler (University of Toronto, Toronto, Canada), Emily Gibson McDonald (McGill University, Montréal, Canada), Peter Gross (McMaster University, Canada), Brett Houston (University of Manitoba, Winnipeg, Canada), Mansoor Hussain (University of Toronto, Toronto, Canada), Susan Kahn (McGill University, Montréal, Canada), Srinivas Murthy (University of British Columbia, Vancouver, Canada), Arthur Slutsky (University of Toronto, Toronto, Canada), Tobias Tritschler (University of Ottawa, Ottawa, Canada). Patient Partners: Margaret Ostrowski and Suzanne Dubois (Canada).

**Global Project Management**

*Ozmosis Research Inc.*: Lindsay Bond, Jackie Amaral, Vanessa Wareham, Karlee Trafford, Mila Khanna
*University of Manitoba:* Nicole Marten, Dayna Solvason, Kerrie Hayes, Lynne Hiebert, Holly Musto
*University Health Network:* Maria Kannu

**Global Data Coordinating Center**

*SOCAR Research:* Bridget-Anne Kirwan, Sophie de Brouwer, David La Framboise

**Regional Project Management**

***Brazil***

*Avanti:* Andrea Martinez, Pedro Ohara, Juliana Bacca, Natalia de Jesus, Sandra Zier, Douglas Assis
*InCor:* Jose Nicolau, Natassja Huemer, Neury Martins, Fabiana Nakajima

***Mexico***
*UNAM:* Jorge Escobedo

**Adjudication Committee**
Brendan Everett, Sean van Diepen, Gregoire Le Gal, Deborah Siegal, Jean-Philippe Galanaud, Sheila Hegde, Yuri Kim, Natalia Rost, Aneesh Singhal

**Statistical Analysis Committee**
Roger Lewis, Michelle Detry, Anna McGlothlin (Chair), Mark Fitzgerald, Christina Saunders, Maria Brooks

**Laboratory Committee**
Peter Gross, Rita Selby, Patrick Lawler

**Site Principal Investigators and Research Coordinators**
***Canada***
*Health Sciences Centre:* Brett Houston, Soumya Alias, Rhonda Silva
*St. Boniface General Hospital:* Vi Dao, Nicole Marten, Maureen Hutmacher, Lisa Rigaux, Quinn Tays, Hessam Kashani
*Grace Hospital*: Glen Drobot, Nicole Marten Maureen, Hutmacher, Nora Choi
*University Health Network:* Ewan Goligher, Richard Dunbar-Yaffe, Mohammad Shafiee, Jenna Wong
*Hamilton Health Sciences:* Peter Gross, Michele Zondag
*Ottawa Hospital:* Lana Castellucci, Penny Philips, Moussa Meteb, Irene Watpool, Rebecca Porteous
*Centre Hospitalier Universitaire de Québec – Université Laval*: Alexis F. Turgeon, David Bellemare, Olivier Costerousse, Ève Cloutier, Rana Daher, Marie-Claude Boulanger, Émilie Couillard-Chénrd, François Lauzier, Charles Francoeur
*Centre Hospitalier de l'Université de Montréal:* Emmanuelle Duceppe, Roberta-Daila Carling, Madeleine Durand
*Jewish General Hospital:* Vicky Tagalakis, Elena Shulikovsky, Sophie Florencio
*McGill University Health Centre:* Emily McDonald, Sarah Elsayed, Kristen Moran
*Institut Universitaire de Cardiologie et de Pneumologie – Université Laval:* Francois Lellouche, Patricia Lizotte
*Regina General Hospital:* Andrea Lavoie, Kendra Townsend
*Victoria General Hospital:* Daniel Ovakim, Deborah Parfett, Fiona Auld
*St. Joseph's Healthcare Hamilton:* Peter Gross, Michele, Zondag
*Hôpital Montfort:* Marc Carrier, Mélodie Carrier
*Centre Hospitalier Universitaire de Sherbrooke:* Francois Lamontagne, Elaine Carbonneau

***United States***
*University of Chicago Medicine:* Jonathan Paul, Cynthia Arevalo, Karli Molignoni
*Ochsner Medical Center*: Mark B. Effron, Sarah Cohen, Hunter McDaniel
*William Beaumont Hospital:* Girish B. Nair, Tammy Osentoski
*St. Louis Veterans Affairs Medical Centre:* Martin Schoen, Kristin Courtright, Kelly Reno
*Maine Medical Centre:* Daniel Meyer, Terilee Gerry
*Vanderbilt University Medical Centre:* Aaron Aday, Emily Shardelow, Michaela Burton
*Henry Ford University:* Scott Kaatz, Stacy Ellsworth
*Emory University Hospital Midtown:* Bryan Wells, Claudia Merlin, Amanda Fieback
*Mayo Clinic:* Vivek Iyer, Matthew Johnson
*Saint Barnabas Medical Center:* Nirav Mistry, Amber Turner

*Cooper University Health Care*: Nitin Puri, Christa Schorr
*Hackensack University Medical Center*: Ronaldo Go, Peter Canino
*Montefiore Health System:* Henny Billett, Ervin Mazniku

***Brazil***
*Instituto do Coração (InCor):* Felipe Gallego Lima, Alexandra Vieira, Renata Maia, Anna Mostachio
*Instituto de Infectologia Emilio Ribas:* Walter Braga, Sunamita Lima
*Hospital 9 de Julho:* Fernanda Santos
*Hospital das Clínicas da Faculdade de Medicina da Universidade de São Paulo*: Rinaldo Siciliano
*Sociedade Beneficente Israelita Hospital Albert Einstein:* Remo Furtado, Fernanda Ferraz Assir, Beatriz Moraes
*Instituto Goiano de Oncologia e Hematologia (INGOH):* Mayler Santos, Luciana Barros
*Instituto de Cardiologia de Santa Catarina:* Artur Herdy, Vera Pereira
*Santa Casa de Votuporanga:* Mauro Hernandes, Renée Amorim, Milena Bandeira
*AngioCor Clínica Médica em Blumenau:* Adrian Kormann, Jádina Spricigo, Sabrina Zimmerman
*Hospital São Vicente de Paulo:* Rogerio Tumelero, Andressa Giordani, Flavia Ghizzoni
*Instituto de Medicina Vascular*: Euler Manenti, Karen Ruschel, Aline Borba
*Instituto de Pesquisa Clínica de Campinas (IPECC):* José Saraiva, Carla Vicente, Marina Silva
*Hospital Agamenon Magalhães: Cardiologia:* Joao Moraes Jr, Shylla Ribeiro, Thais Barros
*Centro de Pesquisas Clínicas Humap - UFMS:* Delcio Silva Jr, Paula Serafin, João Xavier
*Instituto de Cardiologia do Rio Grande do Sul:* Oscar Dutra, Andrea Brum
*Hospital Felicio Rocho:* Ana Procopio, Maria Alves
*Praxis Pesquisa Médica:* Anete Grumach, Lara Bertolini
*Hospital Universitário Pedro Ernesto:* Carmem Porto, Samara Oliveira
*Casa de Saúde Santa Marcelina:* Marcelo Burihan, Monica Santos
*UNIMED Campo Grande:* Delcio Silva Jr, Erica Nery, Paula Serafin
*Instituto de Moléstias Cardio Vasculares de Tatuí:* Wladmir Saporito, Thais Pereira, Bruna Mancini
*Santa Casa de Misericórdia de Itabuna:* Eduardo Kowalski Neto, Bruno Andrade, Jessica Santos
*Clínica de Campo Grande S/A:* Mauricio Pompilio, Rebeca Pompilio
*Parana Medical Research Center:* Sergio Grava, Karen Koga
*Hospital das Clínicas da UFPR:* Miguel Silva, Debora Lemos

***Mexico***
*1 Carlos MacGregor Sánchez Navarro:* Jorge Escobedo, Betriz Villegas
*Hospital de Infectologia del Centro Médico Nacional La Raza:* Eduardo Mateos García, Miguel Ángel Cortés Vázquez, Yessica Sara Pérez González, Paulina Carreño Pérez
*Hospital General regional 2 El Marqués:* Julieta Valenzuela, Juan Anwar Santillán

#### 1.3 ACTIV-4a

**Lead Investigators**

Judith S. Hochman MD, Matthew D. Neal MD, Jeffrey S. Berger MD

**Protocol Development Committee**

Judith S. Hochman MD (Chair), Matthew D. Neal MD (co-Chair), Jeffrey S. Berger MD (co-PI)
Mary Cushman, MD, MSc, Lisa Baumann Kreuziger, MD , Scott Berry, PhD, Michael Farkouh, MD, Michelle N. Gong, MD, Kristin Hudock, MD, MSTR, Keri S. Kim, PharmD, Lucy Z. Kornblith, MD, Patrick R. Lawler, MD, MPH, Eric Leifer, PhD, Bryan J. McVerry, MD, Harmony R. Reynolds, MD, Jennifer G. Wilson, MD

**NYU Grossman School of Medicine/ NYU Langone Health Clinical Coordinating Center and Chairs Office**

Judith Hochman (Chair); Jeffrey Berger (Co-PI), Harmony Reynolds (Co-Investigator), Alexander Bragat, Keith Goldfeld, Erinn Hade, Aira Contreras, Stephanie Mavromichalis, Eduardo Iturrate, Margaret Gilsenan, Anna Naumova, Arlene Roberts

**University of Pittsburgh Data Coordinating Center**

Matthew Neal (Co-Chair), Stephen Wisniewski, Christine Leeper, Derek Angus, Heather Eng, Maria Brooks, Kelsey Linstrum, Christopher Seymour, Timothy Girard, Scott Berry, Stephanie Montgomery, Mary Martinez, Jake Schreiber, Joshua Froess, Zhuxuan Fu, Yongqi Zhong, Ashita Vadlamudi, Frank Sciurba, Alison Morris

**SOCAR Research**

Bridget-Anne Kirwan, Sophie de Brouwer, Emilie Perrin, Caroline Gombault, Sandra Bula, Michael Nelson, Céline Daelemans, Robin Wegmüller, David La Framboise

**National Heart Lung and Blood Institute, National Institutes of Health**

Program: W. Keith Hoots, MD, Andrei Kindzelski, MD, PhD, Traci Mondoro, PhD, Antonello Punturieri, MD, PhD, Gail Weinmann, MD, Eric Leifer, PhD, James F. Troendle, PhD

**RTI International**

Amy S. Kendrick, RN, MSN, Tracy L Nolen, DrPH, Sonia Thomas, DrPH

**Network Coordinating Centers**

*ISCHEMIA/MINOCA-HARP/EPPIC-NET:* Harmony Reynolds (Lead), Aira Contreras, Stephanie Mavromichalis, Margaret Gilsenan, Anna Naumova, Danielle Sin, Elhaji Diene, Ewelina Gwiszcz, Isabelle Hogan, Alair Holden
*PETAL:* Michelle Gong (Lead), Nancy Ringwood, Laura Fitzgerald, John Sharer, Daniel Ceusters, Carolyn Hintlian
*Multi-Net*: Lucy Kornblith (Lead), Brenda Nunez-Garcia, Valerie Uribe, Carolyn Hendrickson, Christen Barua, M. Margaret Knudson, John Park, Ana Gonzalez
*FIBHULP*: Jose Lopez-Sendon (Lead), Paloma Moraga Alapont, Paula Prieto, Victoria Hernandez
*RAPID*: Mary Cushman, Lisa Baumann Kreuziger, Shannon Broaddrick
*IlliNet*: Keri Kim, Sean Quigley
*REMAP-CAP*: Bryan McVerry (Lead), David Huang, Meredith Buxton, Tracey Roberts, Kelsey Linstrum
*StrokeNet*: Hooman Kamel, Pooja Khatri, Jamey Frasure, Amy Silken

**Site Investigators and Research Coordinators**

***Spain***

*Hospital Universitario Ramon Y Cajal*: Jose Luis Lopez-Sendon Moreno, Raquel Morillo Guerrero, Sebastian Garcia Madrona, Almudena Molinera, Otilia Navarro Carrion, Raquel Besse Diaz, Sergio Diz Farina, Fernando Hidalgo Salinas, Paula Gonzalez Ferrandiz, Svetlana Zhilina, Macarena Alpanes Buesa, Andres Gonzalez Garcia

*Hospital Clinico Universitario de Salamanca*: Miguel Marcos Martin, Monica Sanchez, Juan Hernandez, Felipe Alvarez Navid, Moncef Belhassen Garcia, Cristina Carbonell Munoz, Guillermo Hernandez Perez, Amparo Lopez Bernus, Jose Angel Martin Oterino

*Hospital Clínico Universitario de Santiago de Compostela*: Jose Ramon Gonzalez Juanatey, Jose Seijas, Maria Jesus Dominguez Santallas, Antonio Pose Reino, Luis Valdes Cuadrado, Nuria Rodriguez Nunez

***United States***

*NYU Langone Health*: Jeffrey Berger, Norma Keller, Eugene Yuriditsky, Tania Ahuja, James Horowitz, Alexander Hindenburg, Tamta Chkhikvadze, Sam Parnia, Zelda Moran, Maja Fadzan, Julia Levine, Stanley Cobos, Arline Roberts, Lia Mamistvalova, Michela Garabedian, Farzana Ahmed, Gabriela Zapata, Marsha Robinson

*University of Illinois at Chicago Health*: John Quigley, Keri Kim, Jeff Jacobson, Neha Atal

*Montefiore Medical Center*: Michelle Gong, Daniel Ceusters, Omowunmi Amosu, Hiwet Tzehaie, Rahul Nair, Brenda Lopez, Manuel Hache Marliere, Daniel Fein, Obiageli Offor, Michael Kiyatkin, Sweta Chekuri, Benjamin Galen, Aram Hambardzumyan, Aditi Desai, Mahmuda Akhter, Hammad Aleem, Sahil Virdi, Roshni Shah, Aluko Hope, Jen-Ting Chen, Amira Mohamed

*Zuckerberg San Francisco General Hospital*: Lucy Kornblith, John Park, Carolyn Hendrickson, M. Margaret Knudson, Aaron Kornblith, India Shelley, Biniam Ambachew, Brenda Nunez-Garcia

*UPMC Presbyterian*: David Huang, Bryan McVerry, Kelsey Linstrum, Nicole Bensen, Dylan Burbee, Aaron Richardson, Amanda McNamara, Dara Stavor, Menna Abaye, Denise Scholl, Renee Wunderley, Anne Yang, Sher Shah Amin, Emily Berryman, Matthew Gilliam**,** Kim Basile, Giles Clermont, William Garrard, Christopher Horvat, Kyle Kalchthaler, Andrew J. King, Daniel Ricketts, Salim Malakouti, Oscar Marroquin, Edvin Music, Kevin Quinn, Mark Andreae, William Bain, Ian Barbash, Emily Brant, David Barton, Meghan Fitzpatrick, Christopher A Franz, Ghady Haidar, Mahwish Hussain, Georgios D Kitsios, Florian B Mayr, Brian Malley, Erin McCreary, Kaveh Moghbeli, Brian Rosborough, Andrew Schoenling, Faraaz A Shah, Varun U Shetty, Tomeka Suber, Nadine Talia, Alexandra Weissman, Caitlin Schaefer, Michael Muir, Kelly Lynn Urbanek

*Rutgers New Jersey Medical School*: Yonatan Greenstein, Randall (Randy) Teeter, Michael Plump, Olga Kovalenko, Eliana Obando, Yanille Taveras, Brittany Fanka, Nipun Suri, Sunil Patel, Maninderpal Kaur

*University of Cincinnati Medical Center*: Kristin Hudock, Robert Hite, Tammy Roads, Adamsegd Gebremedhen, Simra Kiran, Harshada More

*UC San Diego Hillcrest*: Todd Costantini, Terry Curry, Emmer Trinidad, MaryBeth Tyler, Allison Berndtson, Matthew Allison, Harpreet Bhatia, Julie Denenberg, Brennan Marsh-Armstrong, Ilya Verzhbinsky, Timothy Morris, Timothy Fernandes, Ann Elliott, Amelia Eastman

*Ronald Reagan UCLA Medical Center*: George Lim, Gregory Hendey, Steven Chang, Nida Qadir, Rebecca Beutler, Trisha Agarwal, Julia Vargas, Jason Singer, David Haase, James Murphy, Agatha

Brzezinski, Anna Yap, Dong Han Yao, Claudie Bolduc, Catherine Antonuk, Hannah Spungen, Ashley Vuong

*Stanford University Medical Center*: Jennifer Wilson, Angela Rogers, Joseph Levitt, Rosemary Vojnik, Jonasel Roque, Cynthia Perez

*Oregon Health and Science University*: Akram Khan, Olivia Krol, Kinjal Mistry, Kelly Nguyen, Zhengchun Lu, Milad Karami Jouzestani, Atinderpal Singh, Madeline Mcdougal, Andrew Salar, Simeon Florea, Raya Adi, Chandni Anadkat, Emmanuel Mills, Zachary Zouyed, Rupali Deshmukh, Catherine Hough

*Baylor Scott and White Medical Center-Temple*: Robert Widmer, Wanda Fikes, Elizabeth Kiesle

*University of Michigan*: Robert Hyzy, Pauline Park, Shijing Jia, Jakob McSparron, Bonnie Wang, Sinan Hanna, Kelli McDonough, Amanda Melvin, Kristine Nelson, Norman Olbrich

*Sarah Cannon and HCA Research Institute*: Andrew Goodman, Hallie E. Hank, Drew Quillen, Abdullah Shamsuddin, Logan Michl, Molly Harper, Mariah Phipps, Charita Braker

*Duke University Hospital*: Lana Wahid, Oluwayemisi Mohammed, Stephen Gazda, Jacob Craven, Ryan Jackson, Kira Abuchowski, Rowena Dolor, Thomas Ortel, Maria Manson, Lorraine Vergara, Gloria Pinero, Stephanie Freel

*MetroHealth System:* Vidya Krishnan, Cindy Newman, Pete Leo, Carla Greenwood, Andrew Wright, Edward L. Warren, John Daryl Thornton, Calen Frolkis

*UCSF San Francisco*: Michael Matthay, Kirsten, Kangelaris, Kathleen Liu, Carolyn Calfee, Hanjing Zhuo, Brian Daniel, Kimberly Yee, Alejandra Jauregui, Rajani Ghale, Suzanna Chak, Katherine Wick, Emily Siegel, Chayse Jones, Kimia Ashktorab

*Kansas University Medical Center*: Lewis Satterwhite, Penelope Harris, Kimberly Lovell, Mohamed Mourad, Charles Bengtson, Tahani Atieh, Kyle Brownback, Carolina Aguiar, Megan White, Karisa Deculus, Lawrence Scott, Lindsey English, Stephanie Greer, Sharon Murry, Lisa Woodring, Usman Nazir, Amanda Truong, Nelda Mallett, Shereesa Williams, Heidi Hellwig, Michael Burton

*University of Texas Southwestern Medical Center*: Ambarish Pandey, Christopher Bates, Bienka Lewis, Jesse Tarbutton, Nitin Kondamudi, Ruth Giselle Huet, Xiaohang Xu, Marielle Berger-Nagele, Erika Molina

*Cleveland Clinic Foundation*: Abhijit Duggal, Simon Mucha, Omar Mehkri, Alexander King, Bryan Poynter, Kiran Ashok, Nirosshan Thiruchelvam, Debasis Sahoo, Alice Goyanes, Matthew Siuba, Ravi Sunderkrishnan, Steven Minear, Jaime Hernandez-Montfort, Jinesh Mehta, Carla McWilliams, Chinwe Anekwe, Amy Van, Andrea Calderon, Luz Arazo, Syed Sohaib Nasim, Camila De Carvalho Teixeira, Delmy Zelaya

*Cook County Health*: Saurabh Malhotra, Arlet Nedeltcheva, Katayoun Rezai, Michael Hoffman, Ruben Hernandez Acosta, Juan Sarmiento, Shreeyala Uday

*Ascension St. John Clinical Research Institute*: Nicholas Hanna, Anuj Malik, Stacie Merritt, Julie Davenport, Kathryn Mears, Jane Bryce, Melanie Arnold, Joy Norwood , Cheryl Urias

*University of Mississippi Medical Center*: Matthew Kutcher, James Galbraith, Alan Jones, Utsav Nandi, Vishnu Garla, Rebekah Peacock, Jenna Davis, Emily Grenn, Taylor Shaw, Morgan Moore

*Hennepin County Medical Center*: Matt Prekker, Michael Puskarich, Brian Driver, Jason Baker, Anne Frosch, Adam Kolb, Laura Hubbard, Alex Dunn, Audrey Hendrickson, Ellen Maruggi, Tayne

Andersen, William Miller, Abagail Raiter, Radhika Edpuganti, Quinn Ehlen, Grace Leland, Michael Roth, Tyler Scharber, Walker Tordsen, Mackenzie Reing, Ann Isaksen, Heidi Erickson

*University of Wisconsin Hospital; Meriter Hospital*: John Sheehan, Sarah Stewart, Kraig Kumfer, Rafael Veintimilla, Chris Roginski, Nicole Bonk, Scott Ensminger, Muhammad Shahzeb Munir, Jashan Octain, Ann Sheehy, Alexis Waters, Scott Wilson

*Boston University*: Naomi Hamburg, Erika Teresa Minetti, Karla Damus, Robert Eberhardt, Elizabeth Klings, Rena Zheng, Leili Behrooz, Anna Gao

*Denver Health and Hospital Authority:* Mitchell Cohen, Caitlin Robinson, Andrew Byars, Marcela Fitzpatrik-Wilson, Katherine Ling, Tiffany Bendelow, James Wallace, Ivor Douglas

*University of Alabama*: Sheetal Gandotra, Mark Dransfield, Elizabeth Westfall, Micah Whitson, Donna Harris, Derek Russell, Siddharth Patel

*VA New York Harbor Healthcare System*: Binita Shah, Leandro Maranan, Alana Choy-Shan, Nathaniel Smilowitz, Robert Donnino, Jeffrey Lorin, Mary Keary

*Penn State Health Milton S. Hershey Medical Center:* Steven Moore, Kunal Karamchandani, Pauline Go, Anthony Bonavia, Lonnie Fender, Nancy Campbell, Judie Howrylak, Kevin Gardner, Lisa Fox, Paula Trump, Katie Loffredo, McKenna Snyder, Sharon O`Brien, Lisa Schultz, Shane Kinard

*Washington University School of Medicine, ACCS Research*: Grant Bochicchio, Kelly Bochicchio, Stacey Reese, Ricardo Fonseca, Bryan Sato, Kristen Ferguson, Chris Machica, Jennifer McCarthy, Jose Aldana, Rohit Rasane, Melissa Canas, Hussain Afzal, Tiffany Osborn, Mark Hoofnagle, Jennifer Leonard, Jason Snyder, Douglas Schuerer, Melissa Stewart, Piroska Kopar, Kelly Vallar, Jessica Kramer, Isaiah Turnbull

#### 1.4 REMAP-CAP

<u>**International Trial Steering Committee:**</u>

Farah Al-Beidh, Derek Angus, Djillali Annane, Yaseen Arabi, Abi Beane, Wilma van Bentum-Puijk, Scott Berry, Zahra Bhimani, Marc Bonten, Charlotte Bradbury, Frank Brunkhorst, Meredith Buxton, Allen Cheng, Lennie Derde, Lise Estcourt, Herman Goossens, Anthony Gordon, Cameron Green, Rashan Haniffa, Francois Lamontagne, Patrick Lawler, Kelsey Linstrum, Edward Litton, John Marshall, Colin McArthur, Daniel McAuley, Shay McGuinness, Bryan McVerry, Stephanie Montgomery, Paul Mouncey, Srinivas Murthy, Alistair Nichol, Rachael Parke, Jane Parker, Kathryn Rowan, Marlene Santos, Christopher Seymour, Manu Shankar-Hari, Alexis Turgeon, Anne Turner, Frank van de Veerdonk, Steve Webb (Chair), Ryan Zarychanski

<u>**Regional Management Committees**</u>

***Australia and New Zealand***

Yaseen Arabi, Lewis Campbell, Allen Cheng, Lennie Derde, Andrew Forbes, David Gattas, Cameron Green, Stephane Heritier, Peter Kruger, Edward Litton, Colin McArthur (Deputy Executive Director), Shay McGuinness (Chair), Alistair Nichol, Rachael Parke, Jane Parker, Sandra Peake, Jeffrey Presneill, Ian Seppelt, Tony Trapani, Anne Turner, Steve Webb (Executive Director), Paul Young

***Canadian Regional Management Committee***

Zahra Bhimani, Brian Cuthbertson, Rob Fowler, Francois Lamontagne, John Marshall (Executive Director), Venika Manoharan, Srinivas Murthy (Deputy Executive Director), Marlene Santos, Alexis Turgeon, Ryan Zarychanski

***Critical Care Asia (CCA) Regional Management Committee***

Diptesh Aryal, Abi Beane (Chair), Arjen M Dondrop, Cameron Green, Rashan Haniffa (Executive Director), Madiha Hashmi, Deva Jayakumar, John Marshall, Colin McArthur, Srinivas Murthy, Timo Tolppa, Vanessa Singh, Steve Webb

***European Regional Management Committee***

Farah Al-Beidh, Derek Angus, Djillali Annane, Wilma van Bentum-Puijk, Scott Berry, Marc Bonten (Co-Executive Director), Nicole Brillinger, Frank Brunkhorst, Maurizio Cecconi, Lennie Derde (Co-Executive Director and Chair), Stephan Ermann, Bruno Francois, Herman Goossens, Anthony Gordon, Cameron Green, Sebastiaan Hullegie, Rene Markgraff, Colin McArthur, Paul Mouncey, Alistair Nichol, Mathias Pletz, Pedro Povoa, Gernot Rohde, Kathryn Rowan, Lorraine Parker, Irma Scheepstra-Beukers, Steve Webb

***United States Regional Management Committee***

Brian Alexander, Derek Angus (Executive Director), Kim Basile, Meredith Buxton (Chair), Timothy Girard, Christopher Horvat, David Huang, Kelsey Linstrum, Florian Mayr, Bryan McVerry, Stephanie Montgomery, Christopher Seymour

<u>**Regional Coordinating Centers**</u>

***Australia, CCA region, and Saudi Arabia:*** The Australia and New Zealand Intensive Care Research Centre (ANZIC-RC), Monash University
***Canada:*** St. Michael's Hospital, Unity Health Toronto

***Europe:*** University Medical Center Utrecht (UMCU)
***New Zealand:*** The Medical Research Institute of New Zealand (MRINZ)
***United States:*** Global Coalition for Adaptive Research (GCAR), and University of Pittsburgh Medical Center
**CRIT Care Asia (CCA):** NICS MORU.

**Domain-Specific Working Groups**

***Antibiotic and Macrolide Duration Domain-Specific Working Group***

Richard Beasley, Marc Bonten, Allen Cheng (Chair), Nick Daneman, Lennie Derde, Robert Fowler, David Gattas, Anthony Gordon, Cameron Green, Peter Kruger, Colin McArthur, Steve McGloughlin, Susan Morpeth, Srinivas Murthy, Alistair Nichol, Mathias Pletz, David Paterson, Gernot Rohde, Steve Webb

***Corticosteroid Domain-Specific Working Group***

Derek Angus (Chair), Wilma van Bentum-Puijk, Lennie Derde, Anthony Gordon, Sebastiaan Hullegie, Peter Kruger, Edward Litton, John Marshall, Colin McArthur, Srinivas Murthy, Alistair Nicholla Venkatesh, Steve Webb

***Influenza Antiviral Domain-Specific Working Group***

Derek Angus, Scott Berry, Marc Bonten, Allen Cheng, Lennie Derde, Herman Goossens, Sebastiaan Hullegie, Menno de Jong, John Marshall, Colin McArthur, Srinivas Murthy (Chair), Tim Uyeki, Steve Webb

***COVID-19 Antiviral Domain-Specific Working Group***

Derek Angus, Yaseen Arabi (Chair), Kenneth Baillie, Richard Beasley, Scott Berry, Marc Bonten, Allen Cheng, Menno de Jong, Lennie Derde, Eamon Duffy, Rob Fowler, Herman Goossens, Anthony Gordon, Cameron Green, Thomas Hills, Colin McArthur, Susan Morpeth, Srinivas Murthy, Alistair Nichol, Katrina Orr, Rachael Parke, Jane Parker, Asad Patanwala, Kathryn Rowan, Steve Tong, Tim Uyeki, Frank van de Veerdonk, Steve Webb

**COVID-19 Immune Modulation Domain-Specific Working Group**

Derek Angus, Yaseen Arabi, Kenneth Baillie, Richard Beasley, Scott Berry, Marc Bonten, Frank Brunkhorst, Allen Cheng, Nichola Cooper, Olaf Cremer, Menno de Jong, Lennie Derde (Chair), Eamon Duffy, James Galea, Herman Goossens, Anthony Gordon, Cameron Green, Thomas Hills, Andrew King, Helen Leavis, John Marshall, Florian Mayr, Colin McArthur, Bryan McVerry, Susan Morpeth, Srinivas Murthy, Mihai Netea, Alistair Nichol, Kayode Ogungbenro, Katrina Orr, Jane Parker, Asad Patawala, Ville Pettilä (Deputy Chair), Emma Rademaker, Kathryn Rowan, Manoj Saxena, Christopher Seymour, Wendy Sligl, Steven Tong, Tim Uyeki, Frank van de Veerdonk, Steve Webb, Taryn Youngstein

**COVID-19 Immune Modulation -2 Domain-Specific Working Group**

Derek Angus, Scott Berry, Lennie Derde, Cameron Green, David Huang, Florian Mayr, Bryan McVerry, Stephanie Montgomery, Christopher W. Seymour (Chair), Steve Webb

***Therapeutic Anticoagulation Domain-Specific Working Group***

Derek Angustesh Aryal, Scott Berry, Shailesh Bihari, Charlotte Bradbury, Marc Carrier, Dean Fergusson, Robert Fowler, Ewan Goligher (Deputy Chair), Anthony Gordon, Christopher Horvat, David Huang, Beverley Hunt, Devachandran Jayakumar, Anand Kumar, Mike Laffan, Patrick Lawler, Sylvain Lother,

Colin McArthur, Bryan McVerry, John Marshall, Saskia Middeldorp, Zoe McQuilten, Matthew Neal, Alistair Nichol, Christopher Seymour, Roger Schutgens, Simon Stanworth, Alexis Turgeon, Steve Webb, Ryan Zarychanski (Chair)

***Vitamin C Domain-Specific Working Group***

Neill Adhikari (Chair), Derek Angus, Djillali Annane, Matthew Anstey, Yaseen Arabi, Scott Berry, Emily Brant, Angelique de Man, Lennie Derde, Anthony Gordon, Cameron Green, David Huang, Francois Lamonagne (Chair), Edward Litton, John Marshall, Marie-Helene Masse, Colin McArthur, Shay McGuinness, Paul Mouncey, Srinivas Murthy, Rachael Parke, Alistair Nichol, Tony Trapani, Andrew Udy, Steve Webb

***COVID-19 Immunoglobulin Domain-Specific Working Group***

Derek Angus, Donald Arnold, Phillipe Begin, Scott Berry, Richard Charlewood, Michael Chasse, Mark Coyne, Jamie Cooper, James Daly, Lise Estcourt (Chair, UK lead), Dean Fergusson, Anthony Gordon, Iain Gosbell, Heli Harvala-Simmonds, Tom Hills (New Zealand lead), Christopher Horvat, David Huang, Sheila MacLennan, John Marshall, Colin McArthur (New Zealand lead), Bryan McVerry (USA lead), David Menon, Susan Morpeth, Paul Mouncey, Srinivas Murthy, John McDyer, Zoe McQuilten (Australia lead), Alistair Nichol (Ireland lead), Nicole Pridee, David Roberts, Kathryn Rowan, Christopher Seymour, Manu Shankar-Hari (UK lead), Helen Thomas, Alan Tinmouth, Darrell Triulzi, Alexis Turgeon (Canada lead), Tim Walsh, Steve Webb, Erica Wood, Ryan Zarychanski (Canada lead)

***Simvastatin Domain-Specific Working Group***

Derek Angus, Yaseen Arabi, Abi Beane, Carolyn Calfee, Anthony Gordon, Cameron Green, Rashan Haniffa, Deva Jayakumar, Peter Kruger, Patrick Lawler, Edward Litton, Colin McArthur, Daniel McAuley (Chair), Bryan McVerry, Matthew Neal, Alistair Nichol, Cecilia O'Kane, Murali Shyamsundar, Pratik Sinha, Taylor Thompson, Steve Webb, Ian Young

***Antiplatelet Domain-Specific Working Group***

Derek Angus, Scott Berry, Shailesh Bihari, Charlotte Bradbury (Chair), Marc Carrier, Timothy Girard, Ewan Goligher, Anthony Gordon, Ghady Haidar, Christopher Horvat, David Huang, Beverley Hunt, Anand Kumar, Patrick Lawler, Patrick Lawless, Colin McArthur, Bryan McVerry, John Marshall, Zoe McQuilten, Matthew Neal, Alistair Nichol, Christopher Seymour, Simon Stanworth, Steve Webb, Alexandra Weissman, Ryan Zarychanski

***Mechanical Ventilation Domain***

Derek Angus, Wilma van Bentum-Puijk, Lewis Campbell, Lennie Derde, Niall Ferguson, Timothy Girard, Ewan Goligher, Anthony Gordon, Cameron Green, Carol Hodgson, Peter Kruger, John Laffey, Edward Litton, John Marshall, Colin McArthur, Daniel McAuley, Shay McGuinness, Alistair Nichol (Chair) Neil Orford, Kathryn Rowan, Ary Neto, Steve Webb

***ACE-2 RAAS Domain***

Rebecca Baron, Lennie Derde, Slava Epelman, Claudia Frankfurter, David Gattas, Frank Gommans, Anthony Gordon, Rashan Haniffa, David Huang, Edy Kim, Francois Lamontagne, Patrick Lawler (Chair), David Leaf, John Marshall, Colin McArthur, Bryan McVerry, Daniel McAuley, Muthiah Vaduganathan, Roland van Kimmenade, Frank van de Veerdonk, Steve Webb

**Statistical Analysis Committee**
Michelle Detry, Mark Fitzgerald, Roger Lewis (Chair), Anna McGlothlin, Ashish Sanil, Christina Saunders

**Statistical Design Team**
Lindsay Berry, Scott Berry, Elizabeth Lorenzi

**Data Coordinating Team**
Adrian Buzgau, Cameron Green, Alisa Higgins

**Project Management**
***Australia and Saudi Arabia:*** Jane Parker, Vanessa Singh, Claire Zammit
***Canada:*** Zahra Bhimani, Marlene Santos
***CCA:*** Abi Beane, Rashan Haniffa, Timo Tolppa
***Europe:*** Wilma van Bentum-Puijk, Lorraine Parker, Irma Scheepstra-Beukers, Erika Groeneveld, Svenja Peters, Clementina Okundaye, Denise van Hout, Albertine Smit, Linda Rikkert, Sara Bari, Kik Raymakers, Marion Kwakkenbos-Craanen, Sophie Post, Gerwin Schreuder.
***Germany:*** Nicole Brillinger, Rene Markgraf
***Global:*** Cameron Green
***Ireland:*** Kate Ainscough, Kathy Brickell, Peter Doran
***New Zealand:*** Anne Turner
***United Kingdom:*** Farah Al-Beidh, Aisha Anjum, Janis-Best Lane, Elizabeth Fagbodun, Lorna Miller, Paul Mouncey, Karen Parry-Billings, Sam Peters, Alvin Richards-Belle, Michelle Saull, Stefan Sprinckmoller, Daisy Wiley
***United States of America:*** Kim Basile, Meredith Buxton, Kelsey Linstrum, Stephanie Montgomery, Renee Wunderley

**Database Providers**
***Research Online:*** Marloes van Beurden, Evelien Effelaar, Joost Schotsman,
***Spinnaker Software:*** Craig Boyd, Cain Harland, Audrey Shearer, Jess Wren
***University of Pittsburgh Medical Center:*** Giles Clermont, William Garrard, Christopher Horvat, Kyle Kalchthaler, Andrew King, Daniel Ricketts, Salim Malakouti, Oscar Marroquin, Edvin Music, Kevin Quinn
***NICS MORU: on behalf of CCA*****:** Udara Attanayaka, Abi Beane, Sri Darshana, Rashan Haniffa, Pramodya Ishani, Issrah Jawad, Upulee Pabasara, Timo Tolppa, Ishara Udayanga.

**Clinical Trials Groups**
The REMAP-CAP platform is supported by the Australian and New Zealand Intensive Care Society Clinical Trials Group, the Canadian Critical Care Clinical Trials Group, the Irish Critical Care Clinical Trials Network, the UK Critical Care Research Group and the International Forum of Acute Care Trialists.

REMAP-CAP was supported in the UK by the NIHR Clinical Research Network and we acknowledge the contribution of Kate Gilmour, Karen Pearson, Chris Siewerski, Sally-Anne Hurford, Emma Marsh, Debbie Campbell, Penny Williams, Kim Shirley, Meg Logan, Jane Hanson, Becky Dilley, Louise Phillips, Anne Oliver, Mihaela Sutu, Sheenagh Murphy, Latha Aravindan, Joanne Collins, Holly Monaghan, Adam Unsworth, Seonaid Beddows, Laura Ann Dawson, Sarah Dyas, Adeeba Asghar, Kate Donaldson, Tabitha

Skinner, Nhlanhla Mguni, Natasha Muzengi, Ji Luo, Joanna O'Reilly, Chris Levett, Alison Potter, David Porter, Teresa Lockett, Jazz Bartholomew, Clare Rook, Rebecca McKay, Hannah Williams, Alistair Hall, Hilary Campbell, Holly Speight, Sandra Halden, Susan Harrison, Mobeena Naz, Kaatje Lomme, Paula Sharratt, Johnathan Sheffield, William Van't Hoff, James D Williamson, Alex Barnard, Catherine Birch, Morwenna Brend, Emma Chambers, Sarah Crawshaw, Chelsea Drake, Hayley Duckles-Leech, Justin Graham, Heather Harper, Stephen Lock, Nicola McMillan, Cliodhna O'Flaherty, Eleanor OKell, Amber Hayes, Sally Sam, Heather Slade, Susan Walker, Karen Wilding, Jayne Goodwin, Helen Hodgson, Yvette Ellis, Dawn Williamson, Madeleine Bayne, Shane Jackson, Rahim Byrne, Sonia McKenna, Alsion Clinton, NIHR Urgent Public Health Group: https://www.nihr.ac.uk/documents/urgent-public-health-group-members/24638#Members

REMAP-CAP was supported in France by the CRICS-TRIGGERSEP network.

REMAP-CAP was supported in Ireland by the Irish Critical Care Clinical Trials Network and we acknowledge the contribution of Kate Ainscough, Kathy Brickell and Peter Doran.

REMAP-CAP was supported in the Netherlands by the Research Collaboration Critical Care the Netherlands (RCC-Net).

REMAP-CAP was supported in Canada but the Canadian Institutes of Health Research, St. Michael's Unity Health, and the Canadian Critical Care Trials Group

REMAP-CAP was supported in the United States by the Translational Breast Cancer Research Consortium, the UPMC Learning While Doing Program, and the Global Coalition for Adaptive Research

**<u>Site Investigators and Research Coordinators</u>**

***Australia:***

*The Alfred Hospital*: Andrew Udy, Phoebe McCracken, Meredith Young, Jasmin Board, Emma Martin
*Ballarat Health Services*: Khaled El-Khawas, Angus Richardson, Dianne Hill, Robert J Commons, Hussam Abdelkharim
*Bendigo Hospital*: Cameron Knott, Julie Smith, Catherine Boschert
*Caboolture Hospital*: Julia Affleck, Yogesh Apte, Umesh Subbanna, Roland Bartholdy, Thuy Frakking
*Campbelltown Hospital*: Karuna Keat, Deepak Bhonagiri, Ritesh Sanghavi, Jodie Nema, Megan Ford
*Canberra Hospital*: Harshel G. Parikh, Bronwyn Avard, Mary Nourse
*Concord Repatriation General Hospital*: Winston Cheung, Mark Kol, Helen Wong, Asim Shah, Atul Wagh
*Eastern Health (Box Hill, Maroondah & Angliss Hospitals)*: Joanna Simpson, Graeme Duke, Peter Chan, Brittney Carter, Stephanie Hunter
*Flinders Medical Centre*: Shailesh Bihari, Russell D Laver, Tapaswi Shrestha, Xia Jin
*Fiona Stanley Hospital*: Edward Litton, Adrian Regli, Susan Pellicano, Annamaria Palermo, Ege Eroglu
*Footscray Hospital*: Craig French, Samantha Bates, Miriam Towns, Yang Yang, Forbes McGain
*Gold Coast University Hospital*: James McCullough, Mandy Tallott
*John Hunter Hospital*: Nikhil Kumar, Rakshit Panwar, Gail Brinkerhoff, Cassandra Koppen, Federica Cazzola
*Launceston General Hospital*: Matthew Brain, Sarah Mineall
*Lyell McEwin Hospital*: Roy Fischer, Vishwanath Biradar, Natalie Soar

*Logan Hospital*: Hayden White, Kristen Estensen, Lynette Morrison, Joanne Sutton, Melanie Cooper
*Monash Health (Monash Medical Centre, Dandenong Hospital & Casey Hospital):* Yahya Shehabi, Wisam Al-Bassam, Amanda Hulley, Umesh Kadam, Kushaharan Sathianathan
*Nepean Hospital*: Ian Seppelt, Christina Whitehead, Julie Lowrey, Rebecca Gresham, Kristy Masters
*Princess Alexandra Hospital*: Peter Kruger, James Walsham, Mr Jason Meyer, Meg Harward, Ellen Venz
*The Prince Charles Hospital*: Kara Brady, Cassandra Vale, Kiran Shekar, Jayshree Lavana, Dinesh Parmar
*The Queen Elizabeth Hospital*: Sandra Peake, Patricia Williams, Catherine Kurenda
*Rockhampton Hospital*: Helen Miles, Antony Attokaran
*Royal Adelaide Hospital*: Samuel Gluck, Stephanie O'Connor, Marianne Chapman, Kathleen Glasby
*Royal Darwin Hospital*: Lewis Campbell, Kirsty Smyth, Margaret Phillips
*Royal Melbourne Hospital*: Jeffrey Presneill, Deborah Barge, Kathleen Byrne, Alana Driscoll, Louise Fortune
*Royal North Shore Hospital*: Pierre Janin, Elizabeth Yarad, Frances Bass, Naomi Hammond, Anne O'Connor
*Royal Perth Hospital*: Sharon Waterson, Steve Webb, Robert McNamara
*Royal Prince Alfred Hospital*: David Gattas, Heidi Buhr, Jennifer Coles
*Sir Charles Gardiner Hospital*: Sacha Schweikert, Bradley Wibrow, Matthew Anstey, Rashmi Rauniyar
*St George Hospital*: Kush Deshpande, Pam Konecny, Jennene Miller, Adeline Kintono, Raymond Tung
*St. John of God Midland Public and Private Hospitals*: Ed Fysh, Ashlish Dawda, Bhaumik Mevavala
*St. John of God Hospital, Murdoch*: Annamaria Palermo, Adrian Reglirt De Keulenaer
*St. John of God Hospital, Subiaco*: Ed Litton, Janet Ferrier
*St. Vincent's Hospital (NSW)*: Priya Nair, Hergen Buscher, Claire Reynolds, Sally Newman
*St. Vincent's Hospital (VIC)*: John Santamaria, Leanne Barbazza, Jennifer Homes, Roger Smith
*Sunshine Coast University Hospital*: Peter Garrett, Lauren Murray, Jane Brailsford, Loretta Forbes, Teena Maguire
*Sunshine Hospital*: Craig French, Gerard Fennessy, John Mulder, Rebecca Morgan, Rebecca McEldrew
*The Sutherland Hospital*: Anas Naeem, M, Laura Fagan, Emily Ryan,
*Toowoomba Hospital*: Vasanth Mariappa, Judith Smith
*University Hospital Geelong*: Scott Simpson, Matthew Maiden, Allison Bone, Michelle Horton, Tania Salerno
*Wollongong Hospital*: Martin Sterba, Wenli Geng

***Belgium:***
*Ghent University Hospital*: Pieter Depuydt, Jan De Waele, Liesbet De Bus, Jan Fierens, Stephanie Bracke, Joris Vermassen, Daisy Vermeiren

***Canada*:**
*Brantford General Hospital:* Brenda Reeve, William Dechert
*Centre de recherche de l'Institut universitaire de cardiologie et de pneumologie de Québec*: Francois Lellouche, Patricia Lizotte
*Centre Hospitalier de l'Universite de Montreal:* Michaël Chassé, François Martin Carrier, Dounia Boumahni, Fatna Benettaib, Ali Ghamraoui
*CHU de Québec – Université Laval*: Alexis Turgeon, David Bellemare, Ève Cloutier, Rana Daher, Olivier Costerousse, Marie-Claude Boulanger, Émilie Couillard-Chénard, François Lauzier Charles Francoeur

*Centre Hospitalier Universitaire de Sherbrooke:* François Lamontagne, Frédérick D'Aragon Elaine Carbonneau, Julie Leblond
*Grace Hospital:* Gloria Vazquez-Grande, Nicole Marten
*Grand River Hospital (Kitchener)*: Theresa Liu, Atif Siddiqui
*Health Sciences Centre, Winnipeg:* Ryan Zarychanski, Gloria Vazquez-Grande, Nicole Marten, Maggie Wilson
*Hôpital du Sacré Coeur de Montréal:* Martin Albert, Karim Serri, Alexandros Cavayas, Mathilde Duplaix, Virginie Williams
*Juravinski Hospital:* Bram Rochwerg, Tim Karachi, Simon Oczkowski, John Centofanti, Tina Millen
*McGill University Health Centre*: Kosar Khwaja, Josie Campisi
*Niagara Health (St. Catherine's Hospital):* Erick Duan, Jennifer Tsang, Lisa Patterson
*Regina General Hospital*: Eric Sy, Chiraag Gupta, Sandy SHA Kassir
*Royal Alexandra Hospital*: Demetrios Kutsogiannis, Patricia Thompson
*Sunnybrook Health Sciences Centre*: Rob Fowler, Neill Adhikari, Maneesha Kamra, Nicole Marinoff
*St. Boniface General Hospital*: Ryan Zarychanski, Nicole Marten
*St. Joseph's Healthcare Hamilton*: Deborah Cook, Frances Clarke
*St. Mary's General Hospital (Kitchener)*: Rebecca Kruisselbrink, Atif Siddiqui
*St. Michael's Hospital*: John Marshall, Laurent Brochard, Karen Burns, Gyan Sandhu, Imrana Khalid
*The Ottawa Hospital:* Shane English, Irene Watpool, Rebecca Porteous, Sydney Miezitis, Lauralyn McIntyre
*University Health Network:* Elizabeth Wilcox, Lorenzo del Sorbo, Hesham Abdelhady, Tina Romagnuolo
*University of Alberta:* Wendy Sligl, Nadia Baig, Oleksa Rewa, Sean Bagshaw
*William Osler Health System:* Alexandra Binnie, Elizabeth Powell, Alexandra McMillan, Tracy Luk, Noah Aref

**Critical Care Asia:**
*India Apollo Speciality Hospital - OMR, Chennai*: Devachandran Jayakumar, R Pratheema, Suresh Babu
*Apollo Main Hospital, Chennai*: C Vignesh, Bharath Kumar TV, N Ramakrishnan, Augustian James, Evangeline Elvira

*Apollo Speciality Vanagaram, Vanagaram, Chennai*; R Ebenezer, S Krishnaoorthy, Lakshmi Ranganathan, Manisha, Madhu Shree
*Apollo First Med Hospital, Chennai:* Ashwin Kumar Mani, Meghena Mathew, Revathi

*Nepal Grande International Hospital*: Sushil Khanal, Sameena Amatya
*HAMS Hospital*: Hem Raj Paneru, Sabin Koirala, Pratibha Paudel
*Nepal Mediciti Hospital*: Diptesh Aryal, Kanchan Koirala, Namrata Rai, Subekshya Luitel
*Tribhuvan University Teaching Hospital:* Hem Raj Paneru, Binita Bhattarai
*Pakistan Ziauddin Hospital Clifton Campus:* Prof Madiha Hashmi, Ashok Panjwani, Zulfiqar Ali Umrani, Shoaib Siddiq, Mohiuddin Shaikh
*National Institute of Cardiovascular Diseases Pakistan*: Nawal Salahuddin, Sobia Masood

***Croatia:***
*General Hospital Pozega*: Zdravko Andric, Sabina Cviljevic, Renata Đimoti, Marija Zapalac, Gordan *Mirković*

*University Hospital of Infectious Diseases "Fran Milhajevid"*: Bruno Baršić, Marko Kutleša, Viktor Kotarski
*University Hospital of Zagreb*: Ana Vujaklija Brajković, Jakša Babel, Helena Sever, Lidija Dragija, Ira Kušan

***Finland:***
*Helsinki University Hospital*: Suvi Vaara, Leena Pettilä, Jonna Heinonen, Ville Pettilä
*Tampere University Hospital*: Anne Kuitunen, Sari Karlsson, Annukka Vahtera, Heikki Kiiski, Sanna Ristimäki

***France:***
*Ambroise Pare Hospital*: Amine Azaiz, Cyril Charron, Mathieu Godement, Guillaume Geri, Antoine Vieillard-Baron
*Centre Hospitalier de Melun*: Franck Pourcine, Mehran Monchi
*Centre Hospitalier Simone Veil, Beauvais*: David Luis, Romain Mercier, Anne Sagnier, Nathalie Verrier, Cecile Caplin, Jack Richecoeu, Daniele Combaux
*Centre Hospitalier Sud Essonne*: Shidasp Siami, Christelle Aparicio, Sarah Vautier, Asma Jeblaoui, Delphine Lemaire-Brunel
*Centre Hospitalier Tenon*: Muriel Fartoukh, Laura Courtin, Vincent Labbe, Guillaume Voiriot, Sara Nesrine Salhi
*Centre Hospitalier Victor Dupouy*: Gaetan Plantefeve, Cécile Leparco, Damien Contou
*CHR d'Orleans*: Grégoire Muller, Mai-Anh Nay, Toufik Kamel, Dalila Benzekri, Sophie Jacquier, Isabelle Runge, Armelle Mathonnet, François Barbier, Anne Bretagnol
*CHRU Tours Hopital Bretonneau*: Emmanuelle Mercier, Delphine Chartier, Charlotte Salmon, Pierre-François Dequin, Denis Garot
*Hôpital de Hautepierre, Hôpitaux Universitaires de Strasbourg*: Francis Schneider, Vincent Castelain, Guillaume Morel, Sylvie L'Hotellier
*Hospital Nord Franche-Comté*: Julio Badie, Fernando Daniel Berdaguer, Sylvain Malfroy, Chaouki Mezher, Charlotte Bourgoin, Guy Moneger, Elodie Bouvier
*Lariboisière Hospital*: Bruno Megarbane, Sebastian Voicu, Nicolas Deye, Isabelle Malissin, Laetitia Sutterlin, Aymen Mrad, Adrien Pépin Lehalleur, Giulia Naim, Philippe Nguyen, Jean-Michel Ekhérian, Yvonnick Boué, Georgios Sidéris, Dominique Vodovar, Emmanuelle Guérin, Caroline Grant
*Le Mans Hospital*: Christophe Guitton, Cédric Darreau, Mickaël Landais, Nicolas Chudeau, Alain Robert, Patrice Tirot, Jean Christophe Callahan, Marjorie Saint Martin, Charlène Le Moal, Rémy Marnai, Marie Hélène Leroyer
*Raymond Poincaré Hospital*: Djillali Annane, Pierre Moine, Nicholas Heming, Virginie Maxime, Isabelle Bossard, Tiphaine Barbarin Nicholier, Bernard Clair, David Orlikowski, Rania Bounab, Lilia Abdeladim
*Vendee Hospital*: Gwenhael Colin, Vanessa Zinzoni, Natacha Maquigneau, Matthieu Henri-Lagarrigue, Caroline Pouplet

***Germany:***
*Carl-Thiem-Klinikum Cottbus gGmbH*: Jens Soukup, Richard Wetzold, Madlen Löbel, Dr. Ing, Lisa Starke, Patrick Grimm
*Charité - Universitätsmedizin Berlin*: André Finn, Gabriele Kreß, Uwe Hoff, Carl Friedrich Hinrichs, Jens Nee

*Jena University Hospital*: Mathias W. Pletz, Stefan Hagel, Juliane Ankert, Steffi Kolanos, Frank Bloos
*Klinikum Dortmund gGmbH*: Daniela Nickoleit-Bitzenberger, Bernhard Schaaf, Werner Meermeier, Katharina Prebeg, Harun Said Azzaui, Martin Hower, Klaus-Gerd Brieger, Corinna Elender, Timo Sabelhaus, Ansgar Riepe, Ceren Akamp, Julius Kremling, Daniela Klein, Elke Landsiedel-Mechenbier;
*University Hospital of Leipzig*: Sirak Petros, Kevin Kunz, Bianka Schütze
*Universitätsklinikum Hamburg-Eppendorf*: Stefan Kluge, Axel Nierhaus, Dominik Jarczak, Kevin Roedl
*University Hospital of Frankfurt*: Gernot Gerhard Ulrich Rohde, Achim Grünewaldt, Jörg Bojunga
*University Hospital of Wuerzburg*: Dirk Weismann, Anna Frey, Maria Drayss, M.E. Goebeler, Thomas Flor, Gertrud Fragner, Nadine Wahl, Juliane Totzke, Cyrus Sayehli
*Vivantes Klinikum Neukölln*: Lorenz Reill, Michael Distler, Astrid Maselli

***Hungary:***
*Almási Balogh Pál Hospital, Ózd*: János Bélteczki, István Magyar, Ágnes Fazekas, Sándor Kovács, Viktória Szőke;
*Jósa András County Hospital, Nyíregyháza*: Gábor Szigligeti, János Leszkoven;

***Ireland:***
*Beacon Hospital Dublin*: Daniel Collins, Kathy Brickell, Liadain Reid, Michelle Smyth, Patrick Breen, Sandra Spain
*Beaumont Hospital*: Gerard Curley, Natalie McEvoy, Pierce Geoghegan, Jennifer Clarke
*Galway University Hospitals*: John Laffeyirbre McNicholas, Michael Scully, Siobhan Casey, Maeve Kernan, Aoife Brennan, Ritika Rangan, Riona Tully, Sarah Corbett, Aine McCarthy, Oscar Duffy, David Burke
*St Vincent's University Hospital, Dublin*: Alistair Nichol, Kathy Brickell, Michelle Smyth, Leanne Hayes, Liadain Reid, Lorna Murphy, Andy Neill, Bryan Reidy, Michael O'Dwyer, Donal Ryan, Kate Ainscough

***Netherlands:***
*Canisius Wilhelmina Ziekenhuis*: Oscar Hoiting, Marco Peters, Els Rengers, Mirjam Evers, Anton Prinssen
*Deventer Hospital*: Huub L.A. van den Oever, Arriette Kruisdijk-Gerritsen
*Jeroen Bosch Ziekenhuis*: Koen Simons, Tamara van Zuylen, Angela Bouman
*Meander Medisch Centrum*: Laura van Gulik
*Radboud University Medical Center Nijmegen*: Jeroen Schouten, Peter Pickkers, Noortje Roovers, Margreet Klop-Riehl, Hetty van der Eng
*UMC Leiden*: Evert de Jonge, Jeanette Wigbers, Michael del Prado
*UMC Utrecht*: Marc Bonten, Olaf Cremer, Lennie Derde, Jelle Haitsma Mulier, Anna Linda Peters, Birgit Romberg, Helen Leavis, Roger Schutgens
*Ziekenhuis Gelderse Vallei*: Sjoerd van Bree, Marianne Bouw-Ruiterrbara Festen, Fiona van Gelder, Mark van Iperen, Margreet Osinga, Roel Schellaars, Dave Tjan, Ruben van der Wekken, Max Melchers, Arthur van Zanten
*Haga Ziekenhuis*: Kees van Nieuwkoop, Thomas Ottens, Yorik Visser
*Onze Lieve Vrouwe Gasthuis*: Nicole Juffermans, Matty Koopmans

***New Zealand*:**

*Auckland City Hospital, Cardiothoracic and Vascular ICU:* Shay McGuinness, Rachael Parke, Eileen Guilder, Magdalena Butler, Keri-Anne Cowdrey, Melissa Woollett
*Auckland City Hospital, DCCM:* Colin McArthur, Thomas Hills, Lynette Newby, Yan Chen, Catherine Simmonds, Rachael McConnochie, Caroline O'Connor
*Christchurch Hospital:* Jay Ritzema Carter, Seton Henderson, Kymbalee Van Der Heyden, Jan Mehrtens, Anna Morris, Stacey Morgan
*Middlemore Hospital:* Tony Williams, Alex Kazemi, Susan Morpeth, Rima Song, Vivian Lai, Dinuraj Girijadevi
*North Shore Hospital:* Robert Everitt, Robert Russell, Danielle Hackin
*Rotorua Hospital:* Ulrike Buehner, Erin Williams
*Tauranga Hospital:* Troy Browne, Kate Grimwade, Jennifer Goodson, Owen Keet, Owen Callender
*Waikato Hospital:* Robert Martynoga, Kara Trask, Amelia Butler
*Wellington Hospital:* Paul Young, Chelsea Young, Eden Lesona, Shaanti Olatunji, MClinIm, Leanlove Navarra, Raulle Sol Cruz
*Whangarei Hospital:* Katherine Perry, Ralph Fuchs, Bridget Lambert
*Taranaki Base Hospital:* Jonathan Albrett, Carolyn Jackson, Simon Kirkham

***Portugal*:**
*Hospital de Abrantes:* Nuno José Teodoro Amaro dos Santos Catorze, Tiago Nuno Alfaro Lima Pereira, Ricardo Manuel Castro Ferreira, Joana Margarida Pereira Sousa Bastos, Teresa Margarida Oliveira Batista

***Romania*:**
*"Dr. Victor Babes" Clinical Hospital of Infectious and Tropical Diseases Bucharest:* Simin Aysel Florescu, Delia Stanciu, Mihaela Florentina Zaharia, Alma Gabriela Kosa, Daniel Codreanu

***Saudi Arabia*:**
*King Abdulaziz Medical City- Riyadh:* Yaseen M Arabi, Eman Al Qasim, Haytham Tlayjeh, Lolowa Alswaidan, Brintha Naidu

***Spain*:**
*Hospital del Mar:* Rosana Muñoz-Bermúdez, Judith Marin-Corral, Anna Salazar Degracia, Francisco Parrilla Gómez, Maria Isabel Mateo López
*Reina Sofia University Hospital:* Rafael León López, Jorge Rodriguez, Sheila Cárcel, Rosario Carmona, Carmen de la Fuente, Marina Rodriguez

***United Kingdom*:**
*Aberdeen Royal Infirmary:* Callum Kaye, Angela Allan;
*Addenbrooke's Hospital:* Charlotte Summers, Petra Polgarova;
*Alder Hey Children's NHS Foundation Trust:* Stephen J McWilliam, Daniel B Hawcutt, Laura Rad, Laura O'Malley, Jennifer Whitbread;
*Alexandra Hospital Redditch:* Olivia Kelsall, Nicholas Cowley, Laura Wild, Jessica Thrush, Hannah Wood, Karen Austin;

*Altnagelvin Hospital:* Adrian Donnelly, Martin Kelly, Naoise Smyth, Sinéad O'Kane, Declan McClintock, Majella Warnock, Ryan Campbell, Edmund McCallion;

*Antrim Area Hospital:* Paul Johnson, Shirley McKenna, Joanne Hanley, Andrew Currierbara Allen, Clare Mc Goldrick, Moyra Mc Master;

*Barnet Hospital:* Rajeev Jha, Michael Kalogirou, Christine Ellis, Vinodh Krishnamurthy, Vashish Deelchand, Aibhilin O'Connor;

*Basildon Universty Hospital: Dipak Mukherjee, Agilan Kaliappan, Anirudda Pai, Mark Vertue, Anne Nicholson, Joanne Riches, Gracie Maloney, Lauren Kittridge, Amanda Solesbury, Kezia Allen*

*Belfast Health and Social Care Trust (Belfast City Hospital, Mater Infirmorium, Royal Victoria Hospital):* Jon Silversides, Peter McGuigan, Kathryn Ward, Aisling O'Neill, Stephanie Finn ;

*Brighton and Sussex University Hospitals Trust:* Barbara Phillips, Laura Oritz-Ruiz de Gordoa ;

*Bristol Royal Infirmary:* Jeremy Bewley, Matthew Thomas, Katie Sweet, Lisa Grimmer, Rebekah Johnson;

*Calderdale and Huddersfield Foundation Trust:* Jez Pinnell, Matt Robinson, Lisa Gledhill, Tracy Wood;

*Cardiff and Vale University Health Board:* Matt Morgan, Jade Cole, Helen Hill, Michelle Davies, Angharad Williams, Emma Thomas, Rhys Davies, Matt Wise;

*Charing Cross Hospital:* David Antcliffe, Maie Templeton, Roceld Rojo, Phoebe Coghlan, Joanna Smee ;

*Chesterfield Royal Hospital:* Euan Mackay, Jon Cort, Amanda Whileman, Thomas Spencer, Nick Spittle, Sarah Beavis, Anand Padmakumar, Katie Dale, Joanne Hawes, Emma Moakes, Rachel Gascoyne, Kelly Pritchard, Lesley Stevenson, Justin Cooke, Karolina Nemeth-Roszpopa;

*The Christie NHS Foundation Trust:* Vidya Kasipandian, Amit Patel, Suzanne Allibone, Roman Mary-Genetu ;

*Colchester Hospital:* Mohamed Ramali, Alison Ghosh, Rawlings Osagie, Malka Jayasinghe Arachchige, Melissa Hartley;

*Countess of Chester Hospital:* Peter Bamford, Emily London, Kathryn Cawley, Maria Faulkner, Helen Jeffrey;

*Croydon University Hospital:* Ashok Sundar Raj, Georgios Tsinaslanidis, Reena Nair Khade, Gloria Nwajei Agha, Rose Nalumansi Sekiwala;

*Cumberland Infirmary:* Tim Smith, Chris Brewer, Jane Gregory;

*Darlington Memorial Hospital:* James Limb, Amanda Cowton, Julie O'Brien, Kelly Postlethwaite;

*Derriford Hospital:* Nikitas Nikitas, Colin Wells, Liana Lankester, Helen McMillan;

*Dorset County Hospital:* Mark Pulletz, Patricia Williams, Jenny Birch, Sophie Wiseman, Sarah Horton;

*East Kent Hospitals (Queen Elizabeth the Queen Mother Hospital):* Ana Alegria, Salah Turki, Tarek Elsefi, Nikki Crisp, Louise Allen;

*East Lancashire Hospitals NHS Trust (Royal Blackburn Hospital):* Matthew Smith, Sri Chukkambotla, Wendy Goddard, Stephen Duberley;

*Freeman Hospital and Royal Victoria Infirmary, Newcastle upon Tyne*: Iain J McCullagh, Philip Robinson, Bijal Patel, Sinead Kelly;

*Frimley Health NHS Foundation Trust:* Omar Touma, Susan Holland, Christopher Hodge, Holly Taylor, Meera Alderman, Nicky Barnes, Joana Da Rocha, Catherine Smith, Nicole Brooks, Thanuja Weerasinghe, Julie-Ann Sinclair, Yousuf Abusamra, Ronan Doherty, Joanna Cudlipp, Rajeev Singh, Haili Yu, Admad Daebis, Christopher NG, Sara Kendrick, Anita Saran, Ahmed Makky, Danni Greener, Louise Rowe-Leete, Alexandra Edwards, Yvonne Bland, Rozzie Dolman, Tracy Foster;

*Gateshead Health NHS Trust:* Vanessa Linnett, Amanda Sanderson, Jenny Ritzema, Helen Wild;

*George Eliot Hospital:* Divya Khare, Meredith Pinder, Selvin Selvamoni, Amitha Gopinath;

*Glan Clwyd Hospital:* Richard Pugh, Daniel Menzies, Richard Lean, Xinyi Qiu, Jeremy James Scanlon;

*Glasgow Royal Infirmary:* Kathryn Puxty, Susanne Cathcart, Chris Mc Govern, Samantha Carmichael, MRPharms, Dominic Rimmer;
*Glenfield Hospital Leicester:* Hakeem Yusuff, Graziella Isgro, Chris Brightling, Michelle Bourne, Michelle Craner, Rebecca Boyles;
*Grange University Hospital:* Tamas Szakmany, Shiney Cherian, Gemma Williams, Christie James, Abby Waters;
*Great Western Hospitals NHS Foundation Trust:* Malcolm Watters, Rachel Prout, Louisa Davies, Suzannah Pegler, Lynsey Kyeremeh, Aiman Mian;
*Guy's & St Thomas' NHS Foundation Trust:* Manu Shankar-Hari, Marlies Ostermann, Marina Marotti, Neus Grau Novellas, Aneta Bociek;
*Hammersmith Hospital:* Stephen Brett, Sonia Sousa Arias, Rebecca Elin Hall;
*Homerton University Hospital NHS Foundation Trust:* Susan Jain, Abhinav Gupta, Catherine Holbrook
*James Cook University Hospital:* Jeremy Henning, Stephen Bonner, Keith Hugill, Emanuel Cirstea, Dean Wilkinson, Jessica Jones;
*James Paget University Hospitals:* Michal Karlikowski, Helen Sutherland, Elva Wilhelmsen, Jane Woods, Julie North;
*Kettering General Hospital:* Dhinesh Sundaran, Laszlo Hollos, Susan Coburn, Anna Williams, Samantha Saunders;
*King's College Hospital (Denmark Hill site):* Phil Hopkins, John Smith, Harriet Noble, Maria Theresa Depante, Emma Clarey;
*Lancashire Teaching Hospitals NHS Foundation Trust:* Shondipon Laha, Mark Verlander, Alexandra Williams;
*Leeds Teaching Hospitals Trust:* Elankumaran Paramasivam, Elizabeth Wilby, Bethan Ogg, Clare Howcroft, Angelique Aspinwall, Sam Charlton, Richard Gould, Deena Mistry, Sidra Awan, Caroline Bedford;
*Leicester General Hospital:* Andrew Hall, Jill Cooke, Caroline Gardiner-Hill, Carolyn Maloney, Nigel Brunskill;
*Leicester Royal Infirmary:* Hafiz R QureshiI, Neil Flint, Sarah Nicholson, Sara Southin, Andrew Nicholson, Amardeep Ghattaoraya;
*Lewisham and Greenwich NHS Trust:* Daniel Harding, Sinead O'Halloran, Amy Collins Emma Smith, Estefania Trues;
*Liverpool Foundation Trust Aintree:* Barbara Borgatta, Ian Turner-Bone, Amie Reddy, Laura Wilding;
*Liverpool Heart and Chest Hospital:* Craig Wilson, Zuhra Surti;
*Luton and Dunstable University Hospital:* Loku Chamara Warnapura, Ronan Agno, Prasannakumari Sathianathan, Deborah Shaw, Nazia Ijaz, Dean Burns, Mohammed Nisar, Vanessa Quick, Craig Alexander, Sanil Patel, Nafisa Hussain, Yvynne Croucher, Eva-Maria Langnu Rudran, Syed Gilani, Talia Wieder, Margaret Louise Tate;
*Maidstone and Tunbridge Wells NHS Trust:* David Golden, Miriam Davey, Rebecca Seaman;
*Manchester Royal Infirmary:* Tim Felton, Jonathan Bannard-Smith, Joanne Henry, Richard Clark, Kathrine Birchall, Joanne Henry, Fiona Pomeroy, Rachael Quayle, Katharine Wylie, Anila Sukuraman, John McNamarra;
*Medway Maritime Hospital:* Arystarch Makowski, Beata Misztal, Iram Ahmed, Kevin Neicker, Sam Millington, Rebecca Squires, Masroor Phulpoto;
*Milton Keynes University Hospital:* Richard Stewart, Esther Mwaura, Louise E Mew, Lynn Wren, Felicity Willams;

*Mid & South Essex NHS Foundation Trust:* Aneta Oborska, Rino Maeda, Selver Kalchko-Veyssal, Raji Orat Prabakaran, Bernard Hadebe, Eric Makmur, Guy Nicholls;

*Musgrove Park Hospital:* Richard Innes, Patricia Doble, Libby Graham, Charmaine Shovelton;

*Nevill Hall Hospital:* Vincent Hamlyn, Nancy Hawkins, Anna Roynon-Reed, Sean Cutler, Sarah Lewis;

*Newham University Hospital:* Juan Martin Lazaro, Tabitha Newman;

*Ninewells Hospital:* Pauline Austin, Susan Chapman, Louise Cabrelli;

*Norfolk and Norwich University Hospital:* Simon Fletcher, Jurgens Nortje, Deirdre Fottrell-Gould, Georgina Randell, Katie Stammers;

*Northampton General Hospital:* Mohsin Zaman, Einas Elmahi, Andrea Jones, Kathryn Hall;

*Northern General Hospital, Sheffield:* Gary H Mills, Kim Ryalls, RegPharmTech, Kate Harrington RCN, Helen Bowler, Jas Sall, Richard Bourne;

*North Manchester General Hospital:* Zoe Borrill, Tracy Duncan, Thomas Lamb, Joanne Shaw, Claire Fox, Kirstie Smith, Sarah Holland, Bethany Blackledge, Liam McMorrow, Laura Durrans, Jade Harris;

*North Middlesex University Hospital:* Jeronimo Moreno Cuesta, Kugan Xavier, Dharam Purohit, Munzir Elhassan, Anne Haldeos, Rachel Vincent, Marwa Abdelrazik, Samuel Jenkins, Arunkumar Ganesan, Rohit Kumar, David Carter, Dhanalakshmi Bakthavatsalam;

*Oxford University Hospitals:* Matthew Rowland, Paula Hutton, Archana Bashyal, Neil Davidson, Clare Hird, Sally Beer;

*Pilgrim Hospital Boston:* Manish Chhablani, Gunjan Phalod, Amy Kirkby, Simon Archer, Kimberley Netherton;

*Princess Royal Hospital:* Barbara Philips, Dee Mullan, Denise Skinner, Jane Gaylard, Julie Newman;

*Princess of Wales Hospital:* Sonia Arun Sathe, Lisa Roche, Ellie Davies, Keri Turner;

*Poole Hospital:* Henrik Reschreiter, Julie Camsooksai, PGDE, Sarah Patch, Sarah Jenkins, Charlotte Humphrey;

*Queen Alexandra Hospital Portsmouth:* David Pogson, Steve Rose, Zoe Daly, Lutece Brimfield, Angie Nown;

*Queen Elizabeth Hospital, Birmingham:* Dhruv Parekh, Colin Bergin, Michelle Bates, Christopher McGhee, Daniella Lynch, Khushpreet Bhandal, Kyriaki Tsakiridou, Amy Bamford, Lauren Cooper, Tony Whitehouse, Tonny Veenith;

*Queen Elizabeth University Hospital, Glasgow:* Malcolm AB Sim, Sophie Kennedy Hay, Steven Henderson, MPH, Maria Nygren, Eliza Valentine;

*Queen's Hospital, Burton:* Amro Katary, Gill Bell, Louise Wilcox, Katy English, Ann Adams;

*Queen's Hospital, Romford:* Mandeep-Kaur Phull, Abbas Zaidi, Tatiana Pogreban, Lace Paulyn Rosaroso;

*Queens Medical Centre and Nottingham City Hospital:* Daniel Harvey, Benjamin Lowe, Megan Meredith, Lucy Ryan, DREEAM Research Team;

*The Rotherham NHS Foundation Trust:* Anil Hormis, Rachel Walker, Dawn Collier, Sarah Kimpton, Susan Oakley;

*Royal Alexandra Hospital:* Kevin Rooney, Natalie Rodden, Emma Hughes, Nicola Thomson, Deborah McGlynn, Charlotte Clark, Patricia Clark;

*Royal Berkshire Hospital:* Andrew Walden, Liza Keating, Matthew Frise, Tolu Okeke, Nicola Jacques, Holly Coles, Emma Tilney, Emma Vowell;

*Royal Bournemouth and Christchurch Hospitals:* Martin Schuster-Bruce, Sally Pitts, Rebecca Miln, Laura Purandare, Luke Vamplew;

*Royal Brompton Hospital:* Brijesh Patel, Debra Dempster, Mahitha Gummadi, Natalie Dormand, Shu Fang Wang;

*Royal Cornwall NHS Trust:* Michael Spivey, Sarah Bean, Karen Burt, Lorraine Moore;

*Royal Devon and Exeter NHS Foundation Trust:* Christopher Day, Charly Gibson, Elizabeth Gordon, Letizia Zitter, Samantha Keenan;

*Royal Glamorgan Hospital:* Jayaprakash Singh, Ceri Lynch, Lisa Roche, Justyna Mikusek, Bethan Deacon, Keri Turner;

*Royal Gwent Hospital*: Tamas Szakmany, Evelyn Baker, John Hickey, Shreekant Champanerkar, Lindianne Aitken, Lorraine Lewis Prosser;

*Royal Hallamshire Hospital, Sheffield:* Gary H Mills, Ajay Raithatha, Kris Bauchmuller, Norfaizan Ahmad, Matt Wiles, Jayne Willson;

*Royal Hampshire Hospitals:* Irina Grecu, Jane Martin, Caroline Wrey Brown, Ana-Marie Arias, Emily Bevan;

*Royal Infirmary of Edinburgh:* Thomas H Craven, David Hope, Jo Singleton, Sarah Clark, Corrienne McCulloch;

*Royal Liverpool University Hospital:* Ingeborg D Welters, David Oliver Hamilton, Karen Williams, Victoria Waugh, David Shaw, Suleman Mulla, Alicia Waite, Jaime Fernandez Roman, Maria Lopez Martinez;

*Royal London Hospital:* Zudin Puthucheary, Timothy Martin, Filipa Santos, Ruzena Uddin, Maria Fernandez, Fatima Seidu, Alastair Somerville, Mari Lis Pakats, Priya Dias, Salam Begum, Tasnin Shahid;

*The Royal Free Hospital:* Sanjay Bhagani, Mark De Neef, Helder Filipe, Sara Mingos, Amitaa Maharajh, Glykeria Pakou, Aarti Nandani;

*The Royal Marsden NHS Foundation Trust:* Kate Colette Tatham, Shaman Jhanji, Ethel Blackurs, Arnold Dela Rosaurs, Ryan Howle, Ravishankar Rao Baikady;

*The Royal Oldham Hospital:* Redmond P Tully, Andrew Drummond, Joy Dearden, Jennifer E Philbin, Sheila Munt;

*The Royal Wolverhampton NHS Trust:* Shameer Gopal, Jagtar- Singh Pooni, Saibal Ganguly, Andrew Smallwood, Stella Metherell;

*Royal Papworth Hospital:* Alain Vuylsteke, Charles Chan, Saji Victor, COVID Research Team, Papworth Hospital;

*Royal Stoke Hospital:* Ramprasad Matsa, Minerva Gellamucho, Michelle Davies;

*Royal Surrey County Hospital:* Ben Creagh-Brown, Joe Tooley, Laura Montague, Fiona De Beaux, Laetitia Bullman;

*Royal United Hospital Bath:* Ian Kerslake, Carrie Demetriou, Sarah Mitchard, Lidia Ramos, Katie White;

*Russells Hall Hospital:* Michael Reay, Steve Jenkins, Caroline Tuckwell, Angela Watts, Eleanor Traverse, Stacey Jennings;

*Salisbury NHS Foundation Trust:* Phil Donnison, Maggie Johns, Ruth Casey, Lehentha Mattocks, Sarah Salisbury;

*Salford Royal NHS Foundation Trust:* Paul Dark, Alice Harvey, Reece, Doonan, Liam McMorrow, Karen Knowles;*Sandwell and West Birmingham NHS Trust:* Jonathan Hulme, Santhana Kannan, Sibet Joseph, Fiona Kinney, Ho Jan Senya;

*Sherwood Forest Hospitals NHS Foundation Trust:* Valli Ratnam, Mandy Gill, Jill Kirk, Sarah Shelton

*South Tyneside District Hospital:* Christian Frey, Riccardo Scano, Madeleine McKee, Peter Murphy;

*Southmead Hospital:* Matt Thomas, Ruth Worner, Beverley Faulkner, Emma Gendall, Kati Hayes, Hayley Blakemore, Borislava Borislavova;

*St. Bartholomew's Hospital:* Colin Hamilton-Davies, Carmen Chan, Celina Mfuko, Hakam Abbass, Vineela Mandadapu;

*St. George's Hospital:* Susannah Leaver, Kamal Patel, Sarah Farnell-Ward, Romina Pepermans Saluzzio, John Rawlins;

*St. Mary's Hospital:* Anthony Gordon; Dorota Banach, Ziortza Fernández de Pinedo Artaraz, Leilani Cabreros;

*St. Peter's Hospital, Chertsey:* Ian White, Maria Croft, Nicky Holland, Rita Pereira;

*Stepping Hill Hospital, Stockport:* Ahmed Zaki, David Johnson, Matthew Jackson, Hywel Garrard, Vera Juhaz, Louise Brown;

*Sunderland Royal Hospital:* Alistair Roy, Anthony Rostron, Lindsey Woods, Sarah Cornell;

*Swansea Bay University Health Board:* Suresh Pillai, Rachel Harford, Helen Ivatt, Debra Evans, Suzanne Richards, Eilir Roberts, James Bowen James Ainsworth;

*Torbay and South Devon NHS Foundation Trust:* Thomas Clark, Angela Foulds, Stacey Atkins;

*United Lincolnshire NHS Trust:* Kelvin Lee, Russell Barber, Anette Hilldrith, Claire Hewitt, Gunjan Phalod;

*University Hospitals Coventry & Warwickshire NHS Trust:* Pamela Bremmer, Geraldine Ward, Christopher Bassford;

*University Hospital of North Tees:* Farooq Brohi, Vijay Jagannath, Michele Clark, Sarah Purvis, Bill Wetherill;

*University Hospital Southampton NHS Foundation Trust:* Ahilanandan Dushianthan, Rebecca Cusack, Kim de Courcy-Golder, Karen Salmon, Rachel Burnish, Simon Smith, Susan Jackson, Winningtom Ruiz, Zoe Duke, Magaret Johns, Michelle Male, Kirsty Gladas, Satwinder Virdee, Jacqueline Swabe, Helen Tomlinson;

*Warwick Hospital:* Ben Attwood, Penny Parsons, Bridget Campbell, Alex Smith;

*Watford General Hospital:* Valerie J Page, Xiao Bei Zhao, Deepali Oza, Gail Abrahamson, Ben Sheath, Chiara Ellis;

*Western General Hospital, Edinburgh:* Jonathan Rhodes, Thomas Anderson, Sheila Morris;

*Whipps Cross Hospital:* Charlotte Xia Le Tai, Amy Thomas, Alexandra Keen;

*Whiston Hospital*: Ascanio Tridente, Karen Shuker, Jeanette Anders, Sandra Greer, Paula Scott, Amy Millington, Philip Buchanan, Jodie Kirk

*Wirral University Teaching Hospital NHSFT:* Craig Denmade, Girendra Sadera, Reni Jacob, Cathy Jones, Debbie Hughes;

*Worcester Royal Hospital:* Stephen Digby, Nicholas Cowley, Laura Wild, Jessica Thrush, Hannah Wood, Karen Austin;

*Wrexham Maelor Betsi Cadwaladr University Hospital*: David Southern, Harsha Reddy, Sarah Hulse, Andy Campbell, Mark Garton, Claire Watkins, Sara Smuts;

*Wrightington, Wigan and Leigh Teaching Hospitals NHS Foundation Trust:* Alison Quinn, Benjamin Simpson, Catherine McMillan, Cheryl Finch, Claire Hill, Josh Cooper;

*Wye Valley NHS Trust:* Joanna Budd, Charlotte Small, Ryan O'Leary, Janine Birch, Emma Collins;

*Wythenshawe Hospital*: Peter DG Alexander, Tim Felton, Susan Ferguson, Katharine Sellers, Joanne Bradley-Potts;

*York Teaching Hospital*: David Yates, Isobel Birkinshaw, Kay Kell, Zoe Scott, Harriet Pearson;

***United States of America***

*University of Pittsburgh Research Staff*: CRISMA Center—Kelsey Linstrum, Stephanie Montgomery, Kim Basile, Dara Stavor, Dylan Burbee, Amanda McNamara, Renee Wunderley, Nicole Bensen, Aaron

Richardson; MACRO Center—Peter Adams, Tina Vita, Megan Buhay, Denise Scholl, Matthew Gilliam, James Winters, Kaleigh Doherty, Emily Berryman

*UPMC Hospital Champions*: UPMC Altoona—Mehrdad Ghaffari, UPMC East—Meghan Fitzpatrick; UPMC Jameson and Horizon —Kavitha Bagavathy; UPMC Mercy—Mahwish Hussain, Chenell Donadee; UPMC Williamsport—Emily Brant; UPMC McKeesport—Kayla Bryan-Morris, John Arnold and Bob Reynolds; UPMC Hamot—Gregory Beard; UPMC Presbyterian—Bryan McVerry, David Huang, Ghady Haidar, Alexandra Weissman, Florian Mayr; UPMC Passavant – Matthew Gingo; UPMC Pinnacle—Janice Dunsavage, Salim Saiyed, Erik Hernandez, John Goldman, Cynthia Brown, Susan Comp, James Raczek, Jenny Lynne Morris, Jesus Vargas Jr., Daniel Weiss, Joseph W. Hensley, Erik Kochert, Chris Wnuk, Christopher Nemeth, Brent Mowery, Christina Hutchinson, Lauren Winters

*UPMC COVID Therapeutics Committee*: Erin McCreary, Elise Martin, Ryan Bariola, Alex Viehman, Jessica Daley, Alyssa Lopus, Mark Schmidhofer, UPMC Directors of Pharmacy

*UPMC ICU Service Center*: Rachel Sackrowitz, Chenell Donadee, Aimee Skrtich

*UPMC Wolff Center*: Tami Minnier, Mary Kay Wisniewski, Katelyn Mayak

*UPMC eRecord Team*: Richard Ambrosino, Sherbrina Keen, Sue Della Toffalo, Martha Stambaugh, Ken Trimmer, Reno Perri, Sherry Casali, Rebecca Medva, Brent Massar, Ashley Beyerl, Jason Burkey, Sheryl Keeler, Maryalyce Lowery, Lynne Oncea, Jason Daugherty, Chanthou Sevilla, Amy Woelke, Julie Dice, Lisa Weber, Jason Roth, Cindy Ferringer, Deborah Beer, Jessica Fesz, Lillian Carpio

*Data Collection/Curation Team*: Salim Malakouti (Computer Science, University of Pittsburgh), Edvin Music and Dan Ricketts (CRISMA Center), Andrew King (Biomedical Informatics, University of Pittsburgh), Gilles Clermont (Critical Care Medicine), Robert Bart (UPMC Health Services Division)

*UPMC Clinical Analytics*: Oscar Marroquin, Kevin Quinn, William Garrard, Kyle Kalchthaler

*UPMC Office of Healthcare Innovation*: Derek Angus

*Department of Emergency Medicine*: Alexandra Weissman, Donald Yealy, David Barton, Nadine Talia

*Department of Critical Care Medicine*: David Huang, Florian Mayr, Andrew Schoenling, Mark Andreae, Varun Shetty, Emily Brant, Brian Malley, Chenell Donadee, Derek Angus, Christopher Horvat, Christopher Seymour, Timothy Girard, Gilles Clermont, Rachel Sackrowitz, Robert Bart

*Division of Infectious Diseases*: Ghady Haidar

*Division of Pulmonary, Allergy, and Critical Care Medicine*: Bryan McVerry, William Bain, Ian Barbash, Meghan Fitzpatrick, Christopher Franz, Georgios Kitsios, Kaveh Moghbeli, Brian Rosborough, Faraaz Shah, Tomeka Suber

*Berry Consultants*: Roger Lewis, Michelle Detry, Anna McGlothlin, Christina Saunders, Mark Fitzgerald, Ashish Sanil, Scott Berry

*Global Coalition for Adaptive Research (GCAR):* Meredith Buxton, Brian Alexander, Tracey Roberts

#### 1.5 Funding Agencies

##### 1.5.1 ATTACC

The ATTACC platform was supported by grants from the Canadian Institutes of Health Research, LifeArc Foundation, Thistledown Foundation, Research Manitoba, Ontario Ministry of Health, Peter Munk Cardiac Centre, Cancercare Manitoba Foundation, and Victoria General Hospital Foundation.

##### 1.5.2 ACTIV-4a

The ACTIV-4a platform was sponsored by the National Heart, Lung, and Blood Institute, National Institutes of Health, Bethesda and administered through OTA-20-011. The research was, in part, funded by the National Institutes of Health (NIH) Agreement 1OT2HL156812-01.

##### 1.5.3 REMAP-CAP

Supported by the European Union — through FP7-HEALTH-2013-INNOVATION: the Platform for European Preparedness Against (Re-)emerging Epidemics (PREPARE) consortium (602525), and Horizon 2020 research and innovation program: the Rapid European Covid-19 Emergency Research response (RECOVER) consortium (101003589) — and by the Australian National Health and Medical Research Council (APP1101719), the Health Research Council of New Zealand (16/631), the Canadian Institutes of Health Research (Strategy for Patient-Oriented Research Innovative Clinical Trials Program Grant - 158584, and COVID-19 Rapid Research Operating Grant - 447335), the U.K. NIHR and the NIHR Imperial Biomedical Research Centre, the Health Research Board of Ireland (CTN 2014-012), the UPMC Learning While Doing Program, the Breast Cancer Research Foundation, the French Ministry of Health (PHRC-20-0147), the Minderoo Foundation, Amgen, Eisai, the Global Coalition for Adaptive Research, and the Wellcome Trust Innovations Project (215522). Anthony Gordon is funded by an NIHR Research Professorship (RP-2015-06-18), Manu Shankar-Hari by an NIHR Clinician Scientist Fellowship (CS-2016-16-011), Alexis Turgeon is funded by a Canada Research Chair – Tier 2. Ryan Zarychanski is the recipient of the Lyonel G Israels Research Chair in Hematology (University of Manitoba).

##### 1.5.4 Disclaimer

The views expressed in this publication are those of the author(s) and not necessarily those of the National Health Service (UK), the National Institute for Health Research (UK), the Department of Health and Social Care (UK), or of the National Institutes of Health (USA).

### Section 2 – Supplemental Methods

#### Introduction

The multi-platform randomized controlled trial (mpRCT) described herein represents an unprecedented display of global collaboration whereby three organizationally distinct platforms (ATTACC, ACTIV4a, and REMAP-CAP) harmonized protocols and worked together to answer the important question of whether a strategy of empiric therapeutic anticoagulation benefits patients with Covid-19 compared to usual care thromboprophylaxis.

The concept of the mp-RCT was borne out of discussion and shared interest among clinical trialists and leaders of three major international platforms to evaluate the efficacy of therapeutic anticoagulation for inpatients with Covid-19. To answer the question as quickly as possible and with maximal generalizability, three protocols were harmonized to create a single mpRCT with common eligibility criteria, study intervention details, and key primary and secondary outcomes. Data collection, leadership, and oversight are shared across the platforms, and agreement was established to federate the data collected within each platform into one overarching analysis. Trial data providers are SOCAR (ATTACC, ACTIV IV), SPIRAL (REMAP-CAP), and UPMC (REMAP-CAP). Statisticians from Berry Consultants serve as trial statisticians for all three platforms with an independent group comprising the Statistical Analysis Committee (SAC) for the mpRCT. With agreed upon stopping rules for efficacy and futility for the primary endpoint, the mpRCT operates under a unified multiplatform analysis plan with monthly interim analyses (See Protocol Appendix Statistical Analysis Plan – Page 481). The mpRCT investigators collaborate on making all major decisions.

Oversight was executed collaboratively across the three platforms, each with its own independent DSMB. Each trial DSMB maintained its responsibilities within its platform. The interim efficacy and safety analyses were reported to the DSMBs for each of the trials. Since safety data are not aggregated across trials, safety analysis reports for each platform were shared with the DSMBs of other platforms. Upon viewing safety reports, each DMC reached a consensus on safety topics, and the DSMB chairs or designees from each of the platforms communicated with one another to discuss the efficacy and safety results across trials. Platform leadership and DSMB chairs agreed that individual platforms would not make public disclosures of efficacy without the agreement of all three oversight boards. A pre-defined publication plan was established by platform investigators for the mpRCT.

The results presented in the manuscript reflect the highly collaborative effort of the network of networks efficiently working together to rapidly answer a question of high public health importance. In effort to simplify the data review process, this supplement includes protocol synopses of each contributing platform and three tables that outline the harmonization across the three platforms. A separate protocol appendix contains detailed versions of each individual platform protocol with relevant protocol amendments and the unified mpRCT statistical analysis plan.

#### Supplementary Methods

Several sensitivity analyses of the primary outcome of organ support-free days were conducted in the overall moderate cohort. A sensitivity analysis of the primary outcome was repeated in a model nested in the overall trial population (i.e., allowing dynamic borrowing of information on treatment effect in the overall moderate cohort from participants in the severe cohort). The remainder of models detailed below assumed independent treatment effects from severe participants. An additional sensitivity analysis assessed whether results varied with the inclusion of participants who were randomized as suspected Covid-19 but ultimately did not document SARS-CoV-2 PCR test positivity. A sensitivity analysis assessed whether the results were modified by excluding site and time effects from the model. The analytical model includes site and time covariate terms to account for variability in treatment effect according to site (random effect) and to variation in the endpoint over time (fixed effect) – the influence of these effects was tested with removing these covariables from the model. A sensitivity analysis also categorized the primary model into three ordinal categories (survival to hospital discharge without receipt of organ support, survival to hospital discharge with receipt of organ support, or death during index hospitalization irrespective of receptive of organ support). A sensitivity analysis was performed excluding participants who were receiving an antiplatelet agent at enrollment as part of usual care or who were randomized to either treatment or control in the concurrent antiplatelet domain of REMAP-CAP (https://www.remapcap.org/protocol-documents). Finally, to understand the potential influence of possible treatment cross-over with the public announcement of the adaptive stopping results, a sensitivity analysis was performed restricting to participants enrolled on or before January 7, 2021, who would have had up to the maximum 14 days needed to complete the protocolized intervention prior to public announcement of the adaptive analysis results.

Protocol adherence was defined by the anticoagulant dose equivalent administered within the first 24-48 hours following randomization (see **Page 47** below for consensus dosing categories), with doses categorized as therapeutic and subtherapeutic heparin qualifying as adherent in the therapeutic-dose anticoagulation arm, and low- and intermediate prophylactic-dose anticoagulants qualifying as adherent in the usual care thromboprophylaxis arm.

Secondary outcomes were examined: survival free of organ support through 28 days (dichotomous outcome – without censoring at index hospital discharge); survival free of invasive mechanical ventilation through 28 days (ordinal, with death as the worst possible outcome – without censoring at index hospital discharge); mortality through 28 days (time-to-event – without censoring at index hospital discharge); mechanical respiratory support-free days through 28 days (ordinal, calculated as the primary endpoint, except that death is included as 0 – censored at index hospital discharge); survival to hospital discharge free of major thrombotic events (dichotomous; including a composite of freedom from myocardial infarction, pulmonary embolism, ischemic stroke, systemic arterial embolism, and in-hospital death); survival to hospital discharge free of any macrovascular thrombotic event (including the components of major thrombotic events, as well as deep venous thrombosis); hospital length of stay (time-to-event; beginning at enrollment); and freedom from major bleeding (as defined by the International Society on Thrombosis and Haemostasis – reported during the treatment window). All thrombotic and bleeding events were adjudicated blindly using consensus definitions centrally. For all outcomes, posterior probabilities and median proportional adjusted odds ratios or hazard ratios are reported. **Table S4** also reports crude proportions; importantly, response-adaptive randomization may lead to imbalances in baseline covariates between treatment arms over time, and as such the Bayesian models of treatment effect are necessarily adjusted for age, sex, site, D-dimer group, and time, and as such, the primary results reflect adjusted treatment effects. Models examining secondary outcomes do not use statistical borrowing, except for in-hospital mortality in the D-dimer-defined groups, which derives from the primary outcome. There was no imputation of missing outcomes in either primary or secondary analyses. Cases were excluded on an analysis-by-analysis basis; i.e., patients missing outcome data were included in treatment compliance and safety analyses. The secondary outcomes were also analysed in the unblinded modified intention-to-treat population. The primary safety outcomes were assessed during the treatment window (which extended up to 14 days, or until recovery – defined as hospital discharge or liberation from supplemental oxygen for >24 hours) across groups. Additional secondary outcomes were specified in the core statistical analysis plan but were not included in the sub-statistical analysis plan for this report. These additional outcomes will be presented subsequently, including when more detailed long term outcome data are available.

##### Model Equation: Cumulative Logistic Regression

The equation for the cumulative logistic regression is given by the following:

$$\log\left(\frac{\pi_{isy}}{1-\pi_{isy}}\right) = \alpha_{y,s} - \left[\nu_{Site,s} + \lambda_{Time,s} + \theta_{a,s:d} + \beta_{Age,s} + \beta_{Sex,s} + \beta_{d}\right]$$

Where site = site (random effect), time = 2 week epochs of time (fixed effect), a,s:d = arm within sub-type given d-dimer level, age (categorical variable), sex = sex at birth, d = d-dimer. See statistical analysis plan (Protocol Appendix page 452) for more details.

##### Response-Adaptive Randomization

REMAP-CAP and ATTACC specified the possibility for response-adaptive randomization (RAR). The blinded RAR proportions for ATTACC were based on the mpRCT interim results and varied by D-dimer groups, with the missing D-dimer group maintaining equal randomization. RAR proportions for the Moderate, Low and High D-dimer groups were based on the posterior probability of the odds-ratio for therapeutic anticoagulation being greater than 1 within each subtype. The randomization probabilities were truncated between 0.10 and 0.90. Thus, based on the first mpRCT interim analysis, the randomization probability for therapeutic anticoagulation within ATTACC was changed on December 15, 2020, to 90% in the Low D-dimer group and to 83% in the High D-dimer group. These proportions remained until enrollment was closed on January 22, 2021. REMAP-CAP specified the possibility for, but did not employ, RAR in the moderate state as the statistical trigger was met for the mpRCT prior to implementation of RAR in the moderate state. ACTIV-4a did not specify the possibility for RAR.

#### Protocol Synopses

ATTACC

| | |
|---|---|
| Study Title: | **A**nti**T**hrombotic **T**herapy to **A**meliorate **C**omplications of **C**OVID-19 (**ATTACC**) in collaboration with Accelerating COVID-19 Therapeutic Interventions and Vaccines (ACTIV-4) |
| Study Design: | A phase III prospective, open-label, adaptive multi- platform randomized controlled trial |
| Primary Objective/ Endpoint: | The primary endpoint in the trial is days alive and free of organ support at day 21. This endpoint is defined as the number of days that a patient is alive and free of organ support through the first 21 days after trial entry. |
| Secondary Objectives: | Secondary Safety Endpoints:<br>- Laboratory confirmed heparin induced thrombocytopenia **(HIT)**<br>- **Major bleeding**, defined according to the International Society on Thrombosis and Haemostasis (ISTH)/Scientific and Standardization Committee (SSC) definitions and bleeding assessment tool in non-surgical patients (Schulman *J Thromb Haemost* 2005): fatal bleeding; and/or symptomatic bleeding in a critical area or organ, such as intracranial, intraspinal, intraocular, retroperitoneal, intraarticular or pericardial, or intramuscular with compartment syndrome; and/or bleeding causing a fall in hemoglobin level of ≥20 g/L, or leading to transfusion of 2 or more units of whole blood or red cells.<br><br>Secondary Efficacy Endpoints:<br>- A composite endpoint of death, deep vein thrombosis, pulmonary embolism, systemic arterial thromboembolism, myocardial infarction, or ischemic stroke collected during hospitalization or at 28 days and 90 days after enrollment (whichever is earlier)<br>- Ordered categorical endpoint with three possible outcomes based on the worst status of each patient through day 30 following randomization: no invasive mechanical ventilation, invasive mechanical ventilation, or death. |

| | |
|---|---|
| | - All cause mortality assessed at 28 and 90 days following randomization<br>- All cause mortality during initial hospitalization (includes death after 28 days)<br>- Intubation assessed at 30 days following randomization<br>- Ventilator-free days (days alive not on a ventilator) assessed at 28 days following randomization<br>- Hospital-free days (days alive outside hospital assessed at 28 days following randomization)<br>- Vasopressor-free days (days alive not on a vasopressor) assessed at 28 days following randomization<br>- Renal replacement free days (days alive not on renal replacement) assessed at 28 days following randomization<br>- Hospital re-admission within 28 days<br>- Symptomatic proximal venous thromboembolism (DVT or PE) assessed at 28 and 90 days following randomization<br>- Myocardial infarction assessed at 28 and 90 days following randomization<br>- Ischaemic stroke assessed at 28 and 90 days following randomization<br>- Acute kidney injury as defined by KDIGO criteria<br>- Systemic arterial thrombosis or embolism assessed at 28 and 90 days following randomization<br>- Use of extracorporeal membrane oxygenation (ECMO) support<br>- Mechanical circuit (dialysis or ECMO) thrombosis<br>- WHO ordinal scale (peak scale over 28 days, scale at 14 days, and proportion with improvement by at least 2 categories compared to enrollment, at 28 days) |
| Duration: | The duration of accrual on this study will be ongoing in nature during the COVID-19 pandemic, following outcomes for each patient up to a maximum of 90 days. |

| | |
|---|---|
| Planned Total Sample Size: | The trial is a Bayesian adaptive design and as such is not predicated on a fixed *a priori* sample size. This design was chosen given uncertainty regarding anticipated event rates and potential treatment effect sizes. Approximately 350 to a maximum of 3000 evaluable patients are anticipated to be enrolled in this adaptive trial in combination with the ACTIV 4 and REMAP-CAP trials, with anticipated reprioritization of key subgroups (including D-dimer defined) as the trial is undertaken. |
| Drug Administration: | Participants randomized to the <u>investigational arm</u> will receive therapeutic anticoagulation for 14 days (or until hospital discharge or liberation from the need for supplemental oxygen, whichever comes first) with preference for low-molecular weight heparin (LMWH), or alternative unfractionated heparin (UFH).<br><br>Participants randomized to the <u>control arm</u> will receive usual care, which is anticipated to include thromboprophylactic dose anticoagulation according to local practice. |
| Inclusion/Exclusion Criteria: | Inclusions:<br>1. Patients ≥18 years of age providing (possibly through a substitute decision maker) informed consent who require hospitalization anticipated to last ≥72 hours, for microbiologically confirmed COVID-19, enrolled < 72 hours of hospital admission **or** of COVID-19 confirmation<br><br>Exclusions:<br>1. Requirement for chronic mechanical ventilation via tracheostomy prior to hospitalization<br>2. Patients for whom the intent is to not use pharmacologic thromboprophylaxis<br>3. Active bleeding<br>4. Risk factors for bleeding, including:<br>a. intracranial surgery or stroke within 3 months;<br>b. history of intracerebral arteriovenous malformation;<br>c. cerebral aneurysm or mass lesions of the central nervous system; |

| | |
|---|---|
| | d. intracranial malignancy<br>e. history of intracranial bleeding<br>f. history of bleeding diatheses (e.g., hemophilia)<br>g. history of gastrointestinal bleeding within previous 3 months<br>h. thrombolysis within the previous 7 days<br>i. presence of an epidural or spinal catheter<br>j. recent major surgery <14 days<br>k. uncontrolled hypertension (sBP >200 mmHg, dBP >120 mmHg)<br>l. other physician-perceived contraindications to anticoagulation<br>5. Platelet count <50 x$10^9$/L, INR >2.0, or baseline aPTT >50<br>6. Hemoglobin <80 g/L (to minimize the likelihood of requiring red blood cell transfusion if potential bleeding were to occur)<br>7. Acute or subacute bacterial endocarditis<br>8. History of heparin induced thrombocytopenia (HIT) or other heparin allergy including hypersensitivity<br>9. Current use of dual antiplatelet therapy<br>10. Patients with an independent indication for therapeutic anticoagulation<br>11. Patients in whom imminent demise is anticipated and there is no commitment to active ongoing intervention<br>12. Anticipated transfer to another hospital that is not a study site within 72 hours<br>13. Enrollment in other trials related to anticoagulation or antiplatelet therapy |
| Study Assessments: | Study assessments are depicted in the study schedule. |
| Safety Variables & Analysis: | The safety of therapeutic anticoagulation with LWMH or intravenous UFH infusion will be evaluated by AE reports. Treatment-related AEs include bleeding and HIT. |
| Efficacy Assessments & Analysis | The efficacy of therapeutic-dose parenteral anticoagulation with subcutaneous LMWH or |

| | |
|---|---|
| | intravenous UFH will be evaluated in comparison to usual care. |
| Reasons for premature discontinuation of therapy: | Treatment will continue until any of the following occurs:<br>• Heparin induced thrombocytopenia or other heparin allergy/hypersensitivity<br>• Thrombocytopenia (platelet count <50 x$10^9$/L)<br>• Major bleeding, defined based closely on the International Society on Thrombosis and Haemostasis (ISTH)/Scientific and Standardization Committee (SSC) definitions and bleeding assessment tool in non-surgical patients<br>• Coagulopathy associated with an elevated INR (e.g., >2.0) or hypofibrinogemia<br>• Following invasive procedures where heparin is deemed unsafe to re-institute<br>• Patients requiring systemic fibrinolytic therapy<br>• Treating physician discretion |
| Statistical Analysis: | Data will be analyzed by an intention to treat analysis for the primary analysis; a per-protocol analysis will also be completed as a secondary analysis. Patients who receive at least one dose of drug will be evaluable for safety and efficacy. Response-adaptive randomization based on D-dimer subgroups is embedded. |

ACTIV-4a

| | |
|---|---|
| Title | A Multicenter, Adaptive, Randomized, Open Label Controlled Platform Trial of the Safety and Efficacy of Antithrombotic Strategies in Hospitalized Adults with COVID-19 |
| Short Title | ACTIV-4 ACUTE |
| Brief Summary | This is a randomized, open label, adaptive platform trial to compare the effectiveness of antithrombotic strategies for prevention of adverse outcomes in COVID-19 positive inpatients |
| Objectives | 1. To determine the most effective antithrombotic strategy for increasing the number of days free of organ support and reducing death.<br>2. To determine the most effective antithrombotic strategy on the composite endpoint of death, deep vein thrombosis (DVT), pulmonary embolism (PE), myocardial infarction (MI), ischemic stroke, or other systemic arterial thrombosis (AT).<br>3. To assess the safety of antithrombotic strategies through the endpoint of major bleeding as defined by ISTH.<br>4. To compare the effect of antithrombotic strategies on the endpoint of all-cause mortality in the study population.<br>Assessment of efficacy and safety will yield information of the net clinical benefit of different antithrombotic strategies in the study population. It will also yield information on outcomes specific to under-represented minority populations, specifically African- and Hispanic- descent persons. |
| Methodology | Adaptive Randomized Platform Trial |

| | |
|---|---|
| Endpoints | Primary Endpoint: 21 Day Organ Support Free Days, which is defined as the number of days that a patient is alive and free of organ support through the first 21 days after trial entry. Organ Support is defined as receipt of invasive or non-invasive mechanical ventilation, high flow nasal oxygen, vasopressor therapy, or ECMO support, with death at any time (including beyond 21 days) during the index hospitalization assigned -1 days.<br><br>Key Secondary Endpoint: Composite endpoint of death, pulmonary embolism, systemic arterial thromboembolism, myocardial infarction, or ischemic stroke at hospital discharge or 28 days, whichever occurs first.<br><br>Other Secondary Endpoints: Composite endpoint of death, deep vein thrombosis, pulmonary embolism, systemic arterial thromboembolism, myocardial infarction, or ischemic stroke at hospital discharge or 28 days, whichever occurs first. Acute kidney injury defined by KDIGO criteria, Individual endpoints comprising the key secondary endpoint, death during hospitalization, 28 Day Ventilator-Free Days, 28 Day Vasopressor Free Days, 28 Day Renal Replacement Free Days, WHO clinical scale, 28 Day Hospital Free Days, 28 day organ support free days, and all-cause mortality at 90 days.<br><br>Primary Safety Endpoint: Major bleeding (as defined by the ISTH) Secondary Safety Endpoint: Confirmed heparin induced thrombocytopenia (HIT) |
| Study Duration | Approximately 1 year |
| Participant Duration | Hospital duration with periodic contact at post-discharge, including at 90 days, with potential contact up to 1 year |
| Duration of assigned treatment strategy | During hospitalization (unless otherwise specified in description of arm) |
| Population | Adult patients hospitalized for COVID-19 |
| Study Sites | Approximately 150 sites |
| Number of participants | The sample size is described in each arm-specific appendix. |
| Description of Study Agents | Randomized arms- see appendix<br><br>This platform trial allows for multiple therapies to be investigated in this trial over time. The trial is governed by a Master Protocol that describes the trial design, endpoint collection, primary endpoint, and inclusion/exclusion criteria. Different therapies, referred to as arms, are detailed in arm-specific appendices. These arm-specific appendices work in a modular fashion as arms are removed and added to the platform trial. |
| Key Procedures | Observation during hospitalization, contact at 90 days post-enrollment, and collection of standard of care laboratory results. Ancillary biobanking will be completed in consenting patients at capable centers. |

| | |
|---|---|
| Statistical Analysis | Inferences in this trial are based on a Bayesian statistical model, which considers the variation in outcomes by site, disease state, time, and arm of the trial. The specific analyses for each arm, including interim analysis schedule, are specified in each arm-specific appendix. |

| REMAP-CAP: COVID-19 Therapeutic Anticoagulation Domain Summary | |
|---|---|
| Interventions | • Local standard venous thromboprophylaxis<br>• Therapeutic anticoagulation with intravenous unfractionated heparin or subcutaneous low molecular weight heparin |
| Unit of Analysis, Strata, and State | This domain is analyzed only in the pandemic statistical model.<br><br>The pandemic statistical model includes only patients who are in the Pandemic Infection Suspected or Proven (PISOP) stratum. Within this stratum, the unit-of-analysis is defined by illness severity state at time of enrollment, defined as either Moderate State or Severe State. Unit-of-analysis may also be defined by SARS-CoV-2 infection or d-dimer strata or both. Borrowing is permitted between states and strata. If the SARS-CoV-2 strata is applied in analysis, Response Adaptive Randomization will be applied to all PISOP patients, in each illness severity state, using probabilities derived from the SARS-CoV-2 confirmed stratum. Response Adaptive Randomization may also be applied according to D-dimer strata status. |
| Evaluable treatment-by-treatment Interactions | No interaction will be evaluated with any other domain. |
| Nesting | None |
| Timing of Reveal | Randomization with Immediate Reveal and Initiation or Randomization with Deferred Reveal if prospective agreement to participate is required. |
| Inclusions | Patients will be eligible for this domain if:<br>• COVID-19 infection is suspected by the treating clinician or has been confirmed by microbiological testing<br>• Microbiological testing for SARS-CoV-2 infection of upper or lower respiratory tract secretions or both has occurred or is intended to occur |
| Domain-Specific Exclusions | Patients will be excluded from this domain if they have any of the following:<br>• More than 48 hours has elapsed since ICU admission (noting that this may be operationalized as more than 48 hours has elapsed since commencement of organ failure support)<br>• Clinical or laboratory bleeding risk or both that is sufficient to contraindicate therapeutic anticoagulation, including intention to continue or commence dual anti-platelet therapy<br>• Therapeutic anticoagulation is already present due to prior administration of any anticoagulant agent that is known or likely to still be active or a clinical decision has been made to commence therapeutic anticoagulation<br>• Enrollment in a trial evaluating anticoagulation for proven or suspected COVID-19 infection, where the protocol of that trial requires continuation of the treatment assignment specified in that trial<br>• Known or suspected previous adverse reaction to UFH or LMWH including heparin induced thrombocytopenia (HIT).<br>• The treating clinician believes that participation in the domain would not be in the |

| | |
|---|---|
| | best interests of the patient |
| Intervention-Specific Exclusions | None |

#### Cross-Platform Protocol Comparison Tables

##### Eligibility Criteria

<table>
<tr><th></th><th>REMAP-CAP</th><th>ACTIV-4a</th><th>ATTACC</th></tr>
<tr><td colspan="4">Inclusion Criteria</td></tr>
<tr><td>Age</td><td>• Adult (age not specifically listed)</td><td colspan="2">• ≥ 18 years of age</td></tr>
<tr><td>Duration of hospitalization for COVID-19</td><td>• Expected hospital LOS > 48 hours (i.e. not expected to be discharged today or tomorrow)</td><td colspan="2">• Expected hospital LOS ≥ 72 hours</td></tr>
<tr><td>SARS-CoV-2 infection</td><td>• Suspected or confirmed with intent to test for COVID-19*</td><td colspan="2">• Confirmed</td></tr>
<tr><td>Enrollment Window</td><td>• Less than 48 hours from ICU admission (or initiation of ICU-level care)</td><td colspan="2">• <72 hours from admission OR COVID-19 confirmation</td></tr>
</table>

<table>
<tr><td colspan="4">Exclusion Criteria</td></tr>
<tr><td>Platelet Count</td><td rowspan="2"></td><td>• < 50x $10^9$/L</td><td>• <50 x$10^9$/L, INR >2.0, or baseline aPTT >50 seconds</td></tr>
<tr><td>Hemoglobin</td><td colspan="2">• Hemoglobin <80 g/L (8 g/dL) (to minimize the likelihood of requiring red blood cell transfusion if potential bleeding were to occur)</td></tr>
<tr><td>Heparin Induced Thrombocytopenia (HIT)</td><td>• Known or suspected previous adverse reaction to unfractionated heparin or low molecular weight heparin including HIT</td><td colspan="2">• History of heparin-induced thrombocytopenia (HIT) or other heparin allergy including hypersensitivity</td></tr>
<tr><td>Dual Antiplatelet Therapy</td><td>• Intention to continue or commence dual antiplatelet therapy</td><td>• Patient on dual antiplatelet therapy, when one of the agents cannot be stopped safely</td><td>• Current use of dual antiplatelet therapy</td></tr>
<tr><td>Mechanical Ventilation</td><td></td><td colspan="2">• Chronic mechanical ventilation via tracheostomy prior to hospitalization</td></tr>
<tr><td>Prognosis</td><td>• Death is deemed to be imminent and inevitable during the next 24 hours AND<br>• One or more of the patient, substitute decision maker, or attending physician are not committed to full active treatment</td><td>• Imminent death</td><td>• Patients in whom imminent demise is anticipated and there is no commitment to active ongoing intervention</td></tr>
<tr><td>Co-Enrollment</td><td>• Enrollment in a trial evaluating anticoagulation for proven or suspected</td><td>• Co-enrollment in other trials is permitted as long as</td><td>• Enrollment in other trials related to</td></tr>
</table>

| | | | |
|---|---|---|---|
| | COVID-19 infection, where the protocol of that trial requires continuation of the treatment assignment specified in that trial | the other trial does not test agents with antithrombotic properties and there is no other scientific contraindication | anticoagulation or antiplatelet therapy |
| Bleeding Risk | • Clinical and/or laboratory bleeding risk or both that is sufficient to contraindicate therapeutic anticoagulation | Contraindication to anticoagulation, including but not limited to:<br>• known bleeding within the last 30 days requiring emergency room presentation or hospitalization<br>• known history of an inherited or active acquired bleeding disorder<br>• recent ischemic stroke | • Intracranial surgery or stroke within 3 months<br>• history of intracerebral arteriovenous malformation<br>• cerebral aneurysm or mass lesions of the central nervous system<br>• intracranial malignancy<br>• history of intracranial bleeding<br>• history of bleeding diatheses (e.g., hemophilia)<br>• history of gastrointestinal bleeding within previous 3 months<br>• thrombolysis within the previous 7 days<br>• presence of an epidural or spinal catheter<br>• recent major surgery <14 days<br>• uncontrolled hypertension (sBP >200 mmHg, dBP >120 mmHg)<br>• other physician-perceived contraindications to anticoagulation |
| Miscellaneous | • Treating physician does not feel trial participation is in the best interest of the patient | • Pregnancy | |

Interventions

| | **REMAP-CAP** | **ACTIV-4a** | **ATTACC** |
|---|---|---|---|
| *Intervention arm management* | | | |
| Anticoagulant drug | • Unfractionated heparin or low molecular weight heparin<br>• Patients may be switched between unfractionated heparin and low molecular weight heparin | • Unfractionated heparin or low molecular weight heparin<br>• Patients may be switched between unfractionated heparin and low molecular weight heparin<br>• Patients with impaired renal function were stipulated to received unfractionated heparin | • Unfractionated heparin or low molecular weight heparin<br>• Either agent permitted and patients may be switched between unfractionated heparin and low molecular weight heparin |
| Dose | • Dosed according to local hospital policy, practice, and guidelines for treatment of venous thromboembolism<br>• For UFH, suggested target for aPTT of 1.5 to 2.5 times the upper limit of normal or therapeutic anti-Xa levels<br>• Low molecular weight heparin dosed according to patient weight | • Low molecular weight heparin dosed according to patient weight and creatinine clearance<br>• For UFH, suggested target of anti-Xa of 0.3-0.7 IU/ml or aPTT 1.5 to 2.5 times the upper limit of normal | • Low molecular weight heparin dosed according to patient weight and creatinine clearance according to local practice and policy<br>• For UFH, suggested target of aPTT 1.5 to 2.5 times the upper limit of normal or therapeutic anti-Xa levels |
| Duration of intervention | • Up to 14 days or to hospital discharge, whichever comes first<br>• For ICU patients, therapeutic anticoagulation could be discontinued at ICU discharge | • Up to 14 days or to hospital discharge, whichever comes first | • Up to 14 days or until hospital discharge or recovery (defined as liberation from supplemental oxygen>24 hours, provided oxygen was required), whichever comes first |
| *Usual care arm management* | | | |
| Thromboprophylaxis agent | • Standard venous thromboprophylaxis according to local guidelines or usual practice | • Any one of enoxaparin, dalteparin, tinzaparin, fondaparinux, or heparin according to local preference | • Standard venous thromboprophylaxis according to local guidelines or usual practice |

| | | | |
|---|---|---|---|
| Thromboprophylaxis dose | • Dose of chosen agent should not be sufficient to result in therapeutic anticoagulation | • Dose of agent specified to be consistent with guidelines for low dose thromboprophylaxis | • Dose of chosen agent should not be more than half of the approved therapeutic dose for the treatment of venous thromboembolism |
| Duration of thromboprophylaxis | • Up to 14 days or hospital discharge, whichever comes first<br>• After this period, decisions regarding thromboprophylaxis are at discretion of treating clinician | • Up to 14 days or hospital discharge, whichever comes first<br>• After this period, decisions regarding thromboprophylaxis are at discretion of treating clinician | • Up to 14 days or hospital discharge, whichever comes first<br>• After this period, decisions regarding thromboprophylaxis are at discretion of treating clinician |

Endpoints

| | REMAP-CAP | ACTIV-4a | ATTACC |
|---|---|---|---|
| Primary endpoint | • Days alive and free of organ support to day 21<br>• An ordered categorical endpoint ranging between 0 and 21; patients who die at any time in hospital (through 90 days) are assigned a value of -1 | | |
| Secondary efficacy endpoints | • All-cause mortality at 90 days<br>• Hospital length-of-stay censored at day 90 | • All-cause mortality at 28 days<br>• All-cause mortality in hospital<br>• Hospital re-admission within 28 days<br>• Hospital length-of-stay<br>• WHO ordinal scale at 14 days, proportion with improvement by at least 2 levels compared to enrollment at 28 days | • All-cause mortality at 28 days and 90 days<br>• Hospital re-admission within 28 days<br>• WHO ordinal scale at 14 days, proportion with improvement by at least 2 levels compared to enrollment at 28 days<br>• Intubation at day 30<br>• Ordered categorical endpoint with three possible outcomes based on the worst status of each patient through day 30 (not invasively ventilated, invasively ventilated, or death) |
| Secondary ICU outcomes | • ICU readmission<br>• ICU mortality censored at day 90<br>• ICU length-of-stay censored at day 90<br>• Ventilator-free days at 28 days<br>• Tracheostomy censored at 28 days | • Ventilator-free days at 28 days<br>• Hospital-free days at 28 days<br>• Vasopressor-free days at 28 days<br>• Renal replacement-free days at 28 days<br>• Use of ECMO in hospital | |
| Secondary thrombosis endpoints | • Confirmed deep venous thrombosis in hospital<br>• Confirmed pulmonary embolism<br>• Confirmed ischemic cerebrovascular event<br>• Confirmed acute myocardial infarction<br>• Other thrombotic event including mesenteric ischemia and limb ischemia<br>• Peak troponin between | • Major thrombotic events (death, pulmonary embolism, systemic arterial thromboembolism, myocardial infarction, or ischemic stroke) at 28 days<br>• Symptomatic proximal venous thromboembolism (DVT or PE) assessed at 28 days<br>• Myocardial infarction at 28 days<br>• Ischemic stroke at 28 days<br>• Acute kidney injury as defined by KDIGO criteria<br>• Systemic arterial thrombosis or embolism at 28 days<br>• Mechanical circuit (dialysis or ECMO) thrombosis | |

<table>
<tr><td></td><td>randomization to day 15</td><td></td></tr>
<tr><td>Safety endpoints (assessed during the treatment period)</td><td colspan="2">• Major bleeding, defined according to International Society on Thrombosis and Haemostasis (ISTH) definitions<br>    • Fatal bleeding<br>    • Symptomatic bleeding in a critical area or organ<br>    • Bleeding causing a fall in hemoglobin level of ≥20 g/L<br>    • Requiring a transfusion of 2 or more units of whole blood or red cells<br>• Laboratory-confirmed heparin-induced thrombocytopenia</td></tr>
</table>

#### Categorization of frequently used heparin doses in the ATTACC, ACTIV-4a, and REMAP-CAP mpRCT

Drugs/doses for prophylaxis in the mpRCT falling outside of those below are manually categorized, considering participant body weight/BMI and renal function, informed by American Society of Hematology.[2]

##### Subcutaneous Enoxaparin:

*Low dose:*

- *Standard dose*: 40mg once daily
- *Possible*: If BMI ≥ 40 kg/$m^2$ (or weight ≥ 120 kg) and CrCl ≥ 30 mL/min: 40mg twice daily
- *Possible*: If CrCl < 30 mL/min: 30 mg once daily

*Intermediate dose:*

Twice daily:

- *Standard dose*: up to and including (a) 0.5 mg/kg twice daily + 20% (rounding factor) or (b) 40mg twice daily (whichever is higher)
- *Possible*: If BMI ≥ 40 kg/$m^2$ (or weight ≥ 120 kg) and CrCl ≥ 30 mL/min: up to 60 mg twice daily
- *Possible*: If CrCl < 30 mL/min: intermediate twice daily dose not defined

Daily:

- *Standard dose*: up to 1.0 mg/kg once daily + 20% (rounding factor)
- *Possible*: If BMI ≥ 40 kg/$m^2$ (or weight ≥ 120 kg) and CrCl ≥ 30 mL/min: up to 0.8 mg/kg once daily + 20% (rounding factor)
- *Possible*: If CrCl < 30 mL/min (and weight ≥ 60 kg): up to 0.5 mg/kg once daily +20% (rounding factor)

*Subtherapeutic dose:*

- Between intermediate and therapeutic doses

*Therapeutic dose:*

Twice daily:

- *Standard dose:* starting at 1mg/kg twice daily minus 10% (rounding factor)
- *Possible:* If BMI ≥ 40kg/$m^2$ (or weight ≥ 120kg) and CrCl ≥ 30 mL/min: starting at 0.8 mg/kg twice daily minus 10% (rounding factor)
- *Possible:* If CrCl < 30 mL/min: therapeutic twice daily dose not defined

Daily:

- *Standard dose:* starting at 1.5mg/kg once daily minus 10% (rounding factor)
- *Possible:* CrCl < 30 mL/min: starting at 1mg/kg once daily minus 10% (rounding factor)
- *Possible:* If BMI ≥ 40kg/$m^2$ and CrCl ≥ 30 mL/min: therapeutic once daily dose not defined

##### Subcutaneous Dalteparin:

*Low dose:*

- *Standard dose*: 5,000 units once daily

- *Possible*: If BMI ≥ 40 $kg/m^2$ (or weight ≥120 kg) and CrCl ≥ 30 mL/min: 7,500 units once daily

*Intermediate dose:*

- *Standard dose*: 5,000 units twice daily
- *Possible*: If BMI ≥ 40 $kg/m^2$ (or weight ≥ 120 kg) and CrCl ≥ 30 mL/min: 7,500 units twice daily

*Subtherapeutic dose:*

- Between intermediate and therapeutic doses

*Therapeutic dose:*

Twice daily:

- *Standard dose:* 100 U/kg twice daily minus 10% (rounding factor)

Daily:

- *Standard dose:* starting at 200 U/kg once daily minus 10% (rounding factor)

Subcutaneous Tinzaparin:

*Low dose:*

- *Standard dose*: up to and including (a) 75 anti-Xa units/kg + 20% (rounding factor) once daily or (b) 4,500 units once daily (whichever is higher)
- *Possible*: If BMI ≥ 40 $kg/m^2$ (or weight ≥ 120 kg) and CrCl ≥ 30 mL/min: 8,000 units once daily

*Intermediate dose*:

- *Standard dose*: 4,500 units twice daily
- *Possible*: If BMI ≥ 40 $kg/m^2$ (or weight ≥ 120 kg) and CrCl ≥ 30 mL/min: 8,000 units twice daily

*Subtherapeutic dose:*

- Between intermediate and therapeutic doses

*Therapeutic dose:*

- *Standard dose:* 175 anti-Xa units/kg once daily minus 10% (rounding factor)

Unfractionated heparin:

*Low dose (subcutaneous):*

- *Standard dose*: 5,000 units twice or three times daily
- *Possible*: If BMI ≥ 40 $kg/m^2$ (or weight ≥ 120 kg) and CrCl ≥ 30 mL/min: 7,500 units twice daily

*Intermediate dose (subcutaneous):*

- *Standard dose*: 7,500 units three times daily or 10,000 units twice daily
- *Possible*: If BMI ≥ 40 $kg/m^2$ (or weight ≥ 120 kg) and CrCl ≥ 30 mL/min: 10,000 units twice daily

*Subtherapeutic dose:*

- Not defined for unfractionated heparin

*Therapeutic dose (intravenous):*

- *Standard dose:* continuous intravenous administration per local protocol

### Endpoint Definitions

#### ATTACC

The full list of secondary endpoints is available in the trial protocol. Among these, the following secondary efficacy endpoints will be adjudicated by the CEC:

- Venous thromboembolism (DVT or PE) assessed at 28 and 90 days following randomization.
- Myocardial infarction assessed at 28 and 90 days following randomization.
- Ischemic stroke assessed at 28 and 90 days following randomization.
- Systemic arterial thrombosis or embolism assessed at 28 and 90 days following randomization.

The following secondary safety events will be adjudicated by the CEC (sites instructed to report events occurring during the intervention window as defined in the protocol):

- Laboratory confirmed HIT
- Major bleeding, defined according to the International Society on Thrombosis and Haemostasis (ISTH)/Scientific and Standardization Committee (SSC) definitions and bleeding assessment tool in non-surgical patients[3]

ATTACC endpoint definitions are as described below for ACTIV-4a

#### ACTIV-4a

The full list of secondary endpoints is available in the trial protocol. The CEC will consider for adjudication all cases of the following:

- Deep venous thromboembolism
- Pulmonary embolism
- Arterial thromboembolism
- Myocardial infarction
- Stroke
- Major bleeding
- Death due to cardiovascular, non-cardiovascular, and undetermined cause

*Deep Venous Thromboembolism*

The diagnosis of definite symptomatic deep venous thromboembolism (DVT) requires symptoms of venous thromboembolism with at least one of the following:

- Abnormal compression ultrasound consistent with DVT or abnormal flow pattern or direct clot visualization in veins not amenable to compression.
- One or more new filling defects by venography, CT venography, or MR venography.
- Abnormal compression ultrasound where compression had been normal or, if known to be non-compressible, a substantial increase (≥4mm) in the diameter of a previously non-compressible venous segment.

- Point-of-care ultrasound (POCUS) performed by a provider and documenting DVT in a note.
- An extension of an intraluminal filling defect, or a new intraluminal filling defect, or an extension of non-visualization of veins in the presence of a sudden cut-off on venography.
- Proximal DVT is defined as clot at or proximal to the trifurcation of the popliteal vein (in the lower extremity) OR clot at or proximal to the axillary vein segment (in the upper extremity).
- Distal DVT is defined as clot distal to the trifurcation of the popliteal vein (in the lower extremities) OR clot at or distal to the brachial vein segment (in the upper extremities).
- Non-limb venous thrombosis includes thrombosis of the cerebral, portal, mesenteric, hepatic, gonadal, splenic, renal, or retinal veins, or thrombosis of the superior or inferior vena cava.

The diagnosis of <u>presumed</u> deep venous thromboembolism requires the following:

- In the absence of objective testing, high pre-test probability according to investigator assessment
    - OR adjudicator's gestalt
    - OR Wells score
- AND a treatment plan for DVT was initiated (initiation of anticoagulation, or escalation of anticoagulation dose, frequency, or duration).

*Pulmonary Embolism*

The diagnosis of <u>definite</u> pulmonary embolism requires at least one of the following:

- New intraluminal filling defect at CT pulmonary angiography in a subsegmental or larger vessel.
- New intraluminal filling defect, or an extension of an existing defect, or a new sudden cut-off of vessels > 2.5 mm in diameter at pulmonary angiogram,
- Inconclusive CT pulmonary angiography, pulmonary angiography, or VQ scan evidence of a new or recurrent PE with demonstration of a new or recurrent DVT in the lower extremities by compression ultrasonography or venography.[4,5]
- New clot or intraluminal filling defect noted in the right heart ("clot in transit") or the pulmonary vasculature at echocardiogram
- High probability (revised PIOPED criteria) on planar ventilation/perfusion (V/Q) scan OR positive PE on SPECT ventilation perfusion (V/Q) scan.
- Pulmonary embolism found at autopsy

The diagnosis of <u>presumed</u> pulmonary embolism requires the following:
Clinical signs and symptoms of pulmonary embolism, including but not limited to: dyspnea, cough, hypoxemia, tachycardia, appropriate electrocardiographic changes, or evidence of right heart strain on echocardiogram; AND chest CT or pulmonary angiography are unable to be performed AND therapeutic dose anticoagulation or fibrinolytic therapy is prescribed by a physician

*Arterial Thromboembolism*

The diagnosis of arterial thromboembolism is defined as the following:

- A clinical history and presentation consistent with a sudden significant worsening of end organ or limb perfusion AND

EITHER

- Confirmation of arterial obstruction by imaging, hemodynamics, intraoperative findings, or pathological evaluation

OR

- Requirement for thrombolysis, thrombectomy, or urgent bypass.

Note that arterial thromboembolism includes both acute *in situ* thrombotic events and acute embolic events. Note that while ischemic stroke and myocardial infarction can be arterial thromboembolic events, those events will be adjudicated according to the separate standardized criteria included below.

##### *Myocardial Infarction*

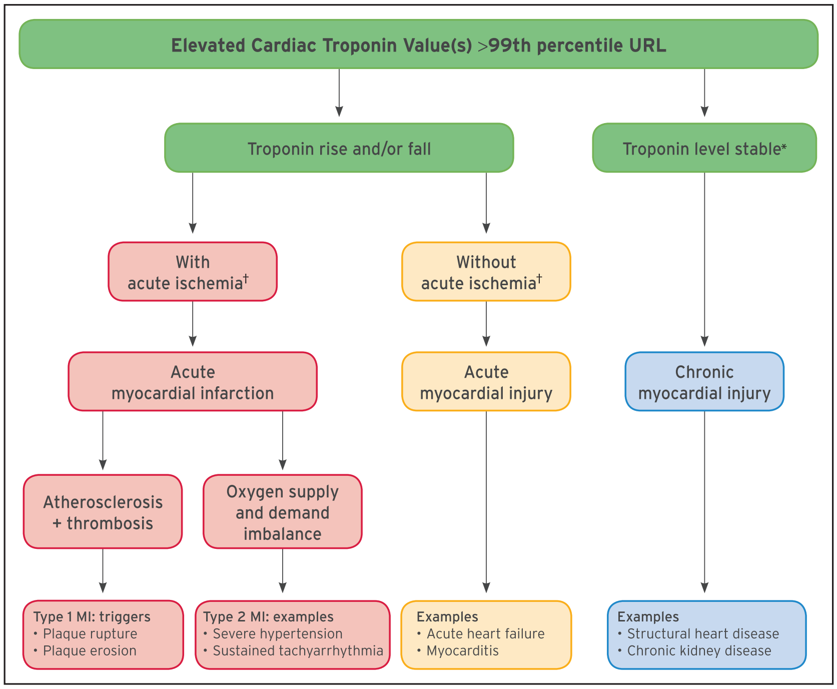

**Figure 1**. Model for interpreting myocardial injury.[1].[1]

COVID-19 patients are well known to have elevations in cardiac troponin concentrations, and these elevations often do not represent arterial thrombosis and downstream myocardial ischemia. Therefore, the CEC will make an effort to distinguish true myocardial infarction from coronary artery obstruction, typically from atherothrombosis (usually considered a "type 1 myocardial infarction") from myocardial infarction due to demand ischemia (usually defined as a "type 2 myocardial infarction") and myocardial injury (an elevation in cardiac troponin typically without symptoms of chest pain or signs of arterial thrombosis). These definitions will be consistent with the 4th Universal Definition of Myocardial Infarction and will take into considerations suggestions made about classification of certain conditions as type 1 as compared to type 2 myocardial infarction.[1,6] Regional coronary venous thrombosis with associated regional myocardial infarction has been reported in COVID. If this mechanism is documented, these will be considered a type 1 MI. The trial and CEC are focused on ascertaining and adjudicating cases of acute myocardial injury and acute myocardial infarction and classifying those cases as described below. COVID also causes microvascular thrombi which are associated with patchy myocardial necrosis. These will be grouped with myocardial injury.

#### 2. UNIVERSAL DEFINITIONS OF MYOCARDIAL INJURY AND MYOCARDIAL INFARCTION: SUMMARY

| Universal definitions of myocardial injury and myocardial infarction |
|---|
| **Criteria for myocardial injury** |
| The term myocardial injury should be used when there is evidence of elevated cardiac troponin values (cTn) with at least 1 value above the 99th percentile upper reference limit (URL). The myocardial injury is considered acute if there is a rise and/or fall of cTn values. |
| **Criteria for acute myocardial infarction (types 1, 2 and 3 MI)** |
| The term acute myocardial infarction should be used when there is acute myocardial injury with clinical evidence of acute myocardial ischemia and with detection of a rise and/or fall of cTn values with at least 1 value above the 99th percentile URL and at least 1 of the following:<br>• Symptoms of myocardial ischemia;<br>• New ischemic ECG changes;<br>• Development of pathological Q waves;<br>• Imaging evidence of new loss of viable myocardium or new regional wall motion abnormality in a pattern consistent with an ischemic etiology;<br>• Identification of a coronary thrombus by angiography or autopsy (not for types 2 or 3 MIs).<br>Postmortem demonstration of acute atherothrombosis in the artery supplying the infarcted myocardium meets criteria for *type 1 MI*. Evidence of an imbalance between myocardial oxygen supply and demand unrelated to acute atherothrombosis meets criteria for *type 2 MI*. Cardiac death in patients with symptoms suggestive of myocardial ischemia and presumed new ischemic ECG changes before cTn values become available or abnormal meets criteria for *type 3 MI*. |
| **Criteria for coronary procedure–related myocardial infarction (types 4 and 5 MI)** |
| Percutaneous coronary intervention (PCI)–related MI is termed *type 4a MI*.<br>Coronary artery bypass grafting (CABG)–related MI is termed *type 5 MI*.<br>Coronary procedure–related MI ≤48 hours after the index procedure is arbitrarily defined by an elevation of cTn values >5 times for *type 4a MI* and >10 times for *type 5 MI* of the 99th percentile URL in patients with normal baseline values. Patients with elevated preprocedural cTn values, in whom the preprocedural cTn level are stable (≤20% variation) or falling, must meet the criteria for a >5 or >10 fold increase and manifest a change from the baseline value of >20%. In addition with at least 1 of the following:<br>• New ischemic ECG changes (this criterion is related to *type 4a MI* only);<br>• Development of new pathological Q waves;<br>• Imaging evidence of loss of viable myocardium that is presumed to be new and in a pattern consistent with an ischemic etiology;<br>• Angiographic findings consistent with a procedural flow-limiting complication such as coronary dissection, occlusion of a major epicardial artery or graft, side-branch occlusion-thrombus, disruption of collateral flow or distal embolization.<br>Isolated development of new pathological Q waves meets the *type 4a MI* or *type 5 MI* criteria with either revascularization procedure if cTn values are elevated and rising but less than the prespecified thresholds for PCI and CABG.<br>Other types of 4 MI include *type 4b MI* stent thrombosis and *type 4c MI* restenosis that both meet *type 1 MI* criteria.<br>Postmortem demonstration of a procedure-related thrombus meets the *type 4a MI* criteria or *type 4b MI* criteria if associated with a stent. |
| **Criteria for prior or silent/unrecognized myocardial infarction** |
| Any 1 of the following criteria meets the diagnosis for prior or silent/unrecognized MI:<br>• Abnormal Q waves with or without symptoms in the absence of nonischemic causes.<br>• Imaging evidence of loss of viable myocardium in a pattern consistent with ischemic etiology.<br>• Patho-anatomical findings of a prior MI. |

CABG indicates coronary artery bypass grafting; cTn, cardiac troponin; ECG, electrocardiogram; MI, myocardial infarction; PCI, percutaneous coronary intervention; URL, upper reference limit.

**Figure 2**. Table from the 4$^{th}$ Universal Definition of Myocardial Infarction summarizing the different definitions of myocardial injury and infarction.[1]

**Myocardial Injury:** The increasing sensitivity of cardiac troponin (cTn) assays means that ongoing myocardial injury is frequently detected. Myocardial injury is a prerequisite for myocardial infarction (MI), but as noted below, criteria in addition to myocardial injury are necessary to make the diagnosis of MI. Adjudicators must distinguish between acute myocardial injury that is not secondary to ischemia but may be due to other conditions (Table 2).

<u>Criteria for Myocardial Injury:</u> Detection of an elevated cTn value above the 99$^{th}$ percentile upper reference limit (URL) is defined as myocardial injury. The injury is considered acute if there is a rise and/or fall of cTn values.[1]

| Table 2. Causes of non-ischemic myocardial injury[2, 6] |
|---|
| Heart failure |
| Myocarditis |
| Cardiomyopathy |
| Takotsubo syndrome |
| Coronary revascularization procedure |
| Cardiac procedure other than revascularization |
| Catheter ablation |
| Defibrillator shocks |
| Cardiac contusion |
| Sepsis, infectious disease |
| Chronic kidney disease |
| Stroke, subarachnoid hemorrhage |
| Pulmonary embolism, pulmonary hypertension |
| Infiltrative disease, e.g., amyloidosis, sarcoidosis |
| Chemotherapeutic agents |
| Critically ill patients |
| Strenuous exercise |
| Other |

<u>Criteria for Procedure Related Myocardial Injury:</u> Cardiac procedural myocardial injury is arbitrary defined by increased in cTn values (>99th percentile URL) in patients with normal baseline values (<99th percentile URL) or a rise of cTn values >20% of the baseline value when it is the above the 99th percentile URL but is stable or falling.

**Myocardial Infarction Type 1**: Detection of rise and/or fall of cardiac biomarkers with at least one value above the 99th percentile of the upper reference limit (URL) together with evidence of myocardial ischemia with at least one of the following:

- Symptoms of ischemia
- New ischemic ECG changes indicative of new ischemia (new ST-T changes or new LBBB)*
- Development of pathological Q waves in the ECG**
- Imaging evidence of new loss of viable myocardium or new regional wall motion abnormality in a pattern consistent with an ischemic etiology
- Identification of a coronary thrombus by angiography including intracoronary imaging or by autopsy†
- *ECG manifestation of acute myocardial ischemia (in the absence of LVH and LBBB):
    - ST Elevation: New ST elevation at the J-point in two contiguous leads with the cut-point: ≥ 1 mm in all leads other than leads V2-V3, where the following cut-points apply: ≥2mm in men ≥40 years; ≥2.5 mm in men <40 years; or ≥ 1.5 mm in women regardless of age.
    - ST-depression and T-wave changes: New horizontal or down-sloping ST depression ≥ 0.5 mm in 2 contiguous leads and/or T inversion ≥ 1 mm in two contiguous leads with prominent R waves or R/S ratio >1.
- **Pathological Q waves:
    - Any Q-wave in leads V2-V3 >0.02 seconds or QS complex in leads V2-V3
    - Q-wave ≥ 0.03 seconds and ≥1 mm deep or QS complex in leads I, II, aVL, aVF, or V4-V6 in any 2 leads of a contiguous lead grouping (I, aVL;V1-V6; II, III, aVF; V7-V9).
    - R-wave ≥ 0.04s in V1–V2 and R/S ≥ 1 with a concordant positive T-wave in the absence of a conduction defect
- †Postmortem demonstration of an atherothrombus in the artery supplying the infarcted myocardium, or a macroscopically large circumscribed area of necrosis with or without intramyocardial hemorrhage meets the type 1 MI criteria regardless of cTn values.
- Consideration will be given to recent proposals to modify myocardial infarction type 1 to include coronary obstruction by spontaneous coronary artery dissection, coronary embolism, or coronary vasospasm or microvascular dysfunction.[5]

**Myocardial Infarction Type 2:** Detection of a rise and/or fall of cTn values with at least 1 value above the 99$^{th}$ percentile URL, and evidence of imbalance between myocardial oxygen supply and demand unrelated to coronary atherothrombosis, requiring at least 1 of the following:

- Symptoms of acute myocardial ischemia
- New ischemic ECG changes
- Development of pathological Q waves;
- Imaging evidence of new loss of viable myocardium or new regional wall motion abnormality in a pattern consistent with ischemic etiology

**Myocardial Infarction Type 3:** Patients who suffer cardiac death, with symptoms suggestive of myocardial ischemia accompanied by presumed new ischemic ECG changes or ventricular fibrillation but die before blood samples for biomarkers can be obtained, or before increases in cardiac biomarkers can be identified, or MI is detected by autopsy examination.

**Myocardial infarction Type 4a and 4b (myocardial infarction associated with percutaneous coronary intervention):** Criteria for percutaneous coronary intervention (PCI)-related MI ≤48 hours after the index procedure are as follows: Coronary intervention-related MI is arbitrarily defined by an elevation of cTn values >5 times the 99the percentile URL in patients with normal baseline values. In patients with elevated preprocedural cTn in whom the cTn levels are stable (≤20% variation) or falling, the post procedure cTn must rise by >20%. However, the absolute procedural value must still be at least 5 times the 99$^{th}$ percentile URL. In addition, 1 of the following elements is required:

- New ischemic ECG changes
- Development of new pathological Q waves; note that the development of new pathological Q waves meets the criteria for procedure-related MI if the cTn values are elevated and rising but <5 times the 99$^{th}$ percentile URL.
- Imaging evidence of new loss of viable myocardium or new regional wall motion abnormality in a pattern consistent with an ischemic etiology
- Angiographic findings consistent with a procedural flow-limiting complication such as coronary dissection, occlusion of a major epicardial artery or side branch occlusion/thrombus, disruption of collateral flow, or distal embolization.
- Type 4a MI is an MI associated with PCI
- Type 4b MI is an MI associated with stent/scaffold thrombosis

**Myocardial Infarction Type 4c**: A type 4c MI is an MI associated with restenosis associated with prior PCI. Possible Type 4c MI is evaluated using the same criteria as Type 1 MI.

**Myocardial Infarction Type 5**: Criteria of coronary artery bypass grafting (CABG)-related MI ≤ 48 hours after the index procedure. CABG-related MI is arbitrarily defined as elevation of cTn values >10 times the 99$^{th}$ percentile URL in patients with normal baseline cTn values. In patients with elevated preprocedural cTn in whom cTn are stale (≤20% variation) or falling, the post procedure cTn must rise by >20%. However, the absolute postprocedural values must still be >10 times the 99$^{th}$ percentile URL. In addition, one of the following elements is required:

- Development of new pathological Q waves; note that the development of new pathological Q waves meets the criteria for procedure-related MI if the cTn values are elevated and rising but <10 times the 99$^{th}$ percentile URL.
- Angiographically documented new graft occlusion or new native coronary artery occlusion;

- Imaging evidence of new loss of viable myocardium or new regional wall motion abnormality in a pattern consistent with an ischemic etiology

**Special or unusual circumstances:** Further guidance on distinguishing myocardial injury from myocardial infarction in the context of non-cardiac surgery, heart failure, myocarditis, Takotsubo syndrome, kidney disease, and in critically ill patients, and myocardial infarction nonobstructive coronary arteries is included in the 4th Universal Definition of MI.[1]

##### *Stroke*

The definition of stroke used here is drawn from the definitions proposed by Hicks et al. and Sacco et al.[8,9] Stroke is defined as the acute onset of focal neurological dysfunction caused by brain, spinal cord, or retinal vascular injury as a result of hemorrhage or infarction.

A stroke is the acute onset of a new persistent neurological deficit attributed to an obstruction in cerebral blood flow with no apparent nonvascular cause (e.g. tumor, trauma, infection). Available neuroimaging studies will be considered to support the clinical impression and to determine if there is a demonstrable lesion compatible with an acute stroke. To the extent possible, all strokes will be classified as ischemic, hemorrhagic or unknown.

For the diagnosis of stroke, the following criteria should be fulfilled:

**1.** Rapid onset of a focal neurological deficit not related to any other known non-cerebrovascular process with at least one of the following:

- Change in level of consciousness
- Hemiplegia
- Hemiparesis
- Numbness or sensory loss affecting one side of the body
- Dysphasia/aphasia
- Hemianopia
- Other new neurological sign/symptom(s) consistent with stroke
- If the timing of onset is uncertain, a diagnosis of stroke may be made provided that there are no plausible non-stroke causes for the clinical presentation.

AND

**2.** Duration of a focal/global neurological deficit that is:

- EITHER ≥ 24 hours,
- OR < 24 hours if:
    - Resolution of symptoms is due to least one of the following interventions:
        1. Pharmacologic: intravenous or intraarterial thrombolysis
        2. Non-pharmacologic: (i.e. neuro-interventional procedure such as intracranial angioplasty)
    - OR available MRI clearly documents a new hemorrhage or infarct
    - OR available head CT clearly documents a new hemorrhage or infarct or excludes a mimic of stroke
    - OR the neurological deficit results in death.

Ideally, at least one of should be present to confirm the diagnosis of stroke:

- Confirmation by neurology or neurosurgery specialist
- Brain imaging procedure (at least one of the following): CT scan, MRI scan, or cerebral vessel angiography
- Lumbar puncture (i.e. spinal fluid analysis diagnostic of intracranial hemorrhage)

If the acute focal signs represent a worsening of a previous deficit, these signs must persist for more than 24 hours and be accompanied by an appropriate new MRI or CT scan finding.

Strokes are sub-classified as follows:

Ischemic (non-hemorrhagic): An acute episode of focal cerebral, spinal, or retinal dysfunction caused by infarction of central nervous system tissue. Hemorrhage may be a consequence of ischemic stroke. In this situation, the stroke is an ischemic stroke with hemorrhagic transformation and not a hemorrhagic stroke but would also be listed as a major bleeding safety event.

Hemorrhagic: An acute episode of focal or global cerebral or spinal dysfunction caused by intraparenchymal, intraventricular, or subarachnoid hemorrhage. Hemorrhage in the brain is documented by neuroimaging or autopsy or lumbar puncture. Note that subdural hematomas are intracranial hemorrhagic events and not strokes.

Undetermined: An acute episode of focal or global neurological dysfunction caused by presumed brain, spinal cord, or retinal vascular injury as a result of hemorrhage or infarction but with insufficient information to allow categorization as either ischemic or hemorrhagic.

##### *Major Bleeding*

Major bleeding is defined as acute clinically overt bleeding associated with one or more of the following (as per ISTH guidelines):[3,10]

- Decrease in hemoglobin of 2 g/dL or more;
- Transfusion of 2 units or more of packed red blood cells;
- Bleeding that occurs in at least one of the following critical sites:
    - Intracranial
    - Intraspinal
    - Intraocular (within the corpus of the eye. A conjunctival bleed is not an intraocular bleed)
    - Pericardial
    - Intraarticular
    - Retroperitoneal
    - Intramuscular with compartment syndrome
- Bleeding that leads to death (primary cause of death or contributes directly to death)

##### *Definitions of Cardiovascular, Non-cardiovascular, and Undetermined Cases of Death*

Definitions of Cardiovascular, Non-Cardiovascular, and Undetermined Cases of Death
The classifications for death are drawn from Hicks et al.[8] Death is classified into one of three categories: cardiovascular, non-cardiovascular, and undetermined cause of death. The intent is to

identify one of these categories as the underlying cause of death. The key priority is differentiating between cardiovascular and non-cardiovascular causes of death. Death attribution can be difficult, particularly for sudden death, even when witnessed.

Cardiovascular death can be due to acute myocardial infarction (MI), sudden cardiac death, heart failure, stroke, pulmonary embolism, a cardiovascular procedure, cardiovascular hemorrhage, or other cardiovascular cause.

Cardiovascular death due to acute MI: Death by any cardiovascular mechanism (arrhythmia, sudden death, heart failure, stroke, pulmonary embolus, PAD) within 30 days after an acute MI, related to the immediate consequences of the MI, such as progressive heart failure or recalcitrant arrhythmia. While there may be assessable (attributable) mechanisms of cardiovascular death during this time period, for simplicity, if the cardiovascular death occurs within 30 days of an acute MI, it will be considered a death due to MI.
Note: Acute MI should be verified to the extent possible by the diagnostic criteria outlined for acute MI or by autopsy findings showing recent MI or recent coronary thrombosis. Death resulting from a procedure to treat an MI (PCI or CABG), or to treat a complication resulting from MI, should also be considered death due to acute MI. Death resulting from an elective coronary procedure to treat myocardial ischemia (i.e., chronic stable angina) or death due to an MI that occurs as a direct consequence of a cardiovascular investigation/ procedure/operation should be considered as a death due to a cardiovascular procedure.

Cardiovascular death due to sudden cardiac death:

Death that occurs unexpectedly, and not within 30 days of an acute MI.

Sudden cardiac death includes the following scenarios:

- Death witnessed and occurring without new or worsening symptoms
- Death witnessed within 60 min of the onset of new or worsening cardiac symptoms unless the symptoms suggest acute MI
- Death witnessed and attributed to an identified arrhythmia (e.g., captured on an electrocardiographic recording, witnessed on a monitor, or unwitnessed but found on ICD review)
- Death after unsuccessful resuscitation from cardiac arrest (e.g., ICD unresponsive sudden cardiac death, pulseless electrical activity arrest)
- Death after successful resuscitation from cardiac arrest and without identification of a specific cardiac or non-cardiac etiology
- Unwitnessed death in a subject seen alive and clinically stable ≤24 h before being found dead without any evidence supporting a specific non- cardiovascular cause of death (information about the patient's clinical status preceding death should be provided if available)

Unless additional information suggests an alternate specific cause of death (e.g., Death due to Other Cardiovascular Causes), if a patient is seen alive ≤24 h before being found dead, sudden cardiac death should be recorded. For patients who were not observed alive within 24 h of death, undetermined cause of death should be recorded (e.g., a subject found dead in bed but who had not been seen by family members for >24 h).

Cardiovascular death due to heart failure (HF): Death associated with clinically worsening symptoms and/or signs of HF, regardless of HF etiology. Note: Deaths due to HF can have various etiologies, including single or recurrent MIs, ischemic or nonischemic cardiomyopathy, hypertension, or valvular disease.

Death due to stroke: Death after a stroke that is either a direct consequence of the stroke or a complication of the stroke. Note: acute stroke should be verified to the extent possible by the diagnostic criteria outlined for stroke.

Cardiovascular death due to cardiovascular procedure: Death caused by the immediate complication(s) of a cardiovascular procedure.

Cardiovascular death due to cardiovascular hemorrhage: Death related to hemorrhage such as a non-stroke intracranial hemorrhage (e.g., subdural hematoma) nonprocedural or non-traumatic vascular rupture (e.g., aortic aneurysm), or hemorrhage causing cardiac tamponade.

Cardiovascular death due to other cardiovascular causes: Cardiovascular death not included in the above categories with specific, known cause (e.g., PE, PAD).

Definition of Non-cardiovascular Death: When death is due to a non-cardiovascular cause, a cardiovascular cause of death is excluded.

- Pulmonary (excludes malignancy)
- Renal
- Gastrointestinal (disease of the esophagus, stomach, or intestines (excludes malignancy)
- Hepatobiliary (disease of the liver, gall bladder, or biliary ducts (excludes malignancy)
- Pancreatic (disease of the pancreas (excludes malignancy)
- Infection (including sepsis)
- Inflammatory/immune (death attributable to an inflammatory or immune-mediated disease or process, including systemic inflammatory response syndrome (SIRS), immunological, and autoimmune disease and disorders. Includes anaphylaxis from environmental allergies)
- Hemorrhage (bleeding that is not considered cardiovascular hemorrhage or stroke
- Non-CV procedure or surgery (death caused by the immediate complications of a non-cardiovascular procedure or surgery)
- Trauma (death attributable to trauma. Includes homicide)
- Suicide
- Nonprescription drug reaction or overdose
- Prescription drug reaction or overdose (includes anaphylaxis)
- Neurological (excludes malignancy, as well as death from ischemic stroke, hemorrhagic stroke, or undetermined cause of stroke, or cardiovascular hemorrhage of central nervous system)
- Malignancy (leukemia, lymphoma, or other malignancy)
- Other (death attributable to a cause other than those listed in this classification; specify organ system)

Undetermined cause of death: Causality may be difficult to determine if information available from the time of death is minimal or nonexistent.

REMAP-CAP

The following are the definitions for the outcomes reported by site investigators, as outlined in the Domain Specific Appendix (DSA) and/or the Case Report Form Data Completion Guide.

*Major bleeding*

Fatal bleeding, symptomatic or clinically manifest bleeding in a critical are or organ (such as intracranial, intraspinal, intraocular, retroperitoneal, intraarticular, pericardial, or intramuscular with compartment syndrome),

OR

Blood loss above 300mls or bleeding causing a fall in haemoglobin of ≥2g/dL (20g/L, 1.24mmol/L), or leading to the transfusion of 2 or more whole blood or red cell units

*Acute Myocardial Infarction (AMI)*

The definition of an AMI requires detection of rise and fall or just a fall of cardiac biomarkers, such as any form of troponin assay, with at least one value above the upper reference limit (URL) PLUS evidence of myocardial ischemia with at least one of the following:

- Symptoms of cardiac ischemia
- ECG changes indicative of new ischemia (new ST-T changes or new LBBB)*
- Development of pathological Q waves in the ECG**
- Imaging evidence of new loss of viable myocardium or new regional wall motion abnormality

*ECG manifestation of acute myocardial ischemia (in the absence of LVH and LBBB):

- ST Elevation - New ST elevation at the J-point in two contiguous leads with the cut-off points of ≥ 0.2 mV in men or ≥ 0.15 mV in women in leads V2-V3 and/or ≥ 0.1 mV in other leads.
- ST depression and T-wave changes – New horizontal or down-sloping ST depression ≥ 0.05 mV in two contiguous leads; and/or T inversion ≥ 0.1 mV in two contiguous leads with prominent R waves or R/S ratio >1.

**Pathological Q waves:

- Any Q-wave in leads V2-V3 ≥ 0.02 seconds or QS complex in leads V2 and V3
- Q-wave ≥ 0.03 seconds and ≥ 0.1 mV deep or QS complex in leads I, II, aVL, aVF, or V4-V6 an any two leads of a contiguous lead grouping (I, aVL, V6; V4-V6; II, III, aVF; V7-V9).
- R-wave ≥ 0.04 s in V1–V2 and R/S ≥ 1 with a concordant positive T-wave in the absence of a conduction defect

*Confirmed deep vein thrombosis*

Proximal deep vein thrombosis is a thrombus located in axillary vein or more proximal, including the internal jugular vein, and a thrombus located in popliteal vein or more proximal. Confirmation requires imaging with techniques that include ultrasound or CT scan.

*Confirmed pulmonary embolus*

Segmental or multi-sub-segmental pulmonary emboli that is confirmed using CT pulmonary angiography or has a high probability ventilation: perfusion lung scan

*Confirmed ischemic cerebrovascular event*

An acute ischemic stroke is defined as central nervous system infarction defined as brain, spinal cord, or retinal cell death attributable to ischemia, based on:

- Pathological, imaging, or other objective evidence of cerebral, spinal cord, or retinal focal ischemic injury in a defined vascular distribution;

OR

- Clinical evidence of cerebral, spinal cord, or retinal focal ischemic injury based on symptoms persisting ≥24 hours or until death, and other etiologies excluded. (Note: CNS infarction includes types I and II hemorrhagic infarctions) OR
- Infarction or hemorrhage in the brain, spinal cord, or retina because of thrombosis of a cerebral venous structure. Symptoms or signs caused by reversible oedema without infarction or hemorrhage do not qualify as stroke.

*Mesenteric ischemia*

Mesenteric Ischemia for arterial or venous mesenteric ischemia diagnosed on contrast imaging by CT or angiography or diagnosed at laparotomy or via laparoscopy.

*Limb ischemia*

Limb ischemia if evidence of acute limb ischemia sufficient to require surgical revascularization including bypass procedure, intraarterial thrombolysis, or embolectomy; amputation of a limb due to acute ischemia; or decision to withdraw or limit treatment because of acute limb ischemia. It is not sufficient for there to be evidence of limb ischemia that does not result in surgical intervention or determine a decision to institute palliative care. Ischemia attributed to vasopressor medication is insufficient unless also meets the above definition.

*Heparin induced thrombocytopenia (HIT)*

Definite HIT:

- Positive Serotonin Release Assay (SRA) or equivalent functional HIT confirmatory test
- Positive Enzyme-linked immunosorbent assay (ELISA) AND treated as HIT Positive
- Quantitative rapid immunoassay (RIA) AND treated as HIT

Possible HIT:

- Positive ELISA or quantitative RIA AND any heparin/LMWH anticoagulation stopped
- 4T score >3 (high or intermediate), no Laboratory testing for HIT done, AND treated as HIT

No HIT:

- Negative HIT ELISA, or Negative Quantitative RIA
- Negative SRA, or Negative equivalent functional confirmatory assay
- No laboratory testing for HIT done AND not treated as HIT (heparin continued or discontinued for other reason and uneventful course - defined as absence of thrombotic event and platelet count recovery during follow-up).

*Derived endpoints*

The following are the definitions for the endpoints outlined in the Statistical Analysis Plan for the Anticoagulation Domain which are derived from the domain-specific secondary endpoints:

Systemic arterial thrombosis or embolism

Clinical evidence of sudden significant worsening of organ or limb perfusion and either confirmation of arterial obstruction (e.g. by imaging, hemodynamics, intraoperative findings or pathology evaluation) or requirement for intervention (thrombolysis, thrombectomy or urgent bypass).

Major thrombotic event

A composite dichotomous endpoint of pulmonary embolism, ischemic cerebrovascular event, myocardial infarction, or systemic arterial thromboembolism diagnosed at any time during the index hospitalization.

All thrombotic events

A composite dichotomous endpoint of deep vein thrombosis, pulmonary embolism, ischemic cerebrovascular event, myocardial infarction, or systemic arterial thromboembolism diagnosed at any time during the index hospitalization or death in hospital.

### Section 3 – Supplemental Results

#### Site Participation in the Moderate State Therapeutic Anticoagulation Trial

During the study period, April 21, 2020, to January 22, 2021, 121 sites in 9 countries (the United States, Canada, the United Kingdom, Brazil, Mexico, Nepal, Australia, the Netherlands, and Spain) enrolled patients into the moderate state therapeutic heparin multiple platform trial.

#### Additional Analyses

**Table S1** presents data from the planned adaptive analyses on January 13, 2021, which included 1398 moderate disease severity participants for whom the primary outcome was available. At this adaptive analysis, the posterior probability of superiority for therapeutic anticoagulation with heparin compared with usual care pharmacological thromboprophylaxis exceeded the pre-specified thresholds for superiority (>99% posterior probability of odds ratio >1.0) in both the high D-dimer (observed posterior probability of superiority 99.1%) and low D-dimer (observed posterior probability of superiority 99.7%) groups. On the basis of these planned results, the DSMB recommended halting enrollment, which occurred on January 22, 2021. **Table S2** presents the baseline clinical characteristics by randomization arm within the D-dimer-defined moderate disease severity participant groups. **Table S3** presents participants' initial antithrombotic agent and categorized dose equivalent, based on the dose administered on the first 24-48 hours following randomization. **Table S4** presents sensitivity analyses of the primary outcome. **Table S5** presents secondary outcomes, including in-hospital mortality, 28-day mortality, hospital length of stay, survival free of invasive mechanical ventilation or death through 28 days, survival free of organ support or death through 28 days, mechanical respiratory support-free days through 28 days, survival to hospital discharge without major thrombotic events or death, survival to hospital discharge without any macrovascular thrombotic event or death, and freedom from major bleeding. **Table S6** presents thrombotic events confirmed by adjudication by treatment group. **Table S7** presents the component criteria on which major bleeding events were confirmed as meeting the definition of a major bleed proposed by the International Society on Thrombosis and Haemostasis by treatment arm.

### Section 4 – Supplemental Tables

#### Table S1 – Results of the Planned Adaptive Analysis of 1398 Participants with Moderate Disease Severity on January 13, 2021, in the Low and High D-dimer Stopping Groups.

| Primary outcome of organ-support-free days | Adjusted odds ratio (95% credible interval)[a] | Posterior probability of superiority of therapeutic anticoagulation |
|---|---|---|
| High D-dimer group[b] | 1.53 (1.09-2.17) | 99.1% |
| Low D-dimer group[b] | 1.57 (1.14-2.19) | 99.7% |

Footnotes:

**a.** effect estimates are adjusted for age, sex, site, D-dimer group, and time epoch; an odds ratio greater than 1 indicates benefit from therapeutic anticoagulation; models incorporate dynamic borrowing (including the possibility of borrowing from 1123 severe disease severity participants, although given observed divergent treatment effects the extent of this was minimal);

**b.** analysis population (total moderate disease severity 1772) included 1398 for whom the primary outcome was available; only non-negative SARS-CoV-2 testing were included, although testing information was not available for a subset; sample size per group included 807 moderate disease severity participants with low D-dimer (<2 times local upper limit of normal for assay) and 471 with high D-dimer (≥2 times local upper limit of normal for assay); D-dimer was not known for 494 moderate disease severity participants – no pre-specified stopping rules were pre-specified in moderate participants with unknown D-dimer, although these participants' information could contribute to adaptive stopping decisions through dynamic borrowing of information on treatment effect in participants with an unknown D-dimer by the group with participants with known D-dimer when treatment effects were similar; when treatment effects are similar, more borrowing occurs.

#### Table S2 – Demographic and Clinical Characteristics of the Patients at Baseline.

| | Baseline D-dimer ≥2 times ULN | | Baseline D-dimer <2 times ULN | | Baseline D-dimer unknown | |
|---|---|---|---|---|---|---|
| **Characteristic** | **Therapeutic heparin (N=343)** | **Usual care thromboprophylaxis (N=292)** | **Therapeutic heparin (N=576)** | **Usual care thromboprophylaxis (N=505)** | **Therapeutic heparin (N=262)** | **Usual care thromboprophylaxis (N=253)** |
| Age – year | 63.1 (13.5) | 62.9 (12.9) | 56.6 (13.8) | 56.1 (13.5) | 59 (14.1) | 59.6 (14.6) |
| Male Sex – no. (%) | 202/343 (58.9) | 154/292 (52.7) | 357/576 (62) | 295/505 (58.4) | 154/262 (58.8) | 148/253 (58.5) |
| Race | | | | | | |
| White – no. (%) | 161/290 (55.5) | 146/239 (61.1) | 332/509 (65.2) | 304/436 (69.7) | 129/195 (66.2) | 114/170 (67.1) |
| Asian – no. (%) | 6/290 (2.1) | 13/239 (5.4) | 29/509 (5.7) | 25/436 (5.7) | 6/195 (3.1) | 5/170 (2.9) |
| Black – no. (%) | 85/290 (29.3) | 53/239 (22.2) | 95/509 (18.7) | 77/436 (17.7) | 39/195 (20) | 32/170 (18.8) |
| First Nations/ American Indian – no. (%) | 30/288 (10.4) | 23/237 (9.7) | 64/503 (12.7) | 37/431 (8.6) | 24/174 (13.8) | 22/151 (14.6) |
| Other – no. (%) | 8/304 (2.6) | 7/255 (2.7) | 8/545 (1.5) | 6/467 (1.3) | 1/260 (0.4) | 3/246 (1.2) |
| Ethnicity | | | | | | |
| Hispanic or Latino – no. (%) | 171/317 (53.9) | 157/272 (57.7) | 295/516 (57.2) | 279/449 (62.1) | 108/171 (63.2) | 101/158 (63.9) |
| Body mass index, kg/m2 | 29.4 (26–33.8) N = 281 | 29.4 (26.4–33.4) N = 231 | 30.4 (26.8–35.6) N = 502 | 30.8 (27–35) N = 430 | 29.1 (25.9–34.4) N = 196 | 30.7 (27–36.2) N = 199 |
| Pre-existing conditions | | | | | | |
| Hypertension – no. (%) | 191/315 (60.6) | 164/268 (61.2) | 260/532 (48.9) | 195/466 (41.8) | 95/176 (54) | 88/158 (55.7) |
| Diabetes mellitus – no. (%) | 119/343 (34.7) | 97/292 (33.2) | 157/576 (27.3) | 135/505 (26.7) | 76/262 (29) | 79/252 (31.3) |
| Severe cardiovascular disease[a] – no. (%) | 48/330 (14.5) | 55/286 (19.2) | 50/574 (8.7) | 41/502 (8.2) | 25/261 (9.6) | 25/250 (10) |
| Chronic kidney disease – no. (%) | 33/342 (9.6) | 32/288 (11.1) | 33/571 (5.8) | 16/502 (3.2) | 17/260 (6.5) | 21/247 (8.5) |
| Chronic respiratory disease[b] – no. (%) | 70/319 (21.9) | 58/262 (22.1) | 115/566 (20.3) | 99/491 (20.2) | 64/247 (25.9) | 55/235 (23.4) |
| Chronic liver disease – no. (%) | 5/323 (1.5) | 4/268 (1.5) | 7/565 (1.2) | 3/495 (0.6) | 2/261 (0.8) | 4/249 (1.6) |
| Immunosuppressive disease – no. (%) | 39/319 (12.2) | 39/262 (14.9) | 50/564 (8.9) | 43/494 (8.7) | 16/260 (6.2) | 21/249 (8.4) |
| Baseline treatments | | | | | | |
| Antiplatelet agent[c] – no. (%) | 65/334 (19.5) | 41/286 (14.3) | 59/561 (4.5) | 45/498 (9) | 24/245 (9.8) | 25/229 (10.9) |
| Remdesivir – no. (%) | 140/343 (40.8) | 117/292 (40.1) | 213/576 (37) | 187/505 (37) | 75/259 (29) | 79/251 (31.5) |
| Corticosteroids – no. (%) | 114/208 (54.8) | 87/155 (56.1) | 246/393 (62.6) | 196/306 (64.1) | 119/190 (62.6) | 132/195 (67.7) |

| | | | | | | |
|---|---|---|---|---|---|---|
| Tocilizumab – no. (%) | 1/343 (0.3) | 2/292 (0.7) | 3/576 (0.5) | 3/505 (0.6) | 2/259 (0.8) | 2/251 (0.8) |
| Baseline acute Respiratory support | | | | | | |
| None – no. (%) | 41/343 (12) | 33/292 (11.3) | 88/576 (15.3) | 66/505 (13.1) | 27/262 (10.3) | 24/253 (9.5) |
| Low flow nasal cannula/face mask – no. (%) | 250/343 (72.9) | 213/292 (72.9) | 408/576 (70.8) | 365/505 (72.3) | 131/262 (50) | 118/253 (46.6) |
| High flow nasal cannula – no. (%) | 6/343 (1.7) | 6/292 (2.1) | 10/576 (1.7) | 9/505 (1.8) | 9/262 (3.4) | 13/253 (5.1) |
| Non-invasive mechanical ventilation – no. (%) | 6/343 (1.7) | 7/292 (2.4) | 4/576 (0.7) | 7/505 (1.4) | 11/262 (4.2) | 10/253 (4) |
| Unspecified[d] – no. (%) | 40/343 (11.7) | 33/292 (11.3) | 66/576 (11.5) | 58/505 (11.5) | 84/262 (32.1) | 88/253 (34.8) |
| Laboratory values | | | | | | |
| D-dimer relative to site ULN | 3.2 (2.4–5.2)<br>N =342 | 3.3 (2.5–5)<br>N = 292 | 1.1 (0.8–1.5)<br>N = 558 | 1.1 (0.8–1.4)<br>N = 487 | -- | -- |
| Platelets x10$^9$/L | 234 (170.5–307.5)<br>N = 339 | 231.8 (174–324.2)<br>N = 288 | 216 (165–283)<br>N = 569 | 214 (173–274)<br>N =501 | 224 (181.8–289.8)<br>N = 252 | 217.5 (168.2–286)<br>N = 242 |
| INR | 1.1 (1–1.2)<br>N = 137 | 1.1 (1–1.1)<br>N =102 | 1 (1–1.1)<br>N =266 | 1 (1–1.1)<br>N=209 | 1.1 (1–1.1)<br>N = 72 | 1.1 (1–1.1)<br>N = 86 |
| Neutrophils x10$^9$/L | 5.6 (3.6–7.9)<br>N = 295 | 5.6 (3.9–8.1)<br>N =244 | 5.1 (3.4–7.8)<br>N = 527 | 4.8 (3.4–7.1)<br>N = 460 | 5.8 (3.6–8)<br>N = 200 | 5.7 (3.8–7.7)<br>N = 198 |
| Lymphocytes x10$^9$/L | 0.9 (0.6–1.2)<br>N = 300 | 1 (0.6–1.3)<br>N =244 | 0.9 (0.7–1.3)<br>N = 529 | 1 (0.7–1.4)<br>N = 463 | 1 (0.7–1.4)<br>N = 203 | 0.9 (0.7–1.4)<br>N = 201 |
| Creatinine (mg/dL) | 1 (0.8–1.3)<br>N = 336 | 0.9 (0.8-1.3)<br>N =285 | 0.8 (0.7–1)<br>N = 567 | 0.8 (0.7–1)<br>492 | 0.9 (0.7–1.1)<br>N = 241 | 0.9 (0.7–1.1)<br>N = 235 |
| Trial platform of enrollment[e] | | | | | | |
| ATTACC[f] – no. (%) | 192/343 (56.0) | 137/292 (46.9) | 351/576 (60.9) | 270/505 (53.5) | 107/262 (40.8) | 102/253 (40.3) |
| ACTIV-4a – no. (%) | 135/343 (39.4) | 137/292 (46.9) | 183/576 (31.8) | 199/505 (39.4) | 69/262 (26.3) | 56/253 (22.1) |
| REMAP-CAP – no. (%) | 16/343 (4.7) | 18/292 (6.2) | 42/576 (7.3) | 36/505 (7.1) | 86/262 (32.8) | 95/253 (37.5) |
| Country of enrollment | | | | | | |
| United States – no. (%) | 204/343 (59.5) | 176/292 (60.3) | 282/576 (49.0) | 248/505 (49.1) | 87/262 (33.2) | 82/252 (32.5) |
| United Kingdom – no. (%) | 12/343 (3.5) | 15/292 (5.1) | 29/576 (5.0) | 29/505 (5.7) | 54/262 (20.6) | 59/252 (23.4) |
| Canada – no. (%) | 37/343 (10.8) | 24/292 (8.2) | 45/576 (7.8) | 43/505 (8.5) | 20/262 (7.6) | 16/252 (6.3) |
| Brazil – no. (%) | 59/343 (17.2) | 42/292 (14.4) | 129/576 (22.4) | 123/505 (24.4) | 46/262 (17.6) | 44/252 (17.5) |
| Other[g] – no. (%) | 31/343 (9) | 35/292 (12) | 91/576 (15.8) | 62/505 (12.3) | 55/262 (21) | 51/252 (20.2) |

Median [IQR] or proportions. **Abbreviations**: No. = number; ULN = upper limit of normal.

**Footnotes**:

**a**. defined in ACTIV-4a and ATTACC as a baseline history of heart failure, myocardial Infarction, coronary artery disease, peripheral arterial disease, or cerebrovascular disease (stroke or transient ischemic attack, and defined in REMAP-CAP as a baseline history of New York Heart Association class IV symptoms;
**b**. defined as a baseline history of asthma, chronic obstructive pulmonary disease, bronchiectasis, interstitial lung disease, primary lung cancer, pulmonary hypertension, active tuberculosis, or through the receipt of home oxygen therapy;
**c**. not included in this summary are 74 participants co-enrolled in the REMAP-CAP Antiplatelet Domain (39 assigned to therapeutic anticoagulation, 35 to usual care thromboprophylaxis);
**d**. in REMAP-CAP, levels of oxygen support, including no support, less than high flow nasal cannula were not differentiated;
**e**. ATTACC implemented response-adaptive randomization on December 15, 2020, which led to imbalanced randomization;
**f**. 215 participants from the United States enrolled in the ATTACC platform were funded under the ACTIV-4 platform by the National Heart Lung and Blood Institute of the NIH;
**g.** other participating countries included: Mexico, Nepal, Australia, the Netherlands, and Spain.

#### Table S3 – Heparins Utilized and Dosage Adherence.[a]

| | Therapeutic anticoagulation N=1181 | Usual care pharmacological thromboprophylaxis N=1050 |
|---|---|---|
| Anticoagulant drug – no. (%) | N= 1093 | N=799 |
| Enoxaparin | 921 (84.3) | 629 (78.7) |
| Dalteparin | 87 (8.0) | 77 (9.6) |
| Tinzaparin | 27 (2.5) | 26 (3.3) |
| Subcutaneous unfractionated heparin | 11 (1.0) | 49 (6.1) |
| Intravenous unfractionated heparin | 41 (3.8) | 4 (0.5) |
| Direct oral anticoagulant | 0 (0) | 12 (1.5) |
| Other | 6 (0.5) | 2 (0.3) |
| Dosage equivalents – no. (%) | N=1043 | N=855 |
| Low dose thromboprophylaxis | 61 (5.8) | 613 (71.7) |
| Intermediate dose thromboprophylaxis | 61 (5.8) | 227 (26.5) |
| Subtherapeutic dose anticoagulation | 91 (8.7) | 7 (0.8) |
| Therapeutic dose anticoagulation | 830 (79.6) | 8 (0.9) |

**Abbreviations**: No. = number.

**Footnotes**:

**a**. data reported reflects participants in whom dosing information was available at the time the dataset was locked for analysis; drug and dose reported are those prescribed in the first 24-48 hours following randomization, classified using a consensus dose categorization (see **Supplementary Appendix Section 2**, p. 48).

#### Table S4 – Sensitivity Analyses of the Primary Outcome among All Moderate Participants.[a]

| Sensitivity Analysis | Therapeutic anticoagulation (no.) | Usual care pharmacological Thromboprophylaxis (no.) | Adjusted median proportional odds ratio (95% CrI)[b] | Posterior probability of superiority of therapeutic anticoagulation |
|---|---|---|---|---|
| Examining the primary outcome as a three-category ordinal outcome[c] | | | | |
| | 1171 | 1048 | 1.29 (1.04 to 1.61) | 99.1% |
| Permitting dynamic borrowing of information on treatment effect from severe participants | | | | |
| | 1171 | 1048 | 1.28 (1.03 to 1.58) | 98.6% |
| Including participants with both confirmed (based on microbiological testing for SARS-CoV-2) and suspected (not confirmed) Covid-19 | | | | |
| | 1180 | 1051 | 1.29 (1.04 to 1.60) | 98.9% |
| Removing site and time effects from the model | | | | |
| | 1171 | 1048 | 1.26 (1.03 to 1.55) | 98.6% |
| Excluding participants receiving concomitant antiplatelet agents as part of usual care at time of enrollment and participants co-randomized to the antiplatelet domain of REMAP-CAP | | | | |
| | 992 | 902 | 1.24 (0.98 to 1.57) | 96.1% |
| Excluding participants randomized after January 7, 2021[d] | | | | |
| | 968 | 895 | 1.36 (1.08 to 1.73) | 99.5% |

**Abbreviations:** CrI = credible interval; no. = number.

**Footnotes**:

**a.** analysis of all moderate participants in the modified intention-to-treat population, assuming the same treatment effect irrespective of D-dimer;

**b.** effect estimates are adjusted for age, sex, site, D-dimer group, and time epoch; an odds ratio greater than 1 indicates benefit from therapeutic anticoagulation; all models, except where indicated, assume independent treatment effects within groups (no dynamic borrowing);

**c.** levels of the ordinal outcome are: 1) alive and free of organ support through day 28; 2) alive with organ support by day 28; and 3) death by day 28;

**d.** the adaptive stopping conclusions were publicly announced on January 22, 2021, at which time a number of participants were still within the treatment window; to exclude the potential influence of possible treatment cross-over among study participants who were still in the treatment window, this sensitivity analysis restricted to participants enrolled on or before January 7, 2021, who would have had up to the maximum 14 days to complete the protocolized intervention prior to public announcement of the adaptive analysis results.

#### Table S5 – Secondary Outcomes.

| Secondary outcome Moderate analysis group[a] | Therapeutic anticoagulation N/N (%)[b] | Usual care venous Thromboprophylaxis N/N (%)[b] | Adjusted odds or hazard ratio (95% CrI)[c] | Posterior probability of superiority of therapeutic anticoagulation |
|---|---|---|---|---|
| Survival to hospital discharge[d] | | | | |
| High D-dimer group | 305/339 (90.0) | 260/291 (89.3) | 1.16 (0.79-1.71) | 78.6% |
| Low D-dimer group | 538/570 (94.4) | 477/505 (94.5) | 1.19 (0.81-1.74) | 81.7% |
| Unknown D-dimer group | 242/262 (92.4) | 225/252 (89.3) | 1.20 (0.81-1.83) | 82.1% |
| All moderate participants[e] | 1085/1171 (92.7) | 962/1048 (91.8) | 1.21 (0.87, 1.68) | 87.1% |
| 28-day survival[f] | | | | |
| All moderate participants[e] | - | - | 1.20 (0.88-1.61) | 87.8% |
| Survival without intubation through 28 days | | | | |
| High D-dimer group | 300/343 (87.5%) | 253/292 (86.6%) | 1.14 (0.71-1.84) | 71.3% |
| Low D-dimer group | 521/576 (90.5%) | 457/505 (90.5%) | 1.20 (0.80-1.83) | 80.9% |
| Unknown D-dimer group | 231/262 (88.2%) | 213/253 (84.2%) | 1.29 (0.79-2.18) | 84.3% |
| All moderate participants[e] | 1052/1181 (89.1%) | 923/1050 (87.9%) | 1.22 (0.93-1.59) | 92.2% |
| Survival without organ support 28 days[g] | | | | |
| High D-dimer group | 257/338 (76.0%) | 205/290 (70.7%) | 1.40 (0.96-2.03) | 96.0% |
| Low D-dimer group | 466/575 (81.0%) | 396/503 (78.7%) | 1.22 (0.90-1.65) | 89.4% |
| Unknown D-dimer group | 209/262 (79.8%) | 188/253 (74.3%) | 1.31 (0.87-1.99) | 90.2% |
| All moderate participants[e] | 932/1175 (79.3%) | 789/1046 (75.4%) | 1.30 (1.05-1.61) | 99.1% |
| Survival free of mechanical respiratory support free days (MRSFD=29[h]) | | | | |
| High D-dimer group | 282/343 (82.2%) | 238/292 (81.5%) | 1.16 (0.77-1.76) | 76.4% |
| Low D-dimer group | 494/576 (85.8%) | 431/505 (85.3%) | 1.16 (0.82-1.65) | 80.1% |
| Unknown D-dimer group | 218/262 (83.2%) | 195/253 (77.1%) | 1.39 (0.90-2.16) | 93.1% |
| All moderate participants[e] | 994/1181 (84.2%) | 864/1050 (82.3%) | 1.22 (0.97-1.55) | 95.3% |
| Survival to hospital discharge without major thrombotic events[i] | | | | |
| High D-dimer group | 305/343 (88.9%) | 253/292 (86.6%) | 1.29 (0.78-2.13) | 84.1% |
| Low D-dimer group | 540/576 (93.8%) | 473/505 (93.7%) | 1.27 (0.78-2.07) | 83.0% |
| Unknown D-dimer group | 241/261 (92.3%) | 216/249 (86.7%) | 1.64 (0.92-3.00) | 95.2% |
| All moderate participants[e] | 1086/1180 (92%) | 942/1046 (90.1%) | 1.39 (1.02-1.88) | 98.0% |
| Survival to hospital discharge without any macrovascular thrombotic events[j] | | | | |
| High D-dimer group | 304/343 (88.6%) | 253/292 (86.4%) | 1.27 (0.77-2.06) | 83.0% |
| Low D-dimer group | 540/576 (93.8%) | 472/505 (93.5%) | 1.31 (0.80-2.15) | 86.0% |
| Unknown D-dimer group | 240/261 (92.0%) | 213/249 (85.5%) | 1.72 (0.99-3.07) | 97.2% |
| All moderate participants[e] | 1084/1180 (91.9%) | 938/1046 (89.7%) | 1.41 (1.04-1.92) | 98.6% |
| Hospital length of stay[f] | | | | |
| All moderate participants[e] | - | - | 1.03 (0.94-1.13) | 72.7% |
| Freedom from major bleeding | | | | |
| High D-dimer group | 335/343 (97.7%) | 288/292 (98.6%) | 0.62 (0.23-1.67) | 17.7%[k] |
| Low D-dimer group | 564/576 (97.9%) | 503/505 (99.6%) | 0.43 (0.16-1.08) | 3.7%[k] |
| Unknown D-dimer group | 259/261 (99.2%) | 247/250 (98.8%) | 1.30 (0.36-4.83) | 65.5%[k] |
| All moderate participants[e] | 1158/1180 (98.1%) | 1038/1047 (99.1%) | 0.56 (0.27-1.11) | 4.8%[k] |

**Abbreviations:** CrI = credible interval; MRFSD = mechanical respiratory support free days. **Footnotes**:

**a.** High D-dimer group defined as moderate participants with baseline D-dimer ≥2 times local upper limit of normal for assay, Low D-dimer group defined as moderate participants with baseline D-dimer <2 times local upper limit of normal for assay, D-Dimer Unknown group defined as moderate participants without available baseline D-dimer;

**b.** unadjusted proportions; no. of participants/total no. (%);

**c.** effect estimates are adjusted for age, sex, site, D-dimer group, and time epoch; an odds ratio greater than 1 indicates benefit from therapeutic anticoagulation; models assume independent treatment effects within groups (no dynamic borrowing), except the survival to discharge outcome as this derives from the primary model;

**d.** model results from D-dimer groups incorporates dynamic borrowing across D-dimer groups and with severe patients; assuming independent effects within each group (without borrowing), the adjusted OR (95% CrI) for in-hospital mortality by groups were: High D-dimer group (aOR = 1.11 [95% CrI 0.65-1.88]; posterior probability of superiority 64.4%), Low D-dimer group (aOR = 1.21 [95% CrI 0.72-2.03]; posterior probability of superiority 76.1%), and D-dimer unknown (aOR = 1.31 [95% CrI 0.73-2.45]; posterior probability of superiority 80.6%);

**e.** analysis of all moderate participants assuming the same treatment effect irrespective of D-dimer, without dynamic borrowing;

**f.** a time-to-event endpoint;

**g.** a separate analysis was performed excluding patients on organ support at baseline: among all moderate participants without organ support at baseline, 907/1133 (80.1%) in the therapeutic arm and 762/998 (76.4%) in the usual care pharmacological thromboprophylaxis arm survived without organ support through 28-days, with an adjusted median proportional odds ratio (95% CrI) of 1.30 (1.06-1.62; posterior probability of superiority of therapeutic anticoagulation 99.3%);

**h.** MRSFD = 29 indicates no mechanical respiratory support through 28 days;

**i.** major thrombotic events was defined by a composite of myocardial infarction, pulmonary embolism, ischemic stroke, and systemic arterial embolism events;

**j.** defined by the composite of major thrombotic events plus deep venous thrombosis;

**k.** there is a 96.3%, 82.3% and 34.5% posterior probability that therapeutic anticoagulation is inferior to usual care thromboprophylaxis in the high D-dimer, low D-dimer, and unknown D-dimer groups, respectively; for the overall moderate cohort, there is a 95.2% posterior probability that therapeutic anticoagulation is inferior to usual care thromboprophylaxis;

**l.** a time-to-event endpoint censored at 28-days; the overall median (interquartile range) hospital length of stay following randomization was 5 (3, 10) days.

#### Table S6 – Confirmed Thrombotic Events Occurring during Index Hospitalization.[a]

| | Therapeutic anticoagulation | Usual care pharmacological thromboprophylaxis |
|---|---|---|
| Number of participants in whom a thrombotic event outcome was available | 1180 | 1046 |
| Number of patients with an in-hospital thrombotic event | 16 | 28 |
| Thrombotic events[b] | | |
| Total events | 19 | 31 |
| Pulmonary embolism | 10 | 19 |
| Myocardial infarction | 1 | 1 |
| Ischemic cerebrovascular event | 1 | 2 |
| Systemic arterial thromboembolism | 1 | 2 |
| Deep venous thrombosis[c] | 6 | 7 |

**Footnotes**:
**a**. thrombotic events were adjudicated in a blinded fashion by clinical endpoints committees using consensus definitions (see **Supplementary Appendix Section 2**, p. 50);
**b.** events are not mutually exclusive; total events are reported;
**c.** deep venous thrombosis was not included in the composite endpoint of major thrombotic events but is included in the all macrovascular thrombotic events endpoint.

#### Table S7 – Confirmed ISTH Major Bleeding Events.[a]

| | Therapeutic anticoagulation | Usual care pharmacological thromboprophylaxis |
|---|---|---|
| Number of patients in whom a major bleeding event outcome was available | 1180 | 1047 |
| Number of patients with a major bleeding event | 22 | 9 |
| Bleeding event breakdown[b] | | |
| Fatal bleeding | 3 | 1 |
| Overt and symptomatic bleeding in a critical area or organ | 9 | 1 |
| Intracerebral hemorrhage | 0 | 0 |
| Overt bleeding causing a fall in hemoglobin ≥ 2 g/dL or leading to transfusion of ≥ 2 units of whole blood or red cells | 27 | 14 |

**Abbreviations**: g/dL = grams per decilitre; ISTH = International Society on Thrombosis and Haemostasis.
**Footnotes:**
**a.** bleeding events were collected during the treatment window in both randomization arms, and adjudicated in a blinded fashion by clinical endpoints committees using the criteria outlines by the International Society on Thrombosis and Haemostasis for major bleeding in non-surgical patients (see **Supplementary Appendix Section 2**);
**b.** bleeding criteria are not mutually exclusive for confirmation of ISTH major bleeding, with one or more required to confirm.

### Section 5 – Supplemental Figures

#### Figure S1

Distribution of organ support free days (OSFD) for therapeutic-dose anticoagulation and usual care pharmacological thromboprophylaxis by D-dimer group. Organ support-free days are shown as horizontally stacked proportions by intervention group, with possible outcomes as: in-hospital death with or without the receipt of organ support (dark red; the worst possible outcome, corresponding to an ordinal scale score of -1); survival, requiring ICU-level organ support (red to blue gradient shading based on number of days alive without organ support; intermediate outcomes, corresponding to an ordinal scale scores of 0-21); and survival to hospital discharge, without requiring ICU-level organ support (dark blue; the best possible outcome, corresponding to an ordinal scale score of 22).

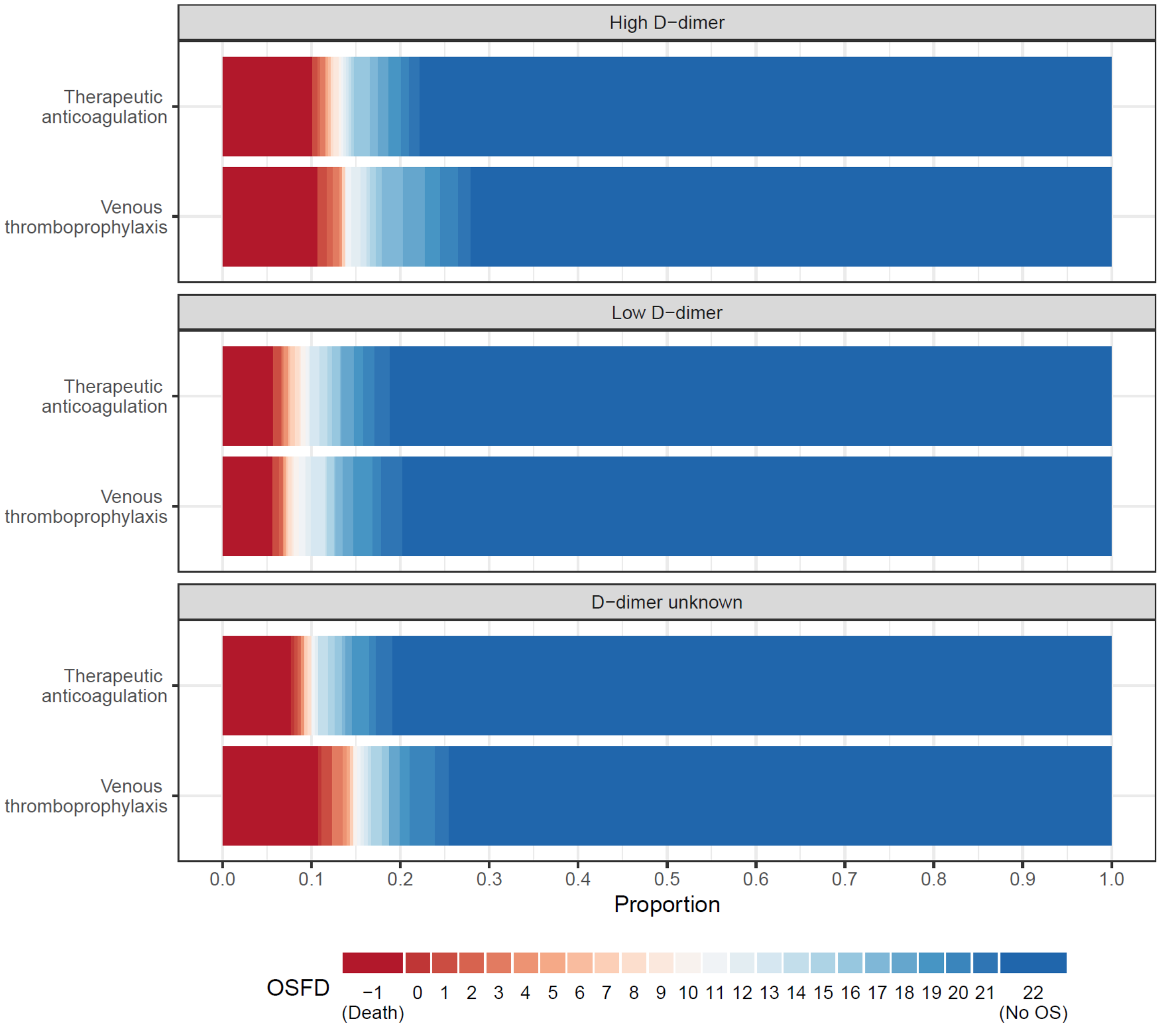

#### Figure S2

Subgroup analyses on the primary endpoint (organ support-free days to day 21) in participants with moderate Covid-19.

| Subgroup | # Patients | Odds Ratio Median (95% CrI) | Pr(Superior) | Sample size PTP | Sample size TAC |
|---|---|---|---|---|---|
| Overall | 2219 | 1.29 (1.04, 1.61) | 99.0 | 1048 | 1171 |
| Age | | | | | |
| <50 | 541 | 1.14 (0.69, 1.88) | 69.4 | 255 | 286 |
| 50−70 | 1216 | 1.29 (0.98, 1.72) | 96.4 | 578 | 638 |
| >70 | 462 | 1.36 (0.92, 2.01) | 93.6 | 215 | 247 |
| Sex | | | | | |
| Male | 1302 | 1.46 (1.13, 1.91) | 99.8 | 595 | 707 |
| Female | 917 | 1.01 (0.71, 1.45) | 52.4 | 453 | 464 |
| Baseline Respiratory Support | | | | | |
| None | 276 | 1.39 (0.70, 2.79) | 82.6 | 123 | 153 |
| Supplemental O2 | 1477 | 1.27 (0.99, 1.64) | 96.9 | 694 | 783 |
| HFNO | 53 | 0.54 (0.20, 1.43) | 10.2 | 28 | 25 |
| Ventilated | 45 | 1.34 (0.46, 4.21) | 70.9 | 24 | 21 |
| Unspecified | 368 | 1.33 (0.72, 2.49) | 81.6 | 179 | 189 |
| Baseline Antiplatelet | | | | | |
| No antiplatelet | 1958 | 1.25 (0.99, 1.57) | 96.6 | 935 | 1023 |
| Antiplatelet | 256 | 1.40 (0.79, 2.41) | 87.5 | 111 | 145 |
| PTP Classification | | | | | |
| Intermediate−dose | 897 | 1.39 (0.97, 2.00) | 96.6 | 450 | 447 |
| Low−dose | 465 | 1.30 (0.80, 2.09) | 85.4 | 177 | 288 |

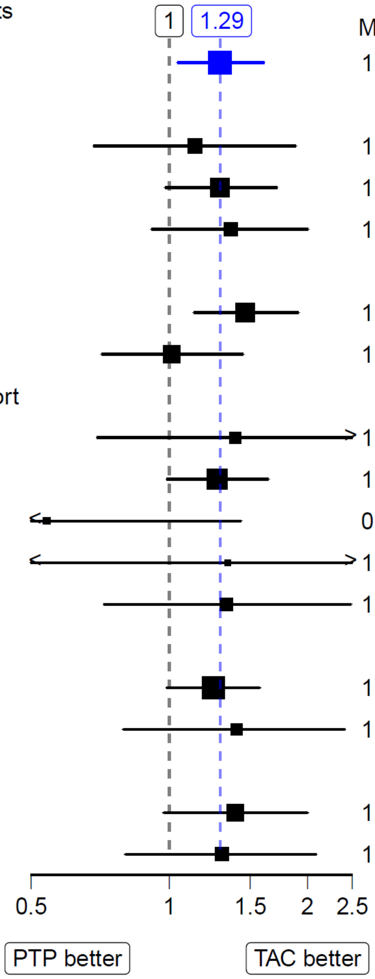

**Abbreviation**: CrI = credible interval; HFNO = high-flow nasal oxygen; Pr = probability; PTP = usual care pharmacological thromboprophylaxis; TAC = therapeutic anticoagulation with heparin.
**Footnotes**: Models are adjusted for age, sex, site, D-dimer group, and time; odds ratio greater than 1.0 favors therapeutic anticoagulation with heparin. Female sex was independently associated with higher organ support-free days (adjusted median odds ratio 1.87, 95% CrI 1.38, 2.56).

### Section 6 – References
